# The Phenome-wide Consequences of Anorexia Nervosa Genes

**DOI:** 10.1101/2021.02.12.21250941

**Authors:** Jessica S. Johnson, Alanna C. Cote, Amanda Dobbyn, Laura G. Sloofman, Jiayi Xu, Liam Cotter, Alexander W. Charney, Eating Disorders Working Group of the Psychiatric Genomics Consortium, Jennifer Jordan, Martin Kennedy, Mikael Landén, Sarah L Maguire, Nicholas G Martin, Preben Bo Mortensen, Cynthia M. Bulik, Laura M. Huckins

## Abstract

Anorexia nervosa (AN) is a psychiatric disorder with complex etiology, with a significant portion of disease risk imparted by genetics. Traditional GWAS studies produce principal evidence for the association of genetic variants with disease, and provide a jumping-off point for downstream functional analyses. Transcriptomic imputation (TI) allows for the translation of SNPs into regulatory mechanisms, which can then be used to assess the functional outcome of genetically regulated gene expression (GReX) in a more broad setting through the use of phenome-wide association studies (PheWAS) in large and diverse clinical biobank populations with electronic health record (EHR) phenotypes. Here, we applied TI using S-PrediXcan to translate the most recent PGC-ED AN GWAS findings into AN-GReX. For significant genes, we imputed AN-GReX in the Mount Sinai Bio*Me*™ Biobank and performed PheWAS on over 2000 clinical outcomes to test the clinical consequences of aberrant expression of these genes. We performed a secondary analysis to assess the impact of BMI on AN-GReX clinical associations.

Our S-PrediXcan analysis identified 47 genes associated with AN, including what is, to our knowledge, the first genetic association of AN with the Major Histocompatibility Complex (MHC). AN-GReX was associated with autoimmune, anthropometric, metabolic, psychiatric and gastrointestinal diagnoses in our biobank cohort, as well as measures of anthropometry, substance use, and pain score. Our analyses reveal that AN-GReX associations with measures of weight and substance use are modified by BMI, and indicate potential avenues of functional mechanism to investigate further.

## INTRODUCTION

Anorexia nervosa (AN) is a severe eating disorder characterized by extreme low body weight and fear of gaining weight, and is often accompanied by compensatory behaviors to lose weight such as dietary restriction, purging, and higher than average physical activity(1). With a lifetime prevalence of 0.9-4%, AN has high mortality, with few effective treatments and an increased risk of suicide(2–4). Clinical and epidemiological studies have shown higher risk of other psychiatric disorders such as OCD, anxiety, and major depression in individuals with AN(3,5), and higher risk of substance use disorders in AN subtypes (6), as well as higher risk of metabolic and autoimmune disorders such as type 1 diabetes(7,8),. Twin studies have established the heritability of AN to be between 50 to 60%, indicating a considerable contribution of genetic factors to AN disease liability(9,10).

Both metabolic and psychiatric factors contribute to anorexia nervosa. Epidemiological studies indicate high rates of psychiatric comorbidity, particularly with depression, anxiety disorders, and OCD(5), and many of these disorders share substantial symptomatology and environmental risk factors(11). Evidence is mounting that AN may lie at the extreme of a BMI spectrum, with obesity lying at the opposite end, and that AN may be a disorder of “fundamental metabolic dysregulation”(2). In line with this hypothesis, individuals with AN are able to reach and maintain incredibly low body weight, and often revert to this weight even after re-nourishment treatment, possibly indicating a fundamentally low BMI biological settling point(12,13). Additionally, individuals with AN have been shown to have lower concentrations of leptin, a hormone involved in regulation of body weight and other metabolic processes(13–16). Microbiome studies have revealed abnormal microflora in the gut of AN patients(17), and evidence from many studies have pointed to a possible brain-gut etiology of AN(9,18–20).

Genetic studies of AN have provided further evidence of both psychiatric and metabolic etiology. The largest AN GWAS to date (N_Cases_=16992) uncovered eight loci associated with AN risk(9), and determined SNP-based heritability (h^2^_SNP_) of 11-17%, similar to other psychiatric disorders, indicating that common variants contribute to polygenic risk of AN. In addition, genetic correlations demonstrate significant overlap with other psychiatric disorders, including schizophrenia(9,21), MDD(9), OCD(9), alcohol use disorder(22) and anxiety disorders(9), as well as physical activity(9,23), indicating shared genetic variation between these traits. Significant negative genetic correlation between AN and anthropometric and metabolic traits, such as BMI(9,21,23), fat mass(9,21,23), obesity(9), type 2 diabetes(9), insulin resistance(9,21), fasting insulin(9,21) and leptin(9) have also been observed, further indicating metabolic components to AN disease risk. One study of AN polygenic risk scores (PRS) has indicated additional genetic associations of AN risk variants with anthropometric, behavioral, and psychiatric traits(24).

Traditional observational genetic studies provide substantial data for factors contributing to risk of a particular disease, but may not capture the genetic factors that are specifically contributing to subthreshold or prodromal disease states. Electronic Health Records (EHR) contain information about an individual’s health history such as diagnoses, medications, laboratory tests, vital signs, and family medical history, and can be utilized for scalable disease research. The vast amounts of data from the EHR can be queried to provide an understanding of the clinical spectrum of disease and disease progression across the lifetime of the patient. The initiation of biobanks to collect biological specimens from large numbers of patients within a healthcare system allows us to connect massive amounts of phenotype data from the EHR to genetic data. A phenome-wide association study or PheWAS is an effective method of querying the EHR to look for the associations of a trait of interest with the clinical phenome,(25–27). Previous studies have demonstrated the efficacy of PheWAS methods in determining clinical outcomes associated with genomic disease risk variants, and have replicated associations from GWASs of strictly defined disease traits, including psychiatric disorders(28–30),. Exploring the phenotypic associations with AN genetic architecture has the potential to clarify how some of these GWAS variants functionally contribute to AN disease risk, symptomatology, and clinical presentation.

Transcriptomic imputation (TI) approaches leverage expression quantitative trait loci (eQTLs) derived from large, well-curated gene expression datasets (for example, the Genotype-Tissue Expression project (GTEx)(31), CommonMind Consortium (CMC)(32), and Depression Genes and Networks (DGN)(33)) in order to derive gene expression predictor models. These models may be applied to predict genetically regulated gene expression (GReX) in large genotyped cohorts, without the need to collect tissue samples or RNA(19,34–36). Here, we explored the association between AN-associated GReX and the clinical phenome. We performed transcriptomic imputation using S-PrediXcan on the most recent PGC-ED AN GWAS to first find GReX associated with AN, and then tested for clinical associations of these genes with structured EHR-encoded phenotypes using PheWAS. Given the relationship of AN with BMI, we further investigated the effect of BMI on the GReX-phenotype associations by stratifying biobank individuals based on sex- and ancestry-adjusted categories of low, mid, and high BMI. Understanding the clinical consequences of aberrant gene expression across the phenome may clarify the biological mechanisms of clinically relevant AN GWAS risk variants.

## METHODS

### AN GENELIST and TRANSCRIPTOMIC IMPUTATION

We performed transcriptomic imputation using S-PrediXcan(35) on the largest available summary statistics of Anorexia Nervosa (AN) (16992 AN cases and 55525 controls) downloaded from the Psychiatric Genomics Consortium (PGC) website(9). We tested for association of genetically-regulated gene expression (GReX) using available GTEx, CMC and DGN predictor models(34–36) for a total of 50 different tissues with AN case-control status. We established two thresholds for significance; first, correcting for all genes tested within each tissue (p_tissue_, values shown in **Table S1**, and second, correcting for all tissues and genes tested (p_bonferroni_=3.75 × 10^−8^).

### Bio*Me™*

The Bio*Me™* Biobank at the Icahn School of Medicine at Mount Sinai includes genotype and EHR data from 31704 individuals recruited from the general patient population of the Mount Sinai healthcare system. Individuals were genotyped on the Illumina Global Screening Array (GSA), an array designed for multi-ethnic populations. The Bio*Me™* population is very diverse, with the proportion of major ancestral population groups representative of the diversity of New York City and Mount Sinai’s patient population: Hispanic (HA, 35%), European (EA, 34%), African (AA, 25%), East Asian (ESA, 3%), South Asian (SAS, 2%), and Native American (NA, 0.3%) (Full Bio*Me™* demographics are shown in **Table 1**). Quality control and imputation of the genotyping data for Bio*Me™* is described in elsewhere(28).

| BioMe BioBank Demographic Information |  |  |  |  |  |  |
| --- | --- | --- | --- | --- | --- | --- |
|  |  | Total N | N Self-report Female (%) | N Self-report Male (%) | Mean Age (1stQ-3rdQ) | Mean BMI (1stQ-3rdQ) |
| Total Cohort |  | 31585 | 17968 (57) | 12417 (39) | 57.12 (45-69) | 28.69 (27.46-32.09) |
| Ancestry | AA | 8044 | 4945 (61) | 2937 (37) | 55.76 (45-66) | 30.63 (25.06-34.81) |
|  | EA | 10788 | 5273 (49) | 4766 (44) | 58.79 (45-72) | 26.58 (22.64-29.39) |
|  | ESA | 957 | 567 (59) | 332 (35) | 50 (38-60) | 24.26 (21.22-26.21) |
|  | HA | 11047 | 6863 (62) | 3980 (36) | 57.42 (45-70) | 29.64 (25.17-32.95) |
|  | SAS | 661 | 275 (42) | 360 (54) | 52.93 (39-66) | 26.14 (23.04-28.52) |
|  | NA | 88 | 45 (51) | 42 (48) | 57.03 (46.5-69) | 28.35 (24.24-31.43) |

Ancestry for Bio*Me™* was initially assigned from self-reported values into six ancestral groupings: African (AA), European (EA), Hispanic (HA), East Asian (ESA), Native American (NA), and O (Other). We plotted the first two principal components from the genotyping to confirm ancestral grouping. For individuals not clearly falling in an ancestral group, and for those designated “Other”, we merged genotype information with phase 3 of 1000Genomes and reran the principal components analysis (PCA) using PLINK(37,38) in order to assign individuals to 1000Genomes super-populations. From this we were able to assign individuals to one of six ancestral groups: African (AA), European (EA), East Asian (ESA), Hispanic (HA), Native American (NA), and South Asian (SAS). A total of 119 samples that could not be grouped were removed, leaving 31585 individuals for analysis.

### PheWAS

We calculated GReX for all *p*_*tissue*_ significant genes across GTEx(31), DGN(33), and CMC(32,39) tissues (N=50) in Bio*Me™* individuals using PrediXcan-2(34,35) software, and performed a phenome-wide association (PheWAS) analysis. Logistic regression between the calculated AN-GReX and phecodes was performed using the PheWAS R package(40), adjusting for age, biological sex, and the first five genotype-derived principal components. PheWAS were run per Bio*Me™* ancestry cohort of (N_Ancestry_=6) and results meta-analyzed using an inverse-variance based approach in METAL(41). Due to the nature of EHR data, sample sizes can vary greatly depending on the phenotype, which can, in turn, affect our ability to detect associations. We believe our study is sufficiently powered given results from a previous study(42) that used simulations of MAF, penetrance and sample number parameters to estimate power for pheWAS studies.

#### Encounter Diagnosis

Current diagnoses for each individual were recorded at each visit using the International Classification of Disease (ICD) coding system and recorded as “Encounter Diagnosis”. Phecodes were assigned from Encounter Diagnosis information by grouping ICD-9 and ICD-10 diagnostic codes(43). Individuals with at least two counts of a particular diagnostic code were considered “cases”, those with zero counts were marked as “controls”, and those with only one count were set to missing. After QC, there were a total of 2178 unique Encounter Diagnosis codes and 1093 unique phecodes in the dataset.

For both encounter diagnoses and phecodes, we required at least 10 occurrences of each phenotype within each population group for inclusion in the analysis, and an overall effective sample size N_*eff*_ > 100 (**Equation 1**).

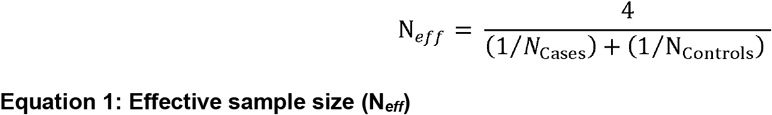

Due to the high correlation between Encounter Diagnosis and phecode files, we combined all results from both PheWAS and performed an FDR correction in *R* to determine overall significant associations with diagnostic phenotypes. Significant associations were set at an FDR-corrected p threshold of 0.05.

### Bio*Me*-R Structured Phenotype Database (BRSPD) PheWAS

In addition to diagnosis code data, we also had access to other types of EHR-derived data that included information on allergy status, vital signs (weight, height, blood pressure, pulse, pulse oximetry, respirations, and temperature), family history, personal history, medication use, obstetrics and gynecology outcomes (OB/GYN), and other social and behavioral traits. This information derived from the EHR was collected into files under the heading BRSPD or Bio*Me*-R Structured Phenotype Database. Summaries and descriptive statistics of all of the BRSPD phenotypes can be found in **Supplementary Methods**. For all BRSPD traits, we required at least ten cases in each ancestry group, and a total N_eff_>100 for inclusion in our analysis.

#### Phenotype QC and Analysis for Continuous Traits

We plotted the distribution of continuous traits across the cohort, and removed implausible or impossible values (likely due to errors in entry; for example, height listed as 1745 inches). Continuous traits were then analyzed as follows:

1. Height (cm): mean measure per individual.
2. Weight (kg): (i) highest recorded weight, (ii) lowest recorded weight, (iii) weight change over time. Weight change over time was calculated by taking the difference between the highest and lowest recorded weights, divided by the number of years between those two measures.
3. Pain scores were self-reported on a 1 to 10 scale and assessed for 25 different body locations. First, we tested pain score regardless of location, using three measures: (i) highest ever pain score, (ii) mean pain score, (iii) total pain score (sum), to encompass both acute and chronic pain conditions. We then repeated this analysis to each reported pain location, with the same three measures.
4. Measures of alcohol, tobacco, and illicit drug use were categorized as follows: alcohol-ounces (oz) per day; tobacco- (i) packs per day and (ii) years used; illicit drug use-frequency of use.
5. Vital signs (blood pressure, pulse, pulse oximetry, respiration, temperature (Celsius)) were analyzed using (i) highest ever value, (ii) lowest ever value, (iii) mean, (iv) variance per individual.

#### Categorical Traits

Categorical traits were converted to binary TRUE/FALSE values (phenotypes included are shown in **Supplemental Information)**. For allergy and medication phenotypes, we collapsed and combined data on equivalent root components and/or active ingredients (e.g., allergies for the antibiotic Vancomycin were coded as “VANCOMYCIN”, “VANCOMYCIN HCL”, “VANCOMYCIN IN D5W”, “VANCOMYCIN IN 0.9% SODIUM CL”, “VANCOMYCIN IN DEXTROSE ISO-OSM”, and “VANCOMYCIN (BULK)”), removed extraneous symbols and corrected typing errors. A full account of QC steps is documented in the **Supplementary Methods**.

### STRATIFICATION BY BMI

Given the nature of AN symptomology and association with weight phenotypes, we tested whether AN-GReX associations varied based on BMI. We stratified Bio*Me™* individuals into three main BMI categories, defined based on ancestry- and sex-specific BMI distributions within our data (**Figures S1 and S2; Tables S2 and S3)**. BMI values used for these analyses were measured at intake into Bio*Me*™. We categorized individuals with BMI within the 1^st^ quartile as “Low” BMI; those above the 3^rd^ quartile as “High” BMI, and those falling between the 1^st^ and 3^rd^ quartiles as “Mid” BMI (**Figure S1**). We then repeated our analysis in these three groups for all significant associations from our full analysis.

#### Testing for hidden case contamination

It is possible that some of the clinical associations within our study stem from undiagnosed ED- and AN-cases, rather than a direct effect of gene expression on phenomic expression. We term this “diagnostic contamination”. To test whether this effect may drive the associations we observe, we simulated the effects of diagnostic contamination occurring at rate *p* within our biobank sample, with effect *β* on GReX.

Full derivations and simulations are shown in our **Supplementary Material**; briefly, we derive the expression among controls, and cases contaminated at rate *p*; the difference in expression between the two groups; the expected variance among cases, and pooled across all samples, and the expected statistical significance (T-score, and p-value). Next, we simulated gene expression distributions for (i) 1000 cases and 1000 controls; (ii) 1000 cases and 10000 controls; (iii) 1000 cases and 30000 controls; each for a range of *p* (0.1%, 0.5%, 1%, 2%, 5%, 10%, 20%, 30%, 40%, 50%) and *β* (1/10, 1/5, ¼, 1/3, ½, 1, 2,3,4,5,10) values. We repeated our simulations 10000 times at each case-control proportion-*p*-*β* combination, and demonstrated that our formulae accurately estimate the desired values (**Figure S3**).

Next, in order to test whether diagnostic contamination may account for the associations observed within our PheWAS, we calculated the expected impact of diagnostic contamination for two genes (*NCKIPSD*-Aorta; *SEMA3F*-Spinal Cord) with the largest effect sizes *β* observed in our S-PrediXcan analysis, under two scenarios:

1. Diagnostic contamination occurs among our cases only, assuming the normal population rate for AN: 0.9% among women; 0.3% among men.
2. Contamination occurs at significantly higher levels than might be expected in the population (1.2%, 3%, 6%), and all of these samples fall into our case category.

For each scenario, we repeated our calculation using 500 and 1000 cases, matched with 1000, 5000, and 30000 controls.

## RESULTS

### S-PrediXcan identifies gene-tissue associations with AN

We applied S-PrediXcan to PGC-AN GWAS summary statistics, and identified 47 genes across 12 loci, two loci on chromosomes 2 and 3 that reached experiment-wide significance (p<3.75 × 10^−8^; **Figure 1, Table 2, Table S4**), and another 10 loci that reached tissue-specific significance (Each tissue type has its own p value threshold of significance; **Figure 1, Table 2, Table S4**). These 12 loci encompass 218 gene-tissue associations (which may include the same gene associated across multiple tissues). The most significant peak on chromosome 3 includes 29 genes with significant associations with AN; this region of the genome overlaps with a GWAS peak for inflammatory bowel disease (IBD)(44), Crohn’s disease, and ulcerative colitis. Many of the AN-associated genes within this peak have been found to be associated with IBD. Genes included in our AN GReX gene set have been previously associated not only with IBD, but with other psychiatric traits such as schizophrenia (*ARNTL, CLIC1*), depression and/or depressive symptoms (*C3orf62, CCDC36, CLIC1, DAG1, GPX1, MST1R*), autism (*CLIC1*), and substance use (*RBM5, RBM6*), as well as anthropometric measures such as BMI, waist-hip ratio, and lean body mass (**Table S5**). Of these genes, *CLIC1* lies within the major histocompatibility complex (MHC), a large region on chromosome 6 that includes genes that code for components of the adaptive immune system (Tibial nerve*-CLIC1*, p=1.47 × 10^−6^). To our knowledge, this is the first genetic association for AN within the MHC region.

| Chromosome | Genes | Zscore | P-value | Top Finding (Gene) | Top Finding (Tissue) |
| --- | --- | --- | --- | --- | --- |
| 1 | *FBLIM1* | -4.851060 | 1.23e-06 | *FBLIM1* | Liver |
| 2 | *ASB3, CHAC2, GPR75* | -5.780228 | **7.46e-09** | *GPR75* | Whole_Blood |
| 3 | *SPINK8, PFKFB4, TMEM89, SLC26A6, CELSR3, NCKIPSD, ARIH2OS, ARIH2, P4HTM, WDR6, DALRD3, NDUFAF3, USP19, LAMB2, CCDC71, CCDC36, C3orf62, GPX1, NICN1, DAG1, APEH, MST1, MST1R, RNF123, RBM6, RBM5, SEMA3F, TUSC2* | 7.321127 | **2.46e-13** | *WDR6* | Skin_Sun_Exposed_Lower_leg |
| 3 | *PROS1, DHFRL1* | 4.687382 | 2.77e-06 | *DHFRL1* | DGN_Whole_Blood |
| 6 | *CLIC1* | 4.814629 | 1.47e-06 | *CLIC1* | Nerve_Tibial |
| 10 | *MGMT, EBF3* | -5.086391 | 3.65e-07 | *MGMT* | Thyroid |
| 11 | *ARNTL* | 4.534957 | 5.76e-06 | *ARNTL* | Whole_Blood |
| 12 | *SUOX, RPS26, SLC26A10* | 4.628307 | 3.69e-06 | *RPS26* | Uterus |
| 17 | *TNFSF12, LINC00324* | 4.982641 | 6.27e-07 | *TNFSF12* | Breast_Mammary_Tissue |
| 20 | *SLC2A10* | 4.921176 | 8.60e-07 | *SLC2A10* | Nerve_Tibial |
| 22 | *KREMEN1* | 4.872670 | 1.10e-06 | *KREMEN1* | Nerve_Tibial |
| 22 | *SCO2* | -4.560015 | 5.11e-06 | *SCO2* | Brain_Cerebellum |

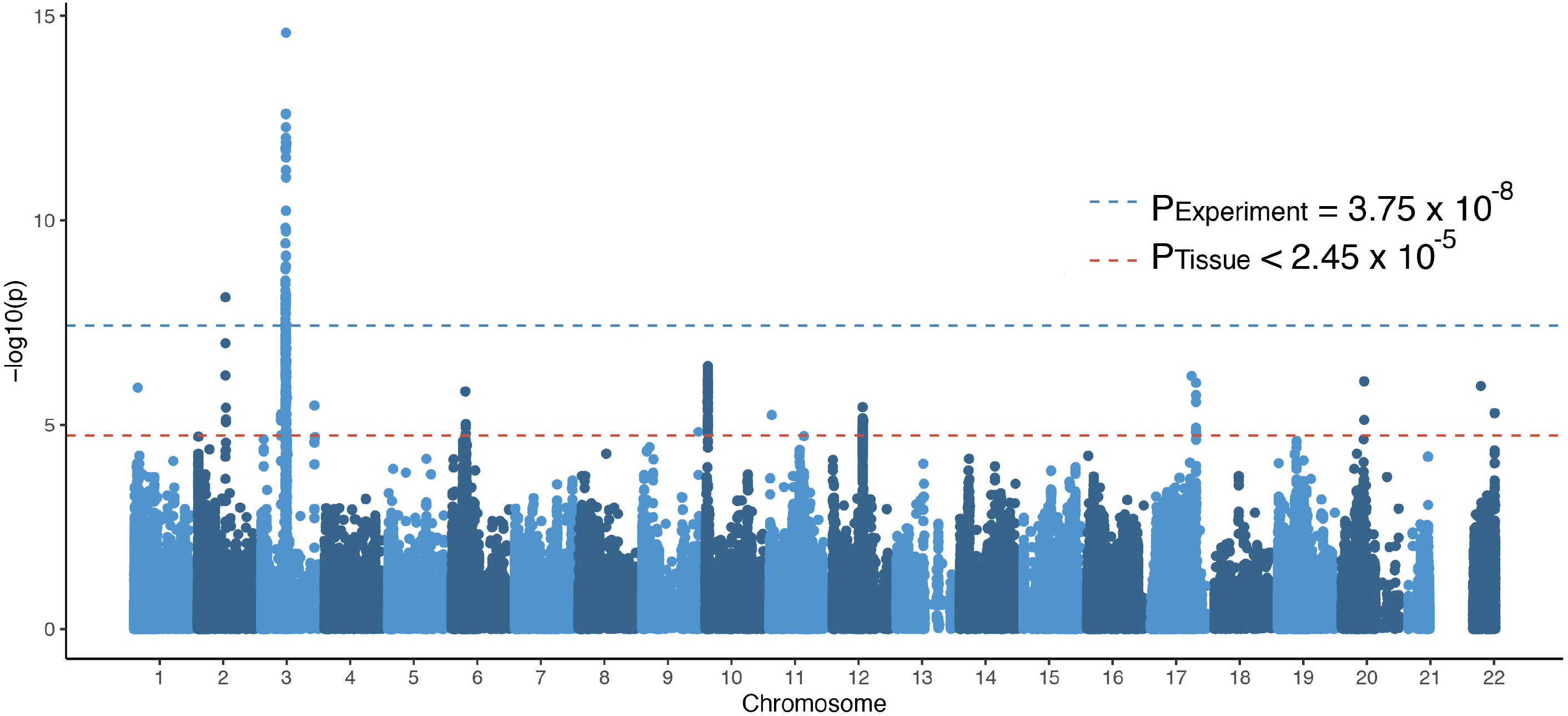

### AN-GREX is associated with anthropometric measures

We tested whether GReX of AN-associated genes was associated with anthropometric phenotypes in our Bio*Me*™ biobank (**Table 3**). In the overall cohort, upregulation of MHC-gene *CLIC1* was associated with lowest recorded weight (Spleen-*CLIC1*, p=6.63 × 10^−4^; Breast Mammary tissue-*CLIC1*, p=4.18 × 10^−4^) and mean height (Subcutaneous Adipose-*CLIC1*, p=1.39 × 10^−5^; **Table S6**). Downregulation of multiple genes, *ARIH2, NCKIPSD, NDUFAF3, NICN1, P4HTM, RNF123, SLC26A10*, was associated with decreased measures of weight change over time in the overall cohort (p<9.86 × 10^−4^)(**Table 3, Figure S4**). Upregulation of *WDR6* was associated with increased measures of weight change over time (p<9.81 × 10^−4^)(**Table 3, Figure S4**). *CTNNB1, GPR75* and *LINC00324* GReX was associated with mean height in Bio*Me*™ (p<6.51 × 10^−4^)(**Table S6**).

### Patient BMI mediates AN-GREX-PheWAS associations

Given the relationship of AN with BMI, we performed a secondary analysis to assess the effect of BMI on the association between AN-GReX and anthropometric phenotypes. Specifically we wanted to look at whether our AN-GReX-phenotype associations in individuals of High and Low BMI were still present when BMI was within a normal range. We split the Bio*Me*™ cohort into three groups: High, Mid, and Low BMI (adjusted for ancestry and sex; see **Methods** and **Supplementary Information** for more details), and re-ran our PheWAS within these groups. Indeed, our AN-GReX anthropometry associations (with highest recorded weight, lowest recorded weight, and weight change over time) varied according to the BMI group tested (**Figure 2, Table 3**). The association between *CLIC1*-GReX and highest and lowest recorded weight measures was strongest in individuals of High BMI compared with the other BMI groups (**Figure 2, Table 3**). Likewise, for multiple genes, associations between AN-GReX and weight change over time differed markedly in the High BMI group (**Table 3, Figure S5**).

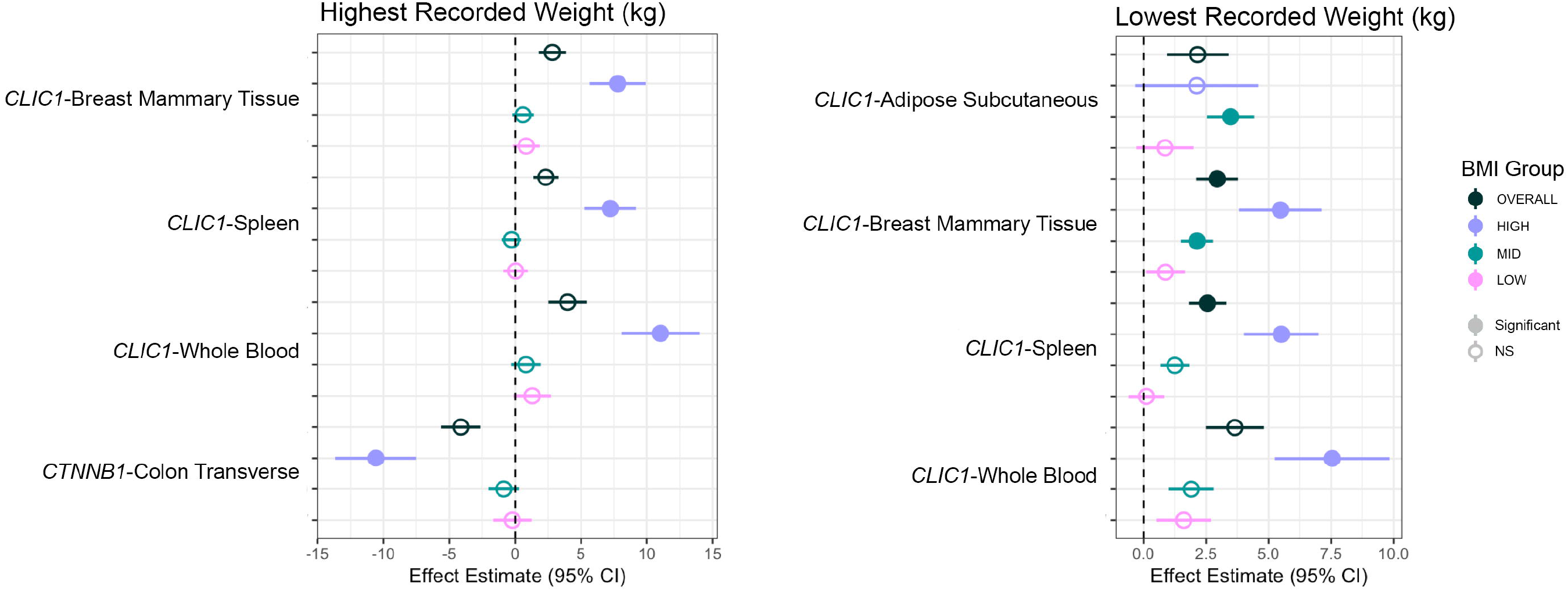

### AN-GREX is associated with autoimmune and autoinflammatory diseases

At a false discovery rate (FDR) p<0.05, we found 17 FDR-significant gene-tissue associations with four Phecode- and Encounter Diagnosis phenotypes: type I diabetes, celiac disease, peptic ulcer, and unspecified immunodeficiency (**Figure 3**). Upregulation of *CLIC1* in spleen was associated with two autoimmune/autoinflammatory phenotypes in the overall Bio*Me*™ cohort: celiac disease and type I diabetes (p=1.30 × 10^−11^, p=5.08 × 10^−20^, respectively) (**Tables S7 and S8**). Downregulation of *SLC26A6* was associated with the phenotype “peptic ulcer, site unspecified” in the overall cohort (Liver-*SLC26A6*, p=8.38 × 10^−8^; Skeletal muscle-*SLC26A6*, p=1.71 × 10^−7^; Thyroid-*SLC26A6*, p=7.16 × 10^−8^). Downregulation of *PFKFB4* was associated with ICD10 code D84.9, “immunodeficiency, unspecified” (Skin not sun exposed-*PFKFB4*, p=1.34 × 10^−7^). We stratified our cohort by BMI (see **Methods**) to assess whether BMI was modifying the association of AN-GReX with Encounter Diagnosis and phecodes. We saw significant associations of AN-GReX phecodes within our Mid BMI group; however, when plotted across the BMI groups, there was no notable difference in the magnitude and direction of effect of AN-GReX on either Encounter Diagnosis or phecodes across the BMI groups (**Figures S6 and S7**). Within individuals with Mid BMI, upregulation of *CLIC1* was similarly associated with type 1 diabetes (Mid-Subcutaneous adipose, Mid-Breast Mammary tissue, Mid-Spleen-*CLIC1*, p<1.96 × 10^−8^).

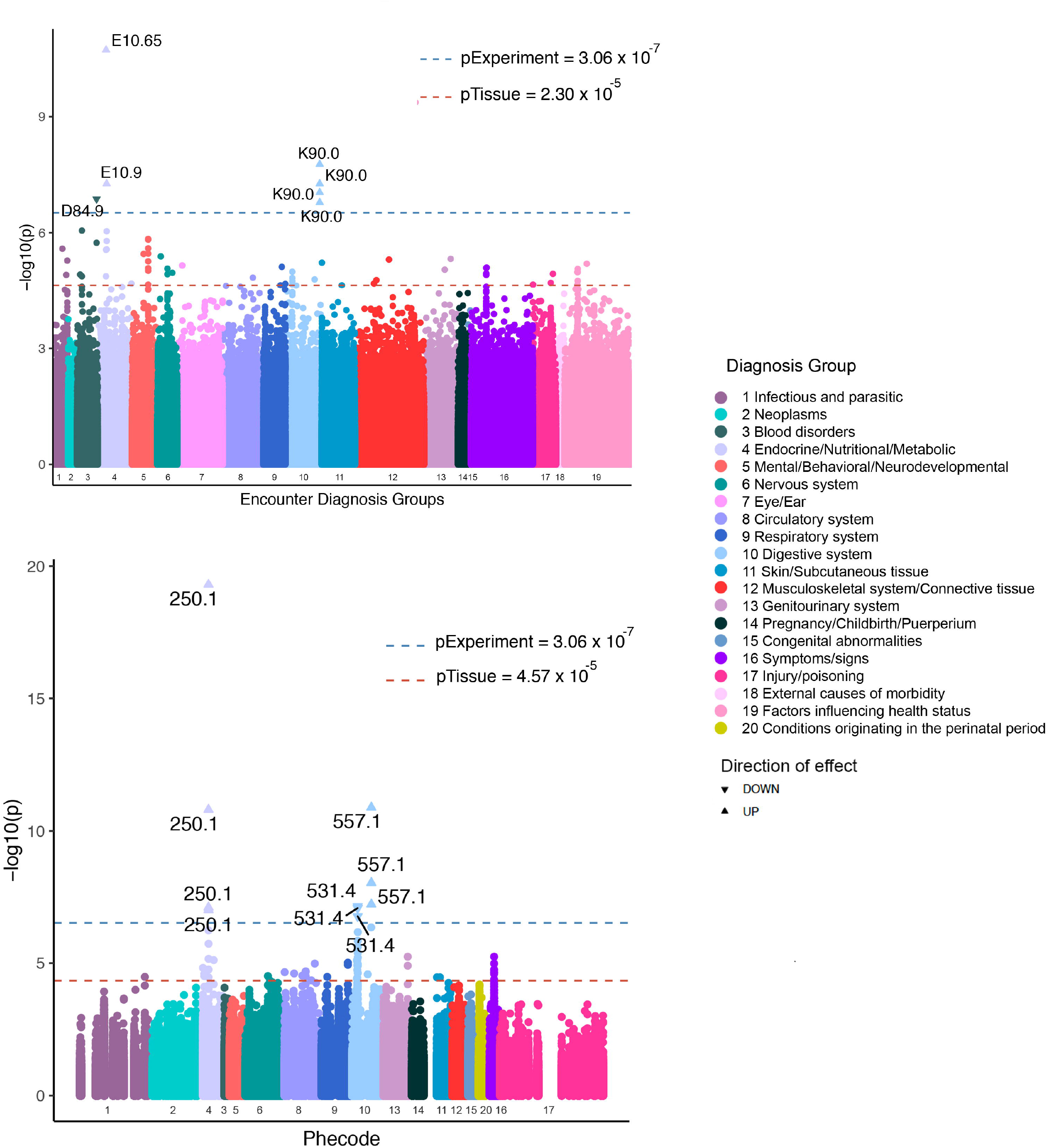

### AN-GREX is associated with tobacco, alcohol, and illicit drug use

Multiple AN genes were associated with both categorical and continuous measures of tobacco use (**Table S9, Figure S8**). *APEH, CCDC36, DALRD3, GPX1, LINC00324, NICN1*, and *P4HTM* were associated with years used/smoked of tobacco in the overall Bio*Me*™ cohort (p<4.97 × 10^−4^, **Table S9**) and upregulation of *MGMT* was associated with being a smoker (Heart Atrial Appendage-*MGMT*, p=8.87 × 10^− 5^; Putamen basal ganglia-*MGMT*, p=1.09 × 10^−4^). Again, we note mediating effects of patient BMI category on the association between AN-GReX and smoking phenotypes. Among the Mid and High BMI groups, different genes were associated with both continuous and categorical measures of tobacco use (**Table S9, Figure S8**), including years of tobacco use, tobacco cigarette packs per day, and smoking every day. In addition to tobacco use measures, we see an association between AN genes and tobacco use related ICD codes. Downregulation of *LINC00324* in Low BMI individuals was associated with ICD10 diagnosis code F17.210 “Nicotine dependence, cigarettes, uncomplicated” (Low-Cells EBV-tranformed-*LINC00324*, p=3.03 × 10^−5^).

Downregulation of *MGMT* was associated with decreased alcohol use (ounces per week) in individuals in the Mid-BMI group (p<9.75 × 10^−4^; **Figure S8**). In individuals of Low BMI, downregulation of *GPR75* and *GPX1*, and upregulation of *NICN1* were associated with decreased and increased continuous measures of illicit drug use frequency respectively (Liver-*GPR75*, p=3.34 × 10^−4^; Nucleus accumbens-*GPX1*, p=2.04 × 10^−4^; Testis-*NICN1*, p=3.34 × 10^−4^; **Figure S8**).

### AN-GREX is associated with measures of pain score and pain location

We tested for association between AN-GReX and three sets of pain score measures: (1) pain location, (2) pain score overall, (3) pain score by body location. AN-GReX was associated with multiple measures of pain score measurement and location across all BMI groups (**Table S10**). Most significantly, upregulation of *TNFSF12* was associated with total knee pain score (sum) in individuals with High BMI (High-Brain Substantia nigra-*TNFSF12*, p=1.50 × 10^−3211^). Downregulation of *TNFSF12* was associated with total abdomen pain score (sum) in individuals with Low BMI (Low-Brain Substantia nigra-*TNFSF12*, p=3.93 × 10^−58^) (**Table S10**).

When looking at pain location irrespective of pain score, upregulation of *TUSC2* in the overall cohort is associated with presence of knee pain (Pancreas-*TUSC2*, p=7.62 × 10^−6^) and downregulation of *CTNNB1* in Mid BMI individuals is associated with the presence of left leg pain (Mid-Stomach-*CTNNB1*, p=1.93 × 10^−5^) (**Figure 4, Table S10**). Repeating this analysis according to BMI category reveals a very large number of associations specific to the Low BMI category (**Figure 4, Table S10**); in particular, we note 129 gene-tissue associations with presence of foot pain, nine of which pass phenotype-wide significance, which may indicate a propensity to excess exercise, or exercise-related injuries among these individuals.

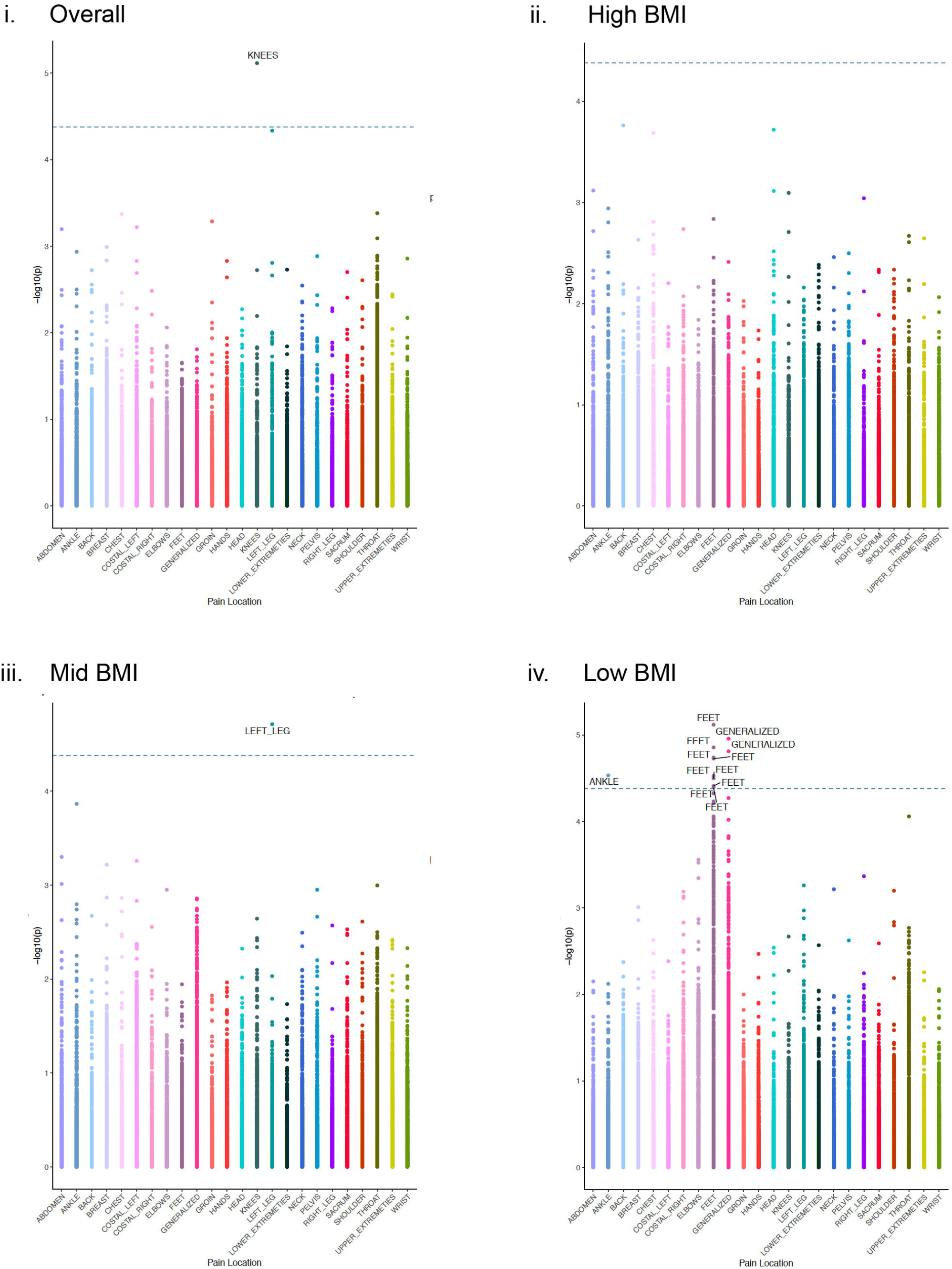

### Upregulation of CLIC1 is associated with glucagon medication

AN-GReX PheWAS with medication phenotypes was associated with glucagon, a hormone used to treat severe hypoglycemia, in the overall Bio*Me*™ cohort. Upregulation of *CLIC1* was associated with glucagon in the overall cohort (Spleen-*CLIC1*, p=4.20 × 10^−9^) and also in individuals with Mid BMI (Mid-Spleen-*CLIC1*, p=2.09 × 10^−6^) (**Figure S9, Table S11**).

### Identifying potential case contamination effects

Our Bio*Me*™ patients have not all been explicitly assessed for eating disorders (ED), and information regarding ED diagnoses and assessments earlier in life may be omitted from the records due to incomplete clinical history assessments. Therefore, it is possible that diagnostic contamination within some of our sample is responsible for the associations observed within our data. In order to test this, we performed a simple experiment.

For this simple experiment, we assume that diagnostic contamination occurs among true cases only, at prevalence *p*. In this case, we expect gene expression among those individuals to be up-or down-regulated by effect size beta; consequently, we expect a gene expression difference between cases and controls proportional to the prevalence of the contamination within the sample (*p*) and the effect size (beta); (**Supplementary information**). We tested whether such contamination may drive the associations observed in our study, assuming the following different contamination scenarios (**Table S12**), for two genes with large S-PrediXcan effect sizes: (i) that contamination occurs among cases, assuming the highest common estimate for AN prevalence (0.9% among women, 0.3% among men); (ii) that contamination occurs within our biobank, assuming the highest common estimate for AN prevalence (0.6%, 190 cases), and that all of these samples fall into our ‘case’ category.

Our model indicates that diagnostic contamination among cases is unlikely to result in significant PheWAS associations in our data. Contamination occurring at population ED prevalence (0.6%) did not result in significant associations for any of the case-control scenarios in our model (**Table S12**); nor did contamination at 1.2% or 3%. Assuming 6% contamination resulted in potentially significant associations (estimated p<2.8×10^−8^; see **Methods** and **Supplementary Methods** for more detail).

## DISCUSSION

### AN-associated genes are enriched for associations with psychiatric, autoimmune, and anthropometric phenotypes

Our S-PrediXcan gene results illustrate the contribution of genes associated with metabolic, anthropometric, autoimmune, and psychiatric phenotypes to AN, and 53% of our genes (25/47) overlap with those from a previous prediXcan analysis of AN cohorts(9). 24/47 AN genes (51%) had variants that were previously associated with some autoimmune disorder in GWAS studies (**Table S5)**. The experiment-wide significant loci on chromosome 3 overlaps with a known GWAS peak for inflammatory bowel disease(44) (IBD), and many of the genes within that peak that we found associated with AN were also associated with disorders under the IBD umbrella such as Crohn’s and ulcerative colitis (**Table S5**), demonstrating further shared genetic architecture of AN with autoimmune and autoinflammatory disease(7,8,45,46). Earlier AN GWAS studies have found AN risk loci that overlap with loci implicated in autoimmune disease(47), but the association of *CLIC1* gene expression in tibial nerve that we have found is the first known association of AN with the MHC locus, a region that has been associated with many other psychiatric disorders(48,49) as well as autoimmune disorders. In particular, *CLIC1* has previously been identified as associated with schizophrenia(50), autism(50), major depressive disorder(51), post-traumatic stress disorder(52), neuroticism(53) and depressive phenotypes(53). Importantly, *CLIC1* variants have also been associated with complement component C4 and C3 protein levels in the blood(54). *C4* is a gene with two isotypes, *C4A* and *C4B*, that, through various structural allelic combinations, give rise to complement component 4, which, along with complement component 3 (C3), is part of the complement cascade of proteins(55). Previous research has shown that the complement cascade is involved in not only immunological functions of pathogen clearance (e.g., tagging pathogens for destruction), but also in synaptic pruning and neuronal connectivity(56). The *C4A* isotype has been associated with schizophrenia risk(55), and both *C4A* and *C4B* genes had subthreshold associations with AN in our dataset.

*CLIC1* (Chloride Intracellular 1) encodes a chloride ion channel protein involved in many necessary cellular functions including the regulation of cell membrane potential, and the proliferation and differentiation of cells(57), including a role in axonal outgrowth of neurons^51^. In addition to neurons, *CLIC1* has been found to be expressed in many different tissue types(59), and in particular, CLIC1 protein has been found to accumulate in peripheral blood mononuclear cells (PBMCs) when the central nervous system is in a state of chronic inflammation(58), and increased CLIC1 expression has been shown to occur in microglia stimulated by the presence of β-amyloid *in vitro*(60,61). Therefore, CLIC1 may potentially act as a biomarker for neurodegenerative disorders such as Alzheimer’s disease, and downregulation of CLIC1 has been shown to have a neuroprotective effect in the presence of β-amyloid proteins(58,60–62). Variants in *CLIC1* may contribute to risk of multiple psychiatric disorders and behaviors, such as schizophrenia, MDD and PTSD, as well as many autoimmune and immune-related phenotypes such as asthma and systemic lupus erythematosus(50–53),(63) (**Table S5**). Many studies have posed the role of inflammation in the development of psychiatric disorders, and in the etiology of AN(7,8,45,46,64).

Multiple genes (8/47 or 17%) in our results were associated with regular gym attendance (**Table S5**), or measures of physical activity. Excessive and compulsive exercise is a behavior often seen in individuals with AN(65,66), and evidence is suggestive that hyperactivity increases risk of chronic AN(67,68). Likewise, genetic studies have shown a strong genetic correlation between AN and physical activity (r_g_=0.17)(9,23,69). These genes may reflect a genetic liability to compulsive behaviors of physical activity. Our pain location results point to possible symptoms of excessive activity, with the strong association of AN-GReX with foot pain in Low BMI individuals. Given the known association of AN disease with low bone mineral density (BMD) and propensity for bone fracture(70), our results may reflect the result of genes associated with compulsive exercise and BMD contributing to increased osteoarticular pain. We also see multiple genes that have been previously associated with decreased sleep and insomnia phenotypes (including a core circadian clock gene *ARNTL*), which have previously been implicated in a range of psychiatric disorders(71–82), including eating disorders(83), as well as in satiety and hunger(84,85).

### In a patient population, expression of AN genes is associated with psychiatric, metabolic, and autoimmune phenotypes

Unlike GWAS, which include carefully constructed case-control cohorts, PheWAS encompass all individuals within a given healthcare system, including patients with subthreshold or partial presentations of a disorder, or individuals with commonly comorbid or co-diagnosed conditions. Importantly, because individuals enrolled in the biobank are not ascertained for any specific disorder, they likely represent a comprehensive clinical picture of the comorbidities and symptomatology associated with AN gene expression. Examining the consequences of aberrant predicted gene expression among these patients (i.e., testing for GReX-associations) may reveal clinical and biological consequences of these genes; for example, studying whether AN-associated genes have anthropometric and metabolic consequences among adults with no evidence for previous AN-or ED-diagnoses may disentangle whether certain endophenotypes present as a cause, or consequence of AN. For example, it has been postulated that behaviors of food avoidance and restriction may arise due to gastrointestinal complaints and distress that provoke these behaviors and precede development of AN(86). Likewise, autoimmune disorders of the gastrointestinal tract, such as celiac disease and Crohn’s disease, show a bidirectional relationship with AN, with previous diagnosis of a GI-associated autoimmune disorders increasing the risk of AN and vice versa(8). Our pheWAS results of AN-GReX associations with gastrointestinal symptoms such as abdominal pain, ascites, and peptic ulcer, as well as GI-related autoimmune disorders like celiac disease, suggest AN-GReX may contribute directly to these diseases and phenotypes, and that food aversive behaviors and gastric distress may be genetically regulated in these individuals, rather than occurring as a consequence of AN. While these associations could be the consequence of undiagnosed AN individuals in our biobank, they more likely reflect real biological associations of expression of those particular genes with the phenotype.

Our results further confirm the contribution of metabolic factors to AN etiology as we see a very robust association of AN-GReX with type 1 diabetes, and our top findings for AN-GReX with medication prescriptions include the hyperglycemic hormone glucagon and various forms of insulin. The association of AN with metabolic traits and abnormalities has been fairly well established. The Anorexia Nervosa Genetics Initiative reported significant negative genetic correlation between AN and a number of metabolic traits including insulin resistance (r_g_=-0.29), fasting insulin (r_g_=-0.24), leptin (r_g_=-0.26) and type 2 diabetes (r_g_=-0.22), and a strong positive genetic correlation with HDL cholesterol (r_g_=0.21)(9). Additional studies have shown associations of AN with fasting insulin(87) and insulin sensitivity(88). Notably, our results point to a similar role of aberrant glycemic regulation in the etiology of AN. Future analyses including EHR-derived lab results (LabWAS) studies may further elucidate AN genes associated with abnormal metabolic regulation and clinical features.

### BMI modification of AN-GReX on clinical outcomes

We stratified our pheWAS analyses by BMI in order to observe whether the effects of predicted AN gene expression on clinical outcomes were modified by BMI status, and whether this modification had a significant impact on clinical phenotypes. Specifically, are the AN-GReX-phenotype associations that we see in individuals of High and Low BMI still present when BMI is within the normal “healthy” range? Understanding how BMI is influencing the association of AN-GReX with the clinical phenome might give us further insight into the biological pathways leading to those outcomes and the conferral of AN disease risk. Additionally, we want to understand how BMI could be interacting with AN genetic risk to influence clinical outcomes. We know from our S-PrediXcan analyses and previous GWAS that many genes found to be associated with AN have variants that have been associated with BMI and other anthropometric traits (**Table S5**). 20/47 AN-genes (42.6%) had evidence from previous GWAS of association with anthropometric measures such as BMI, body fat distribution, waist-to-hip ratio, fat-free mass, obesity, and hip circumference. BMI polygenic risk scores (PRS) have been shown to be associated with disordered eating, with high BMI PRS negatively associated with thin ideal internalization and external eating, where individuals tend to eat in response to external cues of the sight or smell of food(89,90).

We postulate that the modification of AN-GReX-weight associations by BMI could be due to actual biological differences in the regulation of AN gene expression and downstream processes in the presence of different measures of BMI. CLIC1 is involved in pathways of inflammation, and individuals with a higher BMI have been shown to have increased systemic inflammation(91), especially in adipose tissues, so it is possible that the modification effect by BMI on the effect of *CLIC1*-GReX on weight measures is happening via a tissue-specific inflammatory process. If we view AN and obesity as metabolic disorders on opposite ends of the spectrum, then perhaps the interaction of BMI with risk for either disorder may lead to differences in metabolic regulation resulting in higher weight phenotypes for those with increased obesity risk, and lower weight phenotypes in those with higher AN risk.

Although the hypothesis-free, phenome-wide design of our study allows for powerful detection of clinical and biological consequences of AN risk genes, the same design also bears some notable limitations and caveats. One key caveat to our results is the lack of diagnostic detail and insights available for the patients in our study. EHR analyses leverage large, existing data sets to rapidly amass cohorts for analysis, and to yield insights into whole phenome consequences of genotype and GReX associations; however, the scale and scope of these studies precludes deep phenotyping or performing clinical interviews. This lack of diagnostic precision may arise from a number of factors.

First, we make use of ICD codes within the medical record in order to infer diagnoses; since these are primarily used by clinicians for billing purposes, they likely provide an imperfect proxy for true disease state. In order to mitigate spurious results stemming from imperfectly assigned codes, we established criteria for inclusion of diagnoses in our analyses: First, diagnosis codes with fewer than 10 counts in the biobank were removed. Second, we only included diagnoses in our pheWAS with at least 10 counts of a code in the ancestral population (i.e., ancestral groups with fewer than 10 case counts were not included in the analysis for that diagnosis), and with a total effective sample size after meta-analysis that was greater than 100. Lastly, within each pheWAS, individuals had to have two or more counts of a diagnosis code to count as a “case”, whereas individuals with only one count of a diagnosis code were set to “missing” and not included in the analysis.

Second, it is possible that our patients regularly receive treatment at multiple different hospital systems; as such, we may be capturing only partial data for each of our patients. In order to mitigate this, we restricted our analysis to patients with multiple data points within our EHR. Future analyses that seek to harmonize or meta-analyze patient data across EHR (e.g., NYC-CDRN, PsycheMERGE) are ongoing, and will be vital to disentangling this effect further.

Importantly, our patients have not all been explicitly assessed for EDs, and information regarding ED diagnoses and assessments earlier in life may be omitted from the records due to incomplete clinical history assessments. Therefore, it is possible that case-contamination within some of our sample is responsible for the associations observed within our data. In order to address this, we performed a simple experiment to simulate the effect sizes expected if undiagnosed AN contamination drives our result. We estimated the expected association statistics observed due to two possible levels of contamination among PheWAS cases; i, that 0.06% of PheWAS-trait cases are missing a true AN diagnosis-i.e., the proportion of individuals we expect to receive an AN diagnosis based on standard prevalence estimates; ii, that 1.2%, 3% and 6% of individuals in BioMe are missing an AN diagnosis (i.e., double, five times, and ten times the expected population rate), and that all of these individuals fall within our PheWAS trait case group, and applied this model for two genes with large S-PrediXcan effect sizes (*NCKIPSD*-Aorta; *SEMA3F*-Spinal Cord). Our model indicates that diagnostic contamination at 0.06%, 1.2% or 3% is insufficient to account for the effect sizes observed within our PheWAS analysis. Moreover, since these genes are selected due to their large S-PrediXcan effect sizes, we expect that the contamination effects observed for these two genes will be greater than any others in our study; as such, we do not believe that our findings are attributable to hidden case contamination. Contamination at 6% resulted in significant differences; however, since this estimate represents a >five-fold increase in expected numbers of AN within a population, we do not expect that this level of contamination occurs in our sample.

Our results demonstrate that there are real clinical consequences to differences in AN gene expression. Characterization of the phenotypic consequences of AN gene expression in a clinical setting can give us more insight into the biological mechanisms underlying AN and, consequently, how to diagnose and treat the disorder. By understanding the associations of AN gene expression with symptomatology, prodromal or subthreshold disease states, we may gain insights into the biology of the disease, and perhaps identify therapeutic targets and opportunities for clinical interventions. For example, if gastrointestinal complaints are truly the consequence of aberrant AN gene expression, and contribute to disordered eating due to gastrointestinal distress, treatment of those symptoms may help alleviate other AN symptoms or prevent development of later AN(92,93). An understanding of the clinical consequences of AN gene expression can further augment the definition of AN, and could allow clinicians to more broadly identify individuals at greater risk of AN, or those who present with symptom constellations that do not yet meet the established diagnostic threshold.

## Supporting information

Supplemental Author List

## Data Availability

Summary data is made available in the supplementary tables of this article.

## Acknowledgements

We are deeply grateful for the mentorship of Pamela Sklar, whose guidance and wisdom we miss daily. We strive to continue her legacy of thoughtful, innovative, and collaborative science.

JJ and LMH were supported by funding from the Klarman Family Foundation (2019 Eating Disorders Research Grants Program) and the NIMH (R01MH118278). CMB is supported by NIMH (R01MH120170; R01MH119084; R01MH118278; U01 MH109528); Brain and Behavior Research Foundation Distinguished Investigator Grant; Swedish Research Council (Vetenskapsrådet, award: 538-2013-8864); Lundbeck Foundation (Grant no. R276-2018-4581).

This work was supported in part through the resources and staff expertise provided by the Charles Bronfman Institute for Personalized Medicine and The Bio*Me*™ Biobank Program at the Icahn School of Medicine at Mount Sinai.

Research reported in this paper was supported by the Office of Research Infrastructure of the National Institutes of Health under award numbers S10OD018522 and S10OD026880. The content is solely the responsibility of the authors and does not necessarily represent the official views of the National Institutes of Health.

## Competing Interests

CMB reports: Shire (grant recipient, Scientific Advisory Board member); Idorsia (consultant); Pearson (author, royalty recipient). ML declares that over the past 36 months, he has received lecture honoraria from Lundbeck pharmaceutical (No other equity ownership, profit-sharing agreements, royalties, or patent). The remaining authors declare no competing interests.

## Notes

### Author Declarations

Founded in September 2007, BioMe(TM) is a biobank that links genetic and electronic medical record (EMR) data for over 30,000 individuals recruited primarily in ambulatory care settings in the Mount Sinai Health System (MSHS) in New York City. The current study was approved by the Icahn School of Medicine at Mount Sinai Institutional Review Board (Institutional Review Board 07-0529). All study participants provided written informed consent.

### Summary of Updates

An analysis to test for hidden case contamination has been added to the main manuscript and supplemental text. Supplemental files have been updated to reflect this additional analysis.

