## Supplementary material for "The Phenome-wide Consequences of Anorexia Nervosa Genes": Table and Figure Legends

MAIN FIGURES AND TABLES

Figure 1: S-PrediXcan results for PGC-ED AN GWAS (N_Cases_=16,992, N_Controls_=55,525). S-PrediXcan transcriptomic imputation of the PGC-ED AN GWAS summary statistics to determine genetically-regulated gene expression (GReX), with tests for association of GReX with AN disease status. Manhattan plot of S-PrediXcan gene-tissue associations with anorexia nervosa for 50 tissues. Each point represents a different gene-tissue association result; i.e., the same gene may have multiple points within a peak.

**Figure 2: Significant AN-GReX-tissue associations with weight phenotypes**. Effect estimates and 95% CI for associations of AN*-*GReX with (i) highest recorded weight (kg) and (ii) lowest recorded weight (kg). Associations are shown for four BMI groups: Overall, High, Mid and Low BMI. Tissue-specific significance threshold set at p<0.001.

Figure 3: AN-GReX associations with full Bio*Me*™ Encounter Diagnosis ICD codes and Phecodes. Diagnosis codes are plotted along the X-axis and grouped by category. (A) Manhattan PheWAS plot of AN-GReX gene associations with Encounter Diagnosis ICD codes (N=2,178) (B) Manhattan PheWAS plot of AN-GReX gene associations with Phecodes (N=1,093). Encounter Diagnosis ICD-10 code descriptions: D84.9 – Immunodeficiency, unspecified; E10.65 – Type 1 diabetes mellitus with hyperglycemia; E10.9 – Type 1 diabetes mellitus without complications; K90.0 – Celiac disease. Phecode descriptions: 250.1 – Type 1 diabetes; 531.4 – Peptic ulcer, site unspecified; 557.1 – Celiac disease.

Figure 4: AN-GReX associations with pain location. Pain locations are shown along the x-axis (N=25). Associations are shown for four BMI groups (A) Overall cohort (B) High BMI (C) Mid BMI and (D) Low BMI. Bonferroni significance threshold is demarcated by the dotted line (p=4.17 x 10^-5^).

Table 1: Bio*Me*™ Demographics (N=31,585). Ancestry designations: AA (African), EA (European), ESA (East Asian), HA (Hispanic), SAS (South Asian), NA (Native American).

Table 2: S-PrediXcan loci results. Summary of S-PrediXcan of PGC-ED AN GWAS results. Genes associated with Anorexia nervosa from each locus are shown, as well as the top finding Zscore, p-value, gene and tissue for each locus. P-values shaded in bold indicate experiment-wide significant loci (p<3.75 x 10^-8^).

**Table 3: PheWAS AN-GReX tissue associations with weight phenotypes**. AN-GReX association with phenotypes of (i) Highest recorded weight (kg), (ii) Lowest recorded weight (kg), (iii) weight change over time (kg/yr) for four BMI groups: Overall, High, Mid and Low BMI. Tissue-specific significance threshold set at p<0.001.

**SUPPLEMENTARY FIGURES AND TABLES**

Figure S1: BMI stratification of the Bio*Me*™ cohort. We stratified the Bio*Me*™ individuals into three BMI groups: High, Mid and Low, based on the distribution of BMI within each ancestry by sex. Individuals whose BMI fell above the 3^rd^ quartile of BMI distribution were assigned to the High BMI group, those whose BMI fell below the 1^st^ quartile were assigned to Low, and those between the 1^st^ and 3^rd^ quartiles of the distribution were assigned to the Mid BMI group.

Figure S2: Distribution of BMI by Bio*Me*™ ancestry and sex. BMI distributions are plotted by sex (female, male) for each ancestry group. Ancestry is coded as: AA (African), EA (European), ESA (East Asian), HA (Hispanic), NA (Native American), SAS (South Asian).

Figure S3: Distribution of simulated gene expression to test for case contamination. Simulated gene expression distributions for 10,000 permutations at varying levels of contamination rate (*p*) and effect on GReX ($\boldsymbol{\beta}$) for 1000 cases (X) and 1000 controls (Y). (A) p=0.01, $\boldsymbol{\beta}$=0.1 (B) p=0.1, $\boldsymbol{\beta}$=0.5 (C) p=0.001, $\boldsymbol{\beta}$=1 (D) p=0.4, $\boldsymbol{\beta}$=0.25.

Figure S4: Forest plot of Overall weight change over time associations with AN-GReX. Effect estimate and 95% CI measures for AN-GReX (Gene-Tissue) associated with the measurement of weight change over time (measured as kg/year) in the overall Bio*Me*™ cohort. Tissue-specific significance threshold set at p<0.001.

**Figure S5: Forest plot of Overall weight change over time associations with AN-GReX across three BMI groups**. Effect estimate and 95% CI measures for AN-GReX (Gene-Tissue) associated with the measurement of weight change over time (measured as kg/year) in four BMI groups: Overall, High BMI, Mid BMI and Low BMI. Tissue-specific significance threshold set at p<0.001.

Figure S6: Forest plot of Encounter Diagnosis associations across BMI groups. Effect estimates and 95% CI for the associations of AN-GReX with Encounter Diagnosis ICD-10 codes across the four BMI groups: Overall, High BMI, Mid BMI and Low BMI. Diagnosis code and description, as well as Gene-Tissue of association, given on the Y-axis. Experiment-wide significance threshold set at p<3.06 x 10^-7^.

Figure S7: Forest plot of Phecode associations across BMI groups. Effect estimates and 95% CI for the associations of AN-GReX with phecodes across the four BMI groups: Overall, High BMI, Mid BMI and Low BMI. Phecode and description, as well as Gene-Tissue of association, given on the Y-axis. Experiment-wide significance threshold set at p<3.06 x 10^-7^.

**Figure S8: Forest plot of Substance Use AN-GReX associations across BMI groups**. Effect estimates and 95% CI for the associations of AN-GReX with substance use phenotypes across the four BMI groups: Overall, High BMI, Mid BMI and Low BMI. Tissue-specific significance threshold set at p<0.001.

Figure S9: Forest plot of *CLIC1*-Spleen association with Glucagon medication across BMI groups. Effect estimates and 95% CI for the associations of *CLIC1*-Spleen GReX with Glucagon medication across the four BMI groups: Overall, High BMI, Mid BMI and Low BMI. Filled circles indicate Bonferroni-significant associations. Tissue-specific significance threshold set at p<5.26 x 10^-5^.

Figure S10: Graphical depiction of S-PrediXcan and PrediXcan transcriptomic imputation and pheWAS analyses. We used S-PrediXcan predictor models for 50 different tissue types to impute genetically-regulated gene expression (GReX) in the Watson, *et. al*, 2019 AN GWAS and found 47 genes whose GReX was associated with Anorexia Nervosa. We then imputed GReX for those 47 AN genes in our Bio*Me*™ cohort and performed a phenome-wide association study (pheWAS) across available EHR phenotypes. The pheWAS analyses were run within each ancestral population and then meta-analyzed using an inverse-variance approach in METAL. Secondary analyses included stratifying individuals in Bio*Me* by BMI and running the pheWAS analyses within each BMI group.

Figure S11: Descriptive statistics for continuous measures of substance use. Continuous phenotypes for alcohol, illicit drug and tobacco use. Alcohol use was measured as number of ounces of alcohol consumed per week, Illicit drug use was measured as number of times used (frequency), and tobacco use was measured as number of years usage of tobacco and number of packs of cigarettes smoked per day.

Figure S12: Descriptive statistics for continuous OBGYN outcome phenotypes. Counts for the presence of nine OBGYN phenotypes: (1) Current pregnancy (2) Parity (number of pregnancies with outcome of live birth) (3) Gravida (number of pregnancies regardless of birth outcome) (4) Births to term (5) Preterm births (6) Ectopic pregnancies (7) Abortion (8) Spontaneous abortion (miscarriage) (9) Therapeutic abortion.

Figure S13: Descriptive statistics for vital sign measurement phenotypes. Vital sign measurements for blood pressure (systolic, diastolic and pulse pressure), height (cm), pulse (beats per minute), pulse oximetry (%), respirations (breaths per minute) and temperature (degrees Celsius).

Figure S14: Descriptive statistics of weight phenotypes. Distribution of weight measurements (in kg) of highest recorded and lowest recorded weight in Bio*Me*™. Weight change over time was measured as average kg change per year.

Figure S15: Descriptive statistics for pain score and pain location phenotypes. (A) Histograms of pain score (regardless of pain location) recorded on a 0 to 10 scale; measurements used for pheWAS analyses included highest recorded pain score, mean pain score and sum of all pain score measurements. Mean pain score is indicated by the dotted line. (B) Violin plots of pain score by location measured as highest recorded pain score, mean pain score, and sum of pain scores for each pain location. Mean pain score measurements for each phenotype is indicated by the red dot.

Figure S16: Manhattan plot of association of Personal History phenotypes with AN-GReX for four BMI groups. Personal History phenotypes (N=35; X-axis) and association with AN-GRex (-log10(p) on Y-axis) for four BMI groups: Overall, High BMI, Mid BMI, and Low BMI. Experiment-wide p=2.86 x 10^-5^; Tissue-specific p=1.43 x 10^-3^.

**Figure S17: Regression plots for individual *CLIC1*-GReX versus lowest and highest weight measures**. Linear regression model lines with 95% confidence intervals for individual CLIC1-GReX measures for (A) Subcutaneous Adipose, (B) Breast Mammary Tissue, (C) Spleen, and (D) Whole Blood tissues for lowest recorded weight and highest recorded weight measures (kg). Forest plots of effect estimates for associations of *CLIC1*-GReX with lowest and highest recorded weight.

Figure S18: *CLIC1*-GReX versus lowest recorded weight measurement (kg). (A) Individual *CLIC1*-GReX measures versus lowest recorded weight measurements for breast and spleen tissues with regression line and 95% confidence interval. (B) Individual *CLIC1*-GReX measures versus lowest recorded weight measurements (kg) for Breast Mammary tissue and Spleen, with regression line only and 95% confidence interval.

**Figure S19: Distribution of *CLIC1*-GReX by BMI group**. Density plots of *CLIC1*-GReX for (A) Breast Mammary, (B) Spleen, (C) Subcutaneous Adipose, and (D) Whole Blood tissues by three BMI groups, High, Mid and Low. P value for KS test for significance shown for each tissue.

**Table S1: Bonferroni-corrected significance thresholds for each tissue model tested for S-PrediXcan**. Indicates number of genes tested per tissue, with p value for tissue-specific significance set at p=0.05/N_Genes_.

Table S2: BMI stratification values by Bio*Me*™ ancestry. BMI values used for determining assignment to High, Mid or Low BMI groups for each ancestral population and sex. High BMI range indicates the values above the 3^rd^ quartile. Low BMI indicates the BMI values below the 1^st^ quartile, and Mid shows the BMI range between the 1^st^ and 3^rd^ quartiles of BMI distribution.

Table S3: Sample numbers for each BMI group. Sample numbers indicated for each BMI group after stratification. Ancestry is coded as: AA (African), EA (European), ESA (East Asian), HA (Hispanic), NA (Native American), SAS (South Asian).

Table S4: Full summary statistics for S-PrediXcan transcriptomic imputation of the PGC-AN GWAS^9^. Gene-level summary statistics for transcriptomic imputation of AN-GWAS (N_Cases_=16,992, N_Controls_=55,525) using S-PrediXcan models for CMC DLPFC, DGN Whole Blood and 47 GTEx v7 tissues.

**Table S5: AN S-PrediXcan gene summary**. Summary description of 47 AN genes, including information on previous GWAS trait associations.

**Table S6: Vital sign pheWAS results**. GReX-Tissue-Phenotype associations for Vital Sign phenotypes (Blood pressure, Height, Pulse, Pulse Oximetry, Respirations, Temperature and Weight) across all BMI groups: Overall, High, Mid, and Low. Tissue-specific threshold set at p<0.001.

**Table S7: Encounter Diagnosis pheWAS results**. GReX-Tissue-Phenotype associations for encounter diagnosis phenotypes (N=2,178) across all BMI groups: Overall, High, Mid, and Low. Experiment-wide p value threshold p= 3.06 x 10^-7^; Tissue-specific threshold p= 2.30 x 10^-5^.

**Table S8: Phecode pheWAS results**. GReX-Tissue-Phenotype associations for phecode phenotypes (N=1,093) across all BMI groups: Overall, High, Mid, and Low. Experiment-wide p value threshold p = 3.06 x 10^-7^; Tissue-specific threshold p= 4.57 x 10^-5^.

**Table S9: Social History pheWAS results**. GReX-Tissue-Phenotype associations for Social History phenotypes (Alcohol, Illicit Drug and Tobacco use; Sexual History) across all BMI groups: Overall, High, Mid, and Low. Tissue-specific threshold set at p<0.001.

**Table S10: Pain Score and Location pheWAS results**. GReX-Tissue-Phenotype associations for Pain score and location phenotypes across all BMI groups: Overall, High, Mid, and Low. Tissue-specific threshold set at p<0.002.

**Table S11: Medication pheWAS results**. GReX-Tissue-Phenotype associations for medication phenotypes (N=951) across all BMI groups: Overall, High, Mid, and Low. Tissue-specific threshold set at p<5.26 x 10^-5^.

**Table S12: Expected association levels due to diagnostic contamination (ie, PheWAS trait cases with missing, undeclared or unknown AN diagnoses).**

**Table S13: Allergy pheWAS results**. GReX-Tissue-Phenotype associations for allergy phenotypes (N=113) across all BMI groups: Overall, High, Mid, and Low. Tissue-specific threshold set at p<4.42 x 10^-4^.

**Table S14: Encounter orders pheWAS results**. GReX-Tissue-Phenotype associations for Encounter order phenotypes (N=1,609) across all BMI groups: Overall, High, Mid, and Low. Tissue-specific threshold set at p<3.11 x 10^-5^.

**Table S15: Family History pheWAS results**. GReX-Tissue-Phenotype associations for Family history phenotypes (N=144) across all BMI groups: Overall, High, Mid, and Low. Tissue-specific threshold set at p<3.47 x 10^-4^.

**Table S16: Personal History pheWAS results**. GReX-Tissue-Phenotype associations for Personal history phenotypes (N=35) across all BMI groups: Overall, High, Mid, and Low. Tissue-specific threshold set at p<0.001.

**Table S17: OBGYN pheWAS results**. GReX-Tissue-Phenotype associations for OBGYN phenotypes (N=9) across all BMI groups: Overall, High, Mid, and Low. Tissue-specific threshold set at p<0.006.

**Table S18: Full pheWAS summary results**. GReX-Tissue-Phenotype associations for all pheWAS phenotypes (N=6,000+) across all BMI groups: Overall, High, Mid, and Low.

Table S19: Sample Numbers for OBGYN outcomes. Categorical OBGYN phenotypes. Ancestry is coded as: AA (African), EA (European), ESA (East Asian), HA (Hispanic), SAS (South Asian).

Table S20: Sample numbers for Alcohol use categorical phenotypes. Categorical phenotypes for alcohol use in Bio*Me*™: Never used, Not currently using, Not used, Yes using and not asked. Ancestry is coded as: AA (African), EA (European), ESA (East Asian), HA (Hispanic), SAS (South Asian).

Table S21: Sample numbers for Illicit drug use categorical phenotypes. Categorical phenotypes for illicit drug use in Bio*Me*™. Ancestry is coded as: AA (African), EA (European), ESA (East Asian), HA (Hispanic), SAS (South Asian).

Table S22: Sample numbers for categorical tobacco use phenotypes. Categorical phenotypes for tobacco use in Bio*Me*™: Tobacco use and type. Ancestry is coded as: AA (African), EA (European), ESA (East Asian), HA (Hispanic), SAS (South Asian).

Table S23: Sample numbers for categorical sexual history phenotypes. Categorical phenotypes for sexual history in Bio*Me*™. Ancestry is coded as: AA (African), EA (European), ESA (East Asian), HA (Hispanic), SAS (South Asian).
