## Supplemental Author List for "The Phenome-wide Consequences of Anorexia Nervosa Genes"

Roger A. H. Adan^1-3^, Lars Alfredsson^4^, Tetsuya Ando^5^, Ole A. Andreassen^6^, Harald Aschauer^7^, Jessica H. Baker^8^, Vladimir Bencko^9^, Andrew W. Bergen^10,11^, Wade H. Berrettini^12^, Andreas Birgegård^13^, Joseph M. Boden^14^, Ilka Boehm^15^, Vesna Boraska Perica^16,17^, Harry Brandt^18^, Gerome Breen^19,20^, Julien Bryois^13^, Cynthia M. Bulik^8,13, 21^, Roland Burghardt^22^, Laura Carlberg^23^, Matteo Cassina^24^, Sven Cichon^25-27^, Maurizio Clementi^24^, Jonathan R. I. Coleman^19,20^, Roger D. Cone^28^, Philippe Courtet^29^, Steven Crawford^18^, Scott Crow^30^, James J. Crowley^31,32^, Unna N. Danner^2^, Oliver S. P. Davis^33-35^, Martina deZwaan^36^, George Dedoussis^37^, Daniela Degortes^38^, Janiece E. DeSocio^39^, Danielle M. Dick^40-42^, Dimitris Dikeos^43^, Christian Dina^44^, Monika Dmitrzak-Weglarz^45^, Elisa Docampo^46-48^, Laramie E. Duncan^49^, Philibert Duriez^50,51^, Karin Egberts^52^, Stefan Ehrlich^15^, Geòrgia Escaramís^46-48^, Tõnu Esko^53,54^, Thomas Espeseth^55,56^, Xavier Estivill^46-48,57^, Anne Farmer^19^, Angela Favaro^38^, Fernando Fernández-Aranda^58,59^, Manfred M. Fichter^60,61^, Krista Fischer^53^, James A. B. Floyd^62^, Manuel Föcker^63^, Lenka Foretova^64^, Andreas J. Forstner^27,65,66^, Monica Forzan^24^, Christopher S. Franklin^16^, Steven Gallinger^67^, Giovanni Gambaro^68^, Héléna A. Gaspar^19,20^, Ina Giegling^69^, Paola Giusti-Rodríquez^31^, Fragiskos Gonidakis^43^, Scott Gordon^70^, Philip Gorwood^50,51^, Monica Gratacos Mayora^46-48^, Jakob Grove^71-74^, Sébastien Guillaume^29^, Yiran Guo^75^, Hakon Hakonarson^75,76^, Katherine A. Halmi^77^, Ken B. Hanscombe^19^, Konstantinos Hatzikotoulas^16,78^, Joanna Hauser^79^, Johannes Hebebrand^80^, Sietske G. Helder^19,81^, Anjali Henders^82^, Stefan Herms^25,26^, Beate Herpertz-Dahlmann^83^, Wolfgang Herzog^84^, Anke Hinney^80^, L. John Horwood^14^, Christopher Hübel^19,85^, Laura M. Huckins^86-89^, James I. Hudson^90^, Hartmut Imgart^91^, Hidetoshi Inoko^92^, Vladimir Janout^93^, Susana Jiménez-Murcia^58,59^, Craig Johnson^94^, Jessica S. Johnson^86,87^, Jennifer Jordan^95,96^, Antonio Julià^97^, Gursharan Kalsi^19^, Deborah Kaminská^98^, Allan S. Kaplan^99-101^, Jaakko Kaprio^102^, Leila Karhunen^103^, Andreas Karwautz^104^, Martien J. H. Kas^1,105^, Walter H. Kaye^106^, James L. Kennedy^99-101^, Martin A. Kennedy^107^, Anna Keski-Rahkonen^108^, Kirsty Kiezebrink^109^, Youl-Ri Kim^110^, Katherine M. Kirk^111^, Lars Klareskog^112^, Kelly L. Klump^113^, Gun Peggy S. Knudsen^114^, Maria C. La Via^8^, Mikael Landén^13,115^, Janne T. Larsen^72,85,116^, Stephanie Le Hellard^117,118^, Virpi M. Leppä^13^, Dong Li^75^, Paul Lichtenstein^13^, Lisa Lilenfeld^119^, Bochao Danae Lin^1^, Jolanta Lissowska^120^, Astri J. Lundervold^121^, Jurjen Luykx^1^, Pierre J. Magistretti^122-123^, Sarah L. Maguire^194^, Mario Maj^124^, Katrin Mannik^53,125^, Sara Marsal^97^, Christian R. Marshall^126^, Nicholas G. Martin^70^, Manuel Mattheisen^32,71,127,128^, Morten Mattingsdal^6^, Sara McDevitt^129,130^, Peter McGuffin^19^, Sarah E. Medland^111^, Andres Metspalu^53,131^, Ingrid Meulenbelt^132^, Nadia Micali^133,134^, James Mitchell^135^, Karen Mitchell^136,137^, Alessio Maria Monteleone^124^, Palmiero Monteleone^138^, Grant W. Montgomery^82,111,139^, Preben Bo Mortensen^72,85,116^, Melissa A. Munn-Chernoff^8^, Benedetta Nacmias^140,141^, Marie Navratilova^64^, Ioanna Ntalla^37^, Catherine M. Olsen^142^, Roel A. Ophoff^143,144^, Julie O’Toole^145^, Leonid Padyukov^112^, Aarno Palotie^54,102,146^, Jacques Pantel^147^, Hana Papezova^98^, Richard Parker^111^, John F. Pearson^148^, Nancy L. Pedersen^13^, Triinu Peters^80^, Liselotte V. Petersen^72,85,116^, Dalila Pinto^87^, Kirstin L. Purves^19^, Anu Raevuori^108,149^, Nicolas Ramoz^51^, Ted Reichborn-Kjennerud^114,150^, Valdo Ricca^151^, Samuli Ripatti^152^, Stephan Ripke^153-155^, Marion Roberts^19^, Alessandro Rotondo^156^, Dan Rujescu^69^, Filip Rybakowski^157^, Paolo Santonastaso^158^, André Scherag^159^, Stephen W. Scherer^160,161^, Ulrike Schmidt^162^, Nicholas J. Schork^163^, Alexandra Schosser^164^, Jochen Seitz^83^, Lenka Slachtova^165^, P. Eline Slagboom^132^, Margarita C. T. Slof-Op’t Landt^166,167^, Agnieszka Slopien^168^, Nicole Soranzo^16,169-171^, Sandro Sorbi^140,141^, Lorraine Southam^16,78^, Vidar W. Steen^172,173^, Michael Strober^174,175^, Garret D. Stuber^8,176^, Patrick F. Sullivan^8,13,31^, Beata Świątkowska^177^, Jin P. Szatkiewicz^31^, Ioanna Tachmazidou^16^, Friederike I. Tam^15,178^, Elena Tenconi^38^, Laura M. Thornton^8^, Alfonso Tortorella^179^, Federica Tozzi^180^, Janet Treasure^19^, Artemis Tsitsika^181^, Marta Tyszkiewicz-Nwafor^168^, Konstantinos Tziouvas^182^, Annemarie van Elburg^2,183^, Eric F. van Furth^166,167^, Tracey D. Wade^184^, Gudrun Wagner^104^, Esther Walton^15^, Hunna J. Watson^8,185,186^, Thomas Werge^187^, David C. Whiteman^142^, H-Erich Wichmann^188^, Elisabeth Widen^102^, D. Blake Woodside^100,101,189,190^, Jiayi Xu^86^, Shuyang Yao^13^, Zeynep Yilmaz^8,13,31,85^, Eleftheria Zeggini^16,78,191^, Stephanie Zerwas^8^, Stephan Zipfel^192,193^.

^1^Brain Center Rudolf Magnus, Department of Translational Neuroscience, University Medical Center Utrecht, Utrecht, The Netherlands.^2^Center for Eating Disorders Rintveld, Altrecht Mental Health Institute, Zeist, The Netherlands.^3^Sahlgrenska Academy, University of Gothenburg, Gothenburg, Sweden.^4^Institute of Environmental Medicine, Karolinska Institutet, Stockholm, Sweden. ^5^Department of Behavioral Medicine, National Institute of Mental Health, National Center of Neurology and Psychiatry, Kodaira, Tokyo, Japan. ^6^NORMENT KG Jebsen Centre, Division of Mental Health and Addiction, University of Oslo, Oslo University Hospital, Oslo, Norway.^7^Biopsychosocial Corporation, Vienna, Austria. ^8^Department of Psychiatry, University of North Carolina at Chapel Hill, Chapel Hill, North Carolina, USA. ^9^First Faculty of Medicine, Institute of Hygiene and Epidemiology, Charles University, Prague, Czech Republic.^10^BioRealm, LLC, Walnut, California, USA. ^11^Oregon Research Institute, Eugene, Oregon, USA.^12^Department of Psychiatry, Center for Neurobiology and Behavior, University of Pennsylvania Perelman School of Medicine, Philadelphia, Pennsylvania, USA.^13^Department of Medical Epidemiology and Biostatistics, Karolinska Institutet, Stockholm, Sweden.^14^Christchurch Health and Development Study, University of Otago, Christchurch, New Zealand.^15^Division of Psychological and Social Medicine and Developmental Neurosciences, Faculty of Medicine, Technische Universität Dresden, Dresden, Germany.^16^Wellcome Sanger Institute, Wellcome Genome Campus, Hinxton, Cambridge, UK.^17^Department of Medical Biology, School of Medicine, University of Split, Split, Croatia.^18^The Center for Eating Disorders at Sheppard Pratt, Baltimore, Maryland, USA.^19^Institute of Psychiatry, Psychology and Neuroscience, Social, Genetic and Developmental Psychiatry (SGDP) Centre, King’s College London, London, UK.^20^National Institute for Health Research Biomedical Research Centre, King’s College London and South London and Maudsley National Health Service Trust, London, UK.^21^Department of Nutrition, University of North Carolina at Chapel Hill, Chapel Hill, North Carolina, USA.^22^Klinikum Frankfurt/Oder, Frankfurt, Germany.^23^Medical University of Vienna, Vienna, Austria.^24^Clinical Genetics Unit, Department of Woman and Child Health, University of Padova, Padova, Italy.^25^Institute of Medical Genetics and Pathology, University Hospital Basel, Basel, Switzerland.^26^Department of Biomedicine, University of Basel, Basel, Switzerland.^27^Institute of Neuroscience and Medicine (INM-1), Research Center Juelich, Juelich, Germany.^28^Life Sciences Institute and Department of Molecular and Integrative Physiology, University of Michigan, Ann Arbor, Michigan, USA.^29^Department of Emergency Psychiatry and Post-Acute Care, CHRU Montpellier, University of Montpellier, Montpellier, France.^30^Department of Psychiatry, University of Minnesota, Minneapolis, Minnesota, USA.^31^Department of Genetics, University of North Carolina at Chapel Hill, Chapel Hill, North Carolina, USA.^32^Department of Clinical Neuroscience, Karolinska Institutet, Stockholm, Sweden.^33^MRC Integrative Epidemiology Unit, University of Bristol, Bristol, UK.^34^Bristol Medical School, University of Bristol, Bristol, UK.^35^The Alan Turing Institute, London, UK.^36^Department of Psychosomatic Medicine and Psychotherapy, Hannover Medical School, Hannover, Germany.^37^Department of Nutrition and Dietetics, Harokopio University, Athens, Greece.^38^Department of Neurosciences, University of Padova, Padova, Italy.^39^College of Nursing, Seattle University, Seattle, Washington, USA.^40^Department of Psychology, Virginia Commonwealth University, Richmond, Virginia, USA.^41^College Behavioral and Emotional Health Institute, Virginia Commonwealth University, Richmond, Virginia, USA.^42^Department of Human and Molecular Genetics, Virginia Commonwealth University, Richmond, Virginia, USA.^43^First Department of Psychiatry, National and Kapodistrian University of Athens, Medical School, Eginition Hospital, Athens, Greece.^44^L’institut du thorax, INSERM, CNRS, UNIV Nantes, Nantes, France.^45^Department of Psychiatric Genetics, Poznan University of Medical Sciences, Poznan, Poland.^46^Barcelona Institute of Science and Technology, Barcelona, Spain.^47^Universitat Pompeu Fabra, Barcelona, Spain.^48^Centro de Investigación Biomédica en Red en Epidemiología y Salud Pública (CIBERESP), Barcelona, Spain.^49^Department of Psychiatry and Behavioral Sciences, Stanford University, Stanford, California, USA.^50^GHU Paris Psychiatrie et Neurosciences, CMME, Paris Descartes University, Paris, France.^51^INSERM U1266, Institute of Psychiatry and Neurosciences, Paris, France.^52^Department of Child and Adolescent Psychiatry, Psychosomatics and Psychotherapy, University Hospital of Würzburg, Centre for Mental Health, Würzburg, Germany.^53^Estonian Genome Center, University of Tartu, Tartu, Estonia.^54^Program in Medical and Population Genetics, Broad Institute of Massachusetts Institute of Technology and Harvard University, Cambridge, Massachusetts, USA.^55^Department of Psychology, University of Oslo, Oslo, Norway.^56^Bjørknes College, Oslo, Norway.^57^Genomics and Disease, Bioinformatics and Genomics Programme, Centre for Genomic Regulation, Barcelona, Spain.^58^Department of Psychiatry, University Hospital of Bellvitge – IDIBELL and CIBERobn, Barcelona, Spain.^59^Department of Clinical Sciences, School of Medicine, University of Barcelona, Barcelona, Spain.^60^Department of Psychiatry and Psychotherapy, Ludwig-Maximilians-University (LMU), Munich, Germany.^61^Schön Klinik Roseneck affiliated with the Medical Faculty of the University of Munich, Munich, Germany.^62^Genomics plc, Genomics PLC, Oxford, UK.^63^Department of Child and Adolescent Psychiatry, University of Münster, Münster, Germany.^64^Department of Cancer, Epidemiology and Genetics, Masaryk Memorial Cancer Institute, Brno, Czech Republic.^65^Institute of Human Genetics, University of Bonn, School of Medicine and University Hospital Bonn, Bonn, Germany.^66^Centre for Human Genetics, University of Marburg, Marburg, Germany.^67^Department of Surgery, Faculty of Medicine, University of Toronto, Toronto, Ontario, Canada.^68^Division of Nephrology and Dialysis, Department of Medicine, AOVR, Ospedale Maggiore, Verona, Italy.^69^Department of Psychiatry, Psychotherapy and Psychosomatics, Martin Luther University of Halle-Wittenberg, Halle (Saale), Germany.^70^Genetics and Computational Biology Department, QIMR Berghofer Medical Research Institute, Brisbane, Queensland, Australia.^71^Department of Biomedicine, Aarhus University, Aarhus, Denmark.^72^The Lundbeck Foundation Initiative for Integrative Psychiatric Research (iPSYCH), Aarhus, Denmark.^73^Centre for Integrative Sequencing, iSEQ, Aarhus University, Aarhus, Denmark.^74^Bioinformatics Research Centre, Aarhus University, Aarhus, Denmark.^75^Center for Applied Genomics, Children’s Hospital of Philadelphia, Philadelphia, Pennsylvania, USA.^76^Department of Pediatrics, University of Pennsylvania Perelman School of Medicine, Philadelphia, Pennsylvania, USA.^77^Department of Psychiatry, Weill Cornell Medical College, New York, New York, USA.^78^Institute of Translational Genomics, Helmholtz Zentrum München – German Research Centre for Environmental Health, Neuherberg, Germany.^79^Department of Adult Psychiatry, Poznan University of Medical Sciences, Poznan, Poland.^80^Department of Child and Adolescent Psychiatry, University Hospital Essen, University of Duisburg-Essen, Essen, Germany.^81^Zorg op Orde, Delft, The Netherlands.^82^Institute for Molecular Bioscience, University of Queensland, Brisbane, Queensland, Australia.^83^Department of Child and Adolescent Psychiatry, Psychosomatics and Psychotherapy, RWTH Aachen University, Aachen, Germany.^84^Department of General Internal Medicine and Psychosomatics, Heidelberg University Hospital, Heidelberg University, Heidelberg, Germany.^85^National Centre for Register-Based Research, Aarhus BSS, Aarhus University, Aarhus, Denmark.^86^Pamela Sklar Division of Psychiatric Genomics, Icahn School of Medicine at Mount Sinai, New York, New York, USA.^87^Department of Psychiatry, and Genetics and Genomic Sciences, Icahn School of Medicine at Mount Sinai, New York, New York, USA.^88^Seaver Autism Center for Research and Treatment, Icahn School of Medicine at Mount Sinai, New York, New York, USA.^89^Mental Illness Research, Education and Clinical Centers, James J. Peters Department of Veterans Affairs Medical Center, Bronx, New York, USA.^90^Biological Psychiatry Laboratory, McLean Hospital/Harvard Medical School, Boston, Massachusetts, USA.^91^Eating Disorders Unit, Parklandklinik, Bad Wildungen, Germany.^92^Department of Molecular Life Science, Division of Basic Medical Science and Molecular Medicine, School of Medicine, Tokai University, Isehara, Japan.^93^Faculty of Health Sciences, Palacky University, Olomouc, Czech Republic.^94^Eating Recovery Center, Denver, Colorado, USA.^95^Department of Psychological Medicine, University of Otago, Christchurch, New Zealand.^96^Canterbury District Health Board, Christchurch, New Zealand.^97^Rheumatology Research Group, Vall d’Hebron Research Institute, Barcelona, Spain.^98^First Faculty of Medicine, Department of Psychiatry, Charles University, Prague, Czech Republic.^99^Centre for Addiction and Mental Health, Toronto, Ontario, Canada.^100^Institute of Medical Science, University of Toronto, Toronto, Ontario, Canada.^101^Department of Psychiatry, University of Toronto, Toronto, Ontario, Canada.^102^Institute for Molecular Medicine Finland FIMM, HiLIFE, Helsinki Institute of Life Science, University of Helsinki, Helsinki, Finland.^103^Institute of Public Health and Clinical Nutrition, Department of Clinical Nutrition, University of Eastern Finland, Kuopio, Finland.^104^Eating Disorders Unit, Department of Child and Adolescent Psychiatry, Medical University of Vienna, Vienna, Austria.^105^Groningen Institute for Evolutionary Life Sciences, University of Groningen, Groningen, The Netherlands.^106^Department of Psychiatry, University of California San Diego, San Diego, California, USA.^107^Department of Pathology and Biomedical Science, University of Otago, Christchurch, New Zealand.^108^Department of Public Health, University of Helsinki, Helsinki, Finland.^109^Institute of Applied Health Sciences, School of Medicine, Medical Sciences and Nutrition, University of Aberdeen, Aberdeen, UK.^110^Department of Psychiatry, Seoul Paik Hospital, Inje University, Seoul, Korea.^111^QIMR Berghofer Medical Research Institute, Brisbane, Queensland, Australia.^112^Division of Rheumatology, Department of Medicine, Center for Molecular Medicine, Karolinska Institutet and Karolinska University Hospital, Stockholm, Sweden.^113^Department of Psychology, Michigan State University, East Lansing, Michigan, USA.^114^Department of Mental Disorders, Norwegian Institute of Public Health, Oslo, Norway.^115^Department of Psychiatry and Neurochemistry, Institute of Neuroscience and Physiology, The Sahlgrenska Academy at the University of Gothenburg, Gothenburg, Sweden.^116^Centre for Integrated Register-based Research (CIRRAU), Aarhus University, Aarhus, Denmark.^117^Dr. Einar Martens Research Group for Biological Psychiatry, Center for Medical Genetics and Molecular Medicine, Haukeland University Hospital, Bergen, Norway.^118^Department of Clinical Medicine, Laboratory Building, Haukeland University Hospital, Bergen, Norway.^119^The Chicago School of Professional Psychology, Washington D.C., USA.^120^Department of Cancer Epidemiology and Prevention, M. Sklodowska-Curie National Research Institute of Oncology, Warsaw, Poland.^121^Department of Biological and Medical Psychology, University of Bergen, Bergen, Norway.^122^BESE Division, KAUST, KSA, King Abdullah University of Science and Technology, Thuwal, Saudi Arabia.^123^Department of Psychiatry, University of Lausanne-University Hospital of Lausanne (UNIL-CHUV), Lausanne, Switzerland.^124^Department of Psychiatry, University of Campania “Luigi Vanvitelli”, Naples, Italy.^125^Center for Integrative Genomics, University of Lausanne, Lausanne, Switzerland.^126^Department of Paediatric Laboratory Medicine, Division of Genome Diagnostics, The Hospital for Sick Children, Toronto, Ontario, Canada.^127^Center for Psychiatry Research, Stockholm Health Care Services, Stockholm City Council, Stockholm, Sweden.^128^Department of Psychiatry, Psychosomatics and Psychotherapy, University of Würzburg, Würzburg, Germany.^129^Department of Psychiatry, University College Cork, Cork, Ireland.^130^Child and Adolescent Regional Eating Disorder Service (CAREDS), Health Service Executive South, Cork, Ireland.^131^Institute of Molecular and Cell Biology, University of Tartu, Tartu, Estonia.^132^Molecular Epidemiology Section, Department of Biomedical Datasciences, Leiden University Medical Centre, Leiden, The Netherlands.^133^Department of Psychiatry, Faculty of Medicine, University of Geneva, Geneva, Switzerland.^134^Department of Pediatrics, Gynaecology and Obstetrics, University of Geneva, Geneva, Switzerland.^135^Department of Psychiatry and Behavioral Science, University of North Dakota School of Medicine and Health Sciences, Fargo, North Dakota, USA.^136^National Center for PTSD, VA Boston Healthcare System, Boston, Massachusetts, USA.^137^Department of Psychiatry, Boston University School of Medicine, Boston, Massachusetts, USA.^138^Department of Medicine, Surgery and Dentistry “Scuola Medica Salernitana”, University of Salerno, Salerno, Italy.^139^Queensland Brain Institute, University of Queensland, Brisbane, Queensland, Australia.^140^Department of Neuroscience, Psychology, Drug Research and Child Health (NEUROFARBA), University of Florence, Florence, Italy.^141^IRCCS Fondazione Don Carlo Gnocchi, Florence, Italy.^142^Population Health Department, QIMR Berghofer Medical Research Institute, Brisbane, Queensland, Australia.^143^Center for Neurobehavioral Genetics, Semel Institute for Neuroscience and Human Behavior, University of California Los Angeles, Los Angeles, California, USA.^144^Department of Psychiatry, Erasmus MC, University Medical Center Rotterdam, Rotterdam, The Netherlands.^145^Kartini Clinic, Portland, Oregon, USA.^146^Center for Human Genome Research, Massachusetts General Hospital, Boston, Massachusetts, USA.^147^INSERM U1124, Université de Paris, Paris, France.^148^Biostatistics and Computational Biology Unit, University of Otago, Christchurch, New Zealand.^149^Department of Adolescent Psychiatry, Helsinki University Hospital, Helsinki, Finland.^150^Institute of Clinical Medicine, University of Oslo, Oslo, Norway.^151^Department of Health Science, University of Florence, Florence, Italy.^152^Department of Biometry, University of Helsinki, Helsinki, Finland.^153^Analytic and Translational Genetics Unit, Department of Medicine, Massachusetts General Hospital and Harvard Medical School, Boston, Massachusetts, USA.^154^Stanley Center for Psychiatric Research, Broad Institute of the Massachusetts Institute of Technology and Harvard University, Cambridge, Massachusetts, USA.^155^Department of Psychiatry and Psychotherapy, Charité – Universitätsmedizin, Berlin, Germany.^156^Department of Psychiatry, Neurobiology, Pharmacology, and Biotechnologies, University of Pisa, Pisa, Italy.^157^Department of Psychiatry, Poznan University of Medical Sciences, Poznan, Poland.^158^Department of Neurosciences, Padua Neuroscience Center, University of Padova, Padova, Italy.^159^Institute of Medical Statistics, Computer and Data Sciences, Jena University Hospital, Jena, Germany.^160^Department of Genetics and Genomic Biology, The Hospital for Sick Children, Toronto, Ontario, Canada.^161^McLaughlin Centre, University of Toronto, Toronto, Ontario, Canada.^162^Institute of Psychiatry, Psychology and Neuroscience, Psychological Medicine, King’s College London, London, UK.^163^J. Craig Venter Institute (JCVI), La Jolla, California, USA.^164^Department of Psychiatry and Psychotherapy, Medical University of Vienna, Vienna, Austria.^165^First Faculty of Medicine, Department of Biology and Medical Genetics, Charles University, Prague, Czech Republic.^166^Center for Eating Disorders Ursula, Rivierduinen, Leiden, The Netherlands.^167^Department of Psychiatry, Leiden University Medical Centre, Leiden, The Netherlands.^168^Department of Child and Adolescent Psychiatry, Poznan University of Medical Sciences, Poznan, Poland.^169^Donor Health and Genomics, National Institute for Health Research Blood and Transplant Unit, Cambridge, UK.^170^Division of Cardiovascular Medicine, British Heart Foundation Centre of Excellence, Cambridge, UK.^171^Department of Haematology, University of Cambridge, Cambridge, UK.^172^Center for Medical Genetics and Molecular Medicine, Haukeland University Hospital, Bergen, Norway.^173^Department of Clinical Science, University of Bergen, Bergen, Norway.^174^Department of Psychiatry and Biobehavioral Science, Semel Institute for Neuroscience and Human Behavior, University of California Los Angeles, Los Angeles, California, USA.^175^David Geffen School of Medicine, University of California Los Angeles, Los Angeles, California, USA.^176^Department of Cell Biology and Physiology, University of North Carolina at Chapel Hill, Chapel Hill, North Carolina, USA.^177^Department of Environmental Epidemiology, Nofer Institute of Occupational Medicine, Lodz, Poland.^178^Eating Disorders Research and Treatment Center, Department of Child and Adolescent Psychiatry, Faculty of Medicine, Technische Universität Dresden, Dresden, Germany.^179^Department of Psychiatry, University of Perugia, Perugia, Italy.^180^Brain Sciences Department, Stremble Ventures, Limassol, Cyprus.^181^Adolescent Health Unit, Second Department of Pediatrics, “P. & A. Kyriakou” Children’s Hospital, University of Athens, Athens, Greece.^182^Pediatric Intensive Care Unit, “P. & A. Kyriakou” Children’s Hospital, University of Athens, Athens, Greece.^183^Faculty of Social and Behavioral Sciences, Utrecht University, Utrecht, The Netherlands.^184^School of Psychology, Flinders University, Adelaide, South Australia, Australia.^185^School of Psychology, Curtin University, Perth, Western Australia, Australia.^186^School of Paediatrics and Child Health, University of Western Australia, Perth, Western Australia, Australia.^187^Department of Clinical Medicine, University of Copenhagen, Copenhagen, Denmark.^188^Helmholtz Centre Munich – German Research Center for Environmental Health, Munich, Germany.^189^Centre for Mental Health, University Health Network, Toronto, Ontario, Canada.^190^Program for Eating Disorders, University Health Network, Toronto, Ontario, Canada.^191^TUM School of Medicine, Technical University of Munich and Klinikum Rechts der Isar, Munich, Germany.^192^Department of Internal Medicine VI, Psychosomatic Medicine and Psychotherapy, University Medical Hospital Tuebingen, Tuebingen, Germany.^193^Centre of Excellence for Eating Disorders (KOMET), University Tuebingen, Tuebingen, Germany. ^194^School of Medicine, InsideOut Institute, Sydney, New South Wales, Australia.
