## Supplementary Text for "The Phenome-wide Consequences of Anorexia Nervosa Genes"

**SUPPLEMENTAL METHODS**

A graphical version of our methods can be found in **Figure S10**.

***BMI Stratification***

We stratified our cohort into three categories of BMI: High, Mid, and Low. In order to determine which individuals fell into each group, we looked at the distribution of BMI for each ancestry group and sex (**Figures S1 and S2**). Within each distribution, we decided to use the quartiles of the normal distribution as our cut-offs for assigning BMI group (**Figure S1**). Individuals whose BMI fell above the 3^rd^ quartile of the BMI distribution were assigned to the High BMI category, those who fell below the 1^st^ quartile were assigned to Low BMI, and those whose BMI fell between the 1^st^ and 3^rd^ quartile of the distribution were assigned to the Mid BMI group. **Tables S2** and **S3** describe the BMI values for each ancestry and sex, as well as the range of BMI values included in each group for each ancestry and sex.

***Bio*Me *BRSPD File QC***

All of the Bio*Me*-R Structured Phenotype Database (BRSPD) files underwent a fairly extensive QC process to correct misspelled variable fields, remove extraneous symbols, standardize units of measure, and replace spaces with underscores (“_”). All of the files were originally delimited by the “|” symbol, and thus were converted to a tab-delimited format for pheWAS use. Many files had variable fields with variations on the same phenotype or measure (for example 1mg/mL could be written as “1_mg/mL”, “1mg/mL”, “1mg1mL”, “1mg/1mL”, etc.). These measures and phenotypes needed to be collapsed and standardized before input into pheWAS. Likewise, there were a few phenotypes, specifically the vital sign measurements, where each individual had many measurements, with each measurement representing an encounter with the healthcare system. In order to look at the overall association with these phenotypes, continuous vital sign measures with multiple entries were collapsed into four measures: highest measure recorded, lowest measure recorded, mean measure, and variance of measure.

*Allergies*

Known allergies and notes on allergic reaction were entered in the original Allergy file, which included individual ID, allergen, date of entry, and notes on the allergic reaction (if available). Most QC of the allergy file entailed collapsing variables of the same or similar phenotype. For example, allergy to tree nuts could be noted as “TREE NUT”, “TREE NUTS”, “TREE NUT UNSPECIFIED”, and “NUT, TREE”. All of these phenotypes were collapsed into the category of “TREE NUT”. After QC of the Allergy file and application of selection criteria for sample size and phenotype counts, we had a total of 113 Allergy phenotypes available for pheWAS. These phenotypes are listed in **Table S13**.

*Encounter Diagnosis*

The encounter diagnosis file comprises the primary diagnoses, denoted by the International Classification of Disease (ICD) 9 and 10 codes, assigned to each patient by the clinician in their visits and interactions with the health system. Most diagnosis codes in the encounter diagnosis file are ICD-10 codes, however, there were a few diagnoses that were still using ICD-9 codes that had not been converted. We converted the original encounter diagnosis file, which consisted of information on patient ID, diagnosis code, date of entry, notes, and code description, into counts of each diagnosis code per individual. The PheWAS R package *createPhewasTable* function uses this count information to select cases of a particular diagnosis to have a count of 2 or more diagnosis codes, select controls with counts of 0 and exclude individuals with only 1 count of a particular code. After QC, we had a total of 2,178 phenotypes available for pheWAS (**Table S7**).

*Encounter Orders*

The encounter orders file describes the various lab tests and other procedures ordered by the clinician upon the individual’s encounter with the healthcare system. Similar to the encounter diagnosis file, the encounter orders file includes individual ID, the encounter order procedure code, date of encounter, any notes from the clinician, and notes on whether the order was complete or currently being carried out. For this phenotype, each individual was noted as “TRUE” or “FALSE” as to whether they were assigned a particular order. After QC, there were a total of 1,609 phenotypes available for pheWAS (**Table S14**). Descriptions of the procedure codes were available in the original encounter orders file.

*Family History*

Family history of disease phenotypes were noted with the relation with the disorders such as mother, father, sister, brother, paternal grandmother, maternal grandmother, etc. We collapsed each phenotype based on the relationship degree in order to reduce the number of tests. Relationships of mother, father, sister, brother, son, daughter, and child were categorized as “first degree”. Grandchild, half-brother, half-sister, maternal aunt, maternal grandfather, maternal grandmother, maternal uncle, paternal aunt, paternal grandfather, paternal grandmother, and paternal uncle were categorized as “second degree”. Cousin, maternal great aunt, maternal great grandfather, maternal great grandmother, maternal great uncle, paternal great aunt, paternal great grandfather, paternal great grandmother, and paternal great uncle were categorized as “third degree”. Foster, spouse, stepbrother, stepfather and stepsister were categorized as “no relation”. After QC, there were a total of 144 phenotypes for pheWAS. All available phenotypes are listed in **Table S15**.

*Medications*

We used CLAMP, a clinical natural language processing algorithm, to identify the core ingredients of unique Bio*Me*™ prescriptions from the EHR-derived medications file. Using the unstructured Bio*Me*™ prescriptions as input, CLAMP returns an RxNorm concept unique identifier (RXCUI) for any prescription that contains a suspected drug name. After manual review of 100 random RXCUIs output by CLAMP, each RXCUI from CLAMP was then reintroduced to the RxNorm API to standardize the drugs by their ingredients. From this we were able to collapse and standardize medication names for PheWAS based on their common drug name. For example, sertraline, the generic name for the brand-name SSRI Zoloft, would encompass prescriptions for both the generic name “sertraline” as well as “Zoloft”. After QC a total of 951 medication phenotypes were available for pheWAS (**Table S11**).

*Personal History*

Personal history phenotypes were extracted from a large 156-question questionnaire that had information on demographics, social history, education, behavior, personal medical history, and more. After QC, a total of 35 phenotypes were available for pheWAS. **Table S16** lists all phenotypes available for analysis.

*Social History (Alcohol, Illicit Drug and Tobacco use; Sexual History)*

Social history variables containing information on education, tobacco and alcohol use, illicit drug use, and sexual history were available within the BRSPD files. We split this file into multiple phenotype files encompassing categorical and continuous measures of tobacco, alcohol and illicit drug use, as well as sexual history. Continuous measures of alcohol use were measured as ounces of alcohol consumed per week, illicit drug use was measured as frequency or number of times used, and tobacco use was separated into two continuous measures of number of cigarette packs smoked per day and number of years of tobacco use. Categorical measures of alcohol, illicit drug and tobacco use included Y/N questions on use and exposure (yes/no) and type of user (current, past, never), as well as type of tobacco (cigarettes, cigars, pipes or chew). Sexual history consisted of categorical measures of sexual activity and contraceptive use. Phenotypes available for social history are described in **Table S9**. Distributions of continuous phenotypes are shown in **Figure S11**.

*OBGYN History*

Obstetric history was derived from a file that included individual, type of pregnancy and/or parity (term, preterm, ectopic, abortion, therapeutic abortion, spontaneous abortion, para, and gravida), and date of OBGYN encounter. We took the OBGYN file and created two phenotype types for our pheWAS: (1) categorical Y/N for presence of a certain OBGYN outcome (2) continuous counts of the OBGYN outcome (**Figure S12**). **Table S17** describes OBGYN phenotypes available.

*Vital Signs (Height, Weight, Blood Pressure, Temperature, Respirations, Pulse, Pulse Oximetry, Pain Score, and Pain Location)*

The original vital signs file included information on Individual ID, type of vital sign, vital sign measure, unit of measure, and date of measurement for nine signs: blood pressure, height, pain score and location, pulse, pulse oximetry, respirations, temperature, and weight. Multiple measures were made for each individual’s encounter with the Mount Sinai health system. To account for these multiple measures we first removed any measurements that were clear, biologically impossible outliers (e.g. an individual who had a height measurement of 1,200 inches), then standardized all measurements to the same unit (e.g. cm for height, kg for weight and degrees Celsius for temperature), and finally summarized each individual’s measurements into four measures per phenotype: highest recorded measurement, lowest recorded measurement, mean measurement and variance. Blood pressure measurements were given as separate systolic and diastolic pressure measurements, and we also included a measure of pulse pressure, which was calculated as the difference between systolic and diastolic measurements. **Table S6** and **Figure S13** describe vital sign phenotypes available and their distributions.

For the weight phenotype, we chose to use three measurements: highest recorded weight, lowest recorded weight, and weight change over time, which was calculated by using the earliest and most recent recorded weight measures divided by the number of years between those records as the average number of kg change per year. This obviously will not reflect weight cycling and other trajectories over time, but gives us a rough mean estimate of weight fluctuation over time. Future analyses will include more precise weight trajectories for our pheWAS. Distribution of weight phenotypes is shown in **Figure S14**.

For pain score and location, we created measurements that we hoped captured both the severity of pain recorded (pain score is a 0 to 10 scale, with 0 as no pain and 10 being the most severe pain ever experienced), as well as the number of times an individual reported a particular pain score. We created pain score measurements that included highest recorded (with a maximum value of 10), mean pain score, and pain score sum, which was the sum of all recorded pain score measurements. In addition to pain score, we also had pain location recorded with pain score. We further created pain score by location, where each location was assessed for highest, mean, and sum pain scores. We looked at pain location overall, regardless of score, as an additional phenotype. **Table S10** and **Figure S15** describe pain score phenotypes available and their distributions.

**Testing for hidden case contamination**

Consider a binary trait, D_x_, which occurs in *N_+_* cases within Bio*Me*™. If there is no association between GReX and D_x_, we expect average GReX to be equal among cases and controls; essentially, we observe the same difference in gene expression as if we had randomly sampled any two groups of individuals.

If there is an association between G and D_x_ that is driven by unidentified case contamination (that is, by individuals within our case group that are incorrectly missing information about AN diagnoses) then we can estimate the expect effect size due to this contamination.

First, we assume that all other individuals, with or without the trait D_x,_ have GReX with mean G and standard deviation $g_{var}$.

Next, we assume all GReX differences are driven by contamination, which occurs at prevalence *p* and with effect size $\beta$ on gene expression. Assuming a well-designed and adequately powered GWAS and S-PrediXcan study, as here, we may estimate $\beta$ directly from S-PrediXcan summary statistics.

**Among controls:**

We estimate average GReX as the sum of $\left( G \pm g_{var} \right)$ for all $N_{-}$ individuals not expressing trait D_x_, as follows:

$$\bar{{GReX}_{-}}=G= \frac{\sum_{i=1}^{N_{-}} \left( G \pm g_{var} \right)}{N_{-}}$$

Similarly, it is trivial to derive the expected variance of this distribution:

$$\sigma_{-}^{2}= {g_{var}}^{2}=\frac{\sum_{i=1}^{N_{-}} {\left( (G \pm g_{var} \right)-\bar{x})}^{2}}{N_{-}}$$

**Among cases**:

For the subset of individuals ($N_{AN})$ with undiagnosed or undeclared AN, GReX will be up- or downregulated by S-PrediXcan effect size $\beta$;

For all other individuals ($N_{AN}+1: N_{+}$), GReX follows the previously defined normal distribution:

$$\bar{{GReX}_{+}}= \frac{N_{AN}(\beta)G +G(N_{+}-N_{AN})}{N_{+}}$$

To simplify: $p$ is the proportion of diagnosis-contaminated cases, such that $pN_{+}=N_{AN}$

$$=\frac{\left( G \right)\left( pN_{+}\beta\right)+G\left( N_{+}-pN_{+} \right)}{N_{+}}$$

$$=\frac{pN_{+}\left( \beta\right)G+G(N_{+})(1-p)}{N_{+}}$$

$$= p\left( \beta\right)G+G(1-p)$$

Similarly, we may derive the expected variance among cases as follows:

$$\sigma_{+}^{2}=\frac{N_{AN}\sigma_{AN}^{2}+\left( N_{+}-N_{AN} \right){g_{var}}^{2}+N_{AN}\left( \beta G-\bar{x} \right)^{2}+\left( N_{+}-N_{AN} \right)\left( G-\bar{x} \right)^{2}}{N_{+}}$$

To calculate the expected variance among individuals with undeclared AN:

$$\sigma_{AN}^{2}=\frac{1}{N}\sum_{i=1}^{N} ({\beta\chi_{i}-\beta G)}^{2}$$

$$=\frac{1}{N}\sum_{i=1}^{N} {\beta(\chi_{i}-G)}^{2}$$

$$=(\beta^{2})\frac{1}{N}\sum_{i=1}^{N} {(\chi_{i}-G)}^{2}$$

$$={{(\beta^{2})g}_{var}}^{2}$$

To simplify the expected case variance: we consider that $pN_{+}=N_{AN}$, $\bar{x}=\bar{{GReX}_{+}}$, and $\sigma_{AN}^{2}={{(\beta^{2})g}_{var}}^{2}$

$$\sigma_{+}^{2}=\frac{{pN}_{+}{{(\beta^{2})g}_{var}}^{2}+\left( N_{+}-{pN}_{+} \right){g_{var}}^{2}+{pN}_{+}{(\beta G-\bar{{GReX}_{+}})}^{2}+(N_{+}-pN_{+}){(G-\bar{{GReX}_{+}})}^{2}}{N_{+}}$$

$$=p(\beta^{2})\left( {g_{var}}^{2} \right)+\left( 1-p \right){g_{var}}^{2}+p{(BG-\bar{{GReX}_{+}})}^{2}+(1-p){(G-\bar{{GReX}_{+}})}^{2}$$

**Comparing cases and controls**

The **expected difference** **δ** between D_x_ cases and controls (δ) may be calculated as follows:

$$=\bar{{GReX}_{+}}-\bar{{GReX}_{-}}= p\left( \beta\right)G+G\left( 1-p \right)-G$$

$$=G(p\beta+1-p-1)$$

$$=pG(\beta-1)$$

Finally, the statistical significance of the difference between the two distributions (x,y) may be estimated from first principles using a T-Score, as follows:

$$T=\frac{\bar{{GReX}_{+}}-\bar{{GReX}_{-}}}{\sqrt{\frac{\sigma_{+}^{2}}{N_{+}}+\frac{\sigma_{-}^{2}}{N_{-}}}}$$

Where individual terms are as calculated above.

In order to test this hypothesis, we simulated gene expression following a normal distribution with mean G=2 and standard deviation $g_{var}$=0.1 for (i) 1,000 cases and 1,000 controls; (ii) 1,000 cases and 10,000 controls; (iii) 1,000 cases and 30,000 controls.

We introduced diagnostic contamination into our case group (x) at seven different rates (p=0.1, 0.5, 0.05, 0.02, 0.01, 0.005, 0.001), at nine different effect levels ($\beta$ =1,2,3,4,5,10,0.1, 0.25, 0.5). Following the formulae above, samples within our case group (x) had expression levels $\beta\left( G \pm g_{var} \right)$.

For each p x$\beta$combination we performed 10,000 permutations, and calculated mean expression levels within and across distributions; expected variance within and across distributions; and T-scores and p-values of expected significance of the difference between distributions.

Across all p x$\beta$combinations and case/control proportions, the formulae derived above accurately estimate the observed values (**Figure S3; Supplemental Material**).

**SUPPLEMENTAL RESULTS**

***S-PrediXcan of AN***

We used a binomial test to look for tissue enrichment within our experiment-wide significant gene-tissue associations and found no significant enrichment of any tissue. We then looked at tissue enrichment across nominally significant gene-tissue associations (p<0.05) and found significant enrichment of whole blood tissue (p=5.77 x 10^-7^, proportion of tests=0.0232, proportion of hits=0.0290), as well as significant under-enrichment in CMC DLPFC (p<2.20 x 10^-16^, proportion of tests=0.0398, proportion of hits=0.0171) and GTEx Testis tissues (p=0.02994, proportion of tests=0.0335, proportion of hits=0.0306).

***AN-GReX PheWAS***

Full summary statistics from our AN-GReX PheWAS for all phenotypes can be found in **Table S18**.

***Allergies***

After QC, a total of 179 allergy phenotypes were tested for association with AN-GReX. In the overall Bio*Me*™ cohort, downregulation of *PFKFB4* was associated with allergy to both the antibiotic tetracycline (Skin Not Sun Exposed-*PFKFB4*, p=4.12 x 10^-6^) and gut motility drug metoclopramide (Brain cortex-*PFKFB4*, p=9.49 x 10^-6^). Similarly, in individuals with Mid and Low BMI, downregulation of *PFKFB4* was associated with allergy to tetracycline (Mid-Skin Not Sun Exposed-*PFKFB4*, p=1.13 x 10^-5^; Mid-Skin Sun Exposed-*PFKFB4*, p=2.90 x 10^-4^; Low-Skin Not Sun Exposed-PFKFB4, p=4.51 x 10^-4^). Downregulation of *PFKFB4* in individuals with Mid BMI was also associated with allergy to the diabetic drug metformin (Mid-Lung-*PFKFB4*, p=1.44 x 10^-4^) and upregulation to the analgesic acetaminophen (Mid-Esophagus Mucose-*PFKFB4*, p=9.28 x 10^-6^). Additionally, downregulation of *PFKFB4* was associated with allergy to the antibiotic vancomycin in High BMI individuals (High-Brain Cortex-*PFKFB4*, p=9.89 x 10^-5^; High-Cells Transformed lymphocytes-*PFKFB4*, p=3.37 x 10^-4^). Upregulation of *SLC2A10* was associated with allergy to morphine in the overall cohort (Colon transverse-*SLC2A10*, p=1.85 x 10^-5^; Esophagus Mucosa-*SLC2A10*, p=8.10 x 10^-5^). Additional allergy associations are provided in **Table S13**.

***Encounter Orders***

Encounter orders are coded orders from the EHR for various laboratory tests and procedures, dietary restrictions and other medication orders. After QC, there were a total of 2,147 unique encounter order codes tested for association with AN-GReX. Full summary statistics for encounter orders are provided in **Table S14**.

In the overall cohort, downregulation of *ARIH2*, *NCKIPSD* and *DALRD3*, and upregulation of *CCDC71, SPINK8* and *WDR6* were associated with encounter order “DIET24”, an order for a regular diet (Heart Left Ventricle-ARIH2, p=8.95 x 10^-6^; Multiple tissues-*NCKPISD*, p<2.21 x 10^-5^; Cells Transformed Lymphocytes-*DALRD3*, p=8.33 x 10^-6^; Brain Frontal Cortex-*CCDC71*, p=1.75 x 10^-5^; Brain Hippocampus-*SPINK8*, p=1.21 x 10^-5^; Brain Frontal Cortex-*WDR6*, p=1.02 x 10^-5^; Brain Putamen basal ganglia-*WDR6*, p=7.59 x 10^-6^; Brain Substantia Nigra-WDR6, p=3.79 x 10^-6^). ARIH2, CCDC71 and WDR6 were additionally associated with encounter order “OPH1122”, coded for “Ophthalmoscopy subsequent performed”, along with upregulation of *CCDC36* (Upregulation-Heart Left Ventricle-ARIH2, p=2.17 x 10^-5^; Upregulation-Multiple tissues-*CCDC36*, p<1.95 x 10^-5^; Downregulation-Multiple tissues-CCDC71, p<2.09 x 10^-5^; Downregulation-Brain Putamen basal ganglia-WDR6, p=1.84 x 10^-5^). Additional encounter orders associated with AN-GReX in the overall cohort include order “51701” for insertion of a bladder catheter, “97112” for Neuromuscular re-education, “96372” for therapeutic, prophylactic and diagnostic injections and infusions, “100696” for a pregnancy-induced hypertension panel, “102915” for IFE interpretation, “600536” for pelvic ultrasound and “NUR577” for calorie count (**Table S14**).

Downregulation of *ARIH2*, *CCDC36*, and *DALRD3* and upregulation of *CCDC71* and *WDR6* (all of which were all associated with “DIET24” in the overall cohort), were associated with encounter order “PRE5” for fall precautions in individuals with Low BMI (p<3.85 x 10^-5^; **Table S14**). Additional associations within individuals of Low BMI were downregulation of *SLC26A10* with order “100478” for surveillance of multi-drug-resistant gram-negative bacterial respiratory infection (Low-Liver-*SLC26A10*, p=1.70 x 10^-5^), downregulation of *FBLIM1* with code “600536” for non-obstetric pelvic ultrasound (Low-DLPFC-*FBLIM1*, p=3.49 x 10^-5^), and downregulation of *DALRD3* with “DIET13” for a clear liquid diet (Low-Liver-*DALRD3*, p=2.98 x 10^-5^). In individuals with High BMI, upregulation of ARIH2 was associated with code “82565” for a creatinine serum lab order (High-Brain Amygdala-ARIH2, p=2.87 x 10^-5^) and downregulation of WDR6 and TMEM89 was associated with code “51701” for bladder catheter insertion (High-Artery Aorta-TMEM89, p=1.77 x 10^-5^; High-Thyroid-WDR6, p=2.35 x 10^-5^). Additional encounter order codes associated with AN-GReX in individuals with High BMI were “DIET50” for a renal diet, “CON65” for a consult to social worker, “PR157” for post-diagnostic catheter nursing intervention, “PRL166” for physician notification, “101186” for an abdominal pain panel, “NUR17” for pain assessment, “82040” for albumin blood measurement, and “NUR463” for obtaining height and weight measures (**Table S14**).

***Family History***

After QC a total of 144 phenotypes were available from family history data derived from the EHR (**Table S15**). Family history phenotypes were initially collapsed based on degree of relatedness into “first degree”, “second degree” and “third degree” categories (see **Supplementary Methods**). Upregulation of *MGMT* was associated with a first-degree family history of HIV infection in both the overall cohort (p<3.33 x 10^-4^) and in individuals with low BMI (p<7.93 x 10^-4^) across multiple tissues. This association passed the phenotype-wide threshold of p=1.64 x 10^-5^ in individuals with low BMI (Low-Brain Hippocampus-*MGMT*, p=6.93 x 10^-7^; Low-Subcutaneous Adipose-*MGMT*, p=7.53 x 10^-6^; Low-Brain Caudate basal ganglia-*MGMT*, p=1.09 x 10^-5^; Low-Brain Amygdala-*MGMT*, p=1.14 x 10^-5^; Low-Colon Sigmoid-*MGMT*, p=1.46 x 10^-5^).

***Medications***

After QC, a total of 1,086 unique medications/drug components were tested for association with AN-GReX. All associations with medications are shown in **Table S11**. In the overall cohort, upregulation of MHC-gene *CLIC1* was associated with multiple insulin- and diabetes-related medications including glucagon (Spleen-*CLIC1*, p=4.20 x 10^-9^), insulin aspart, insulin detemir, insulin degludec, insulin glulisine, insulin glargine, insulin lispro, insulin isophane, regular insulin and liraglutide (Subcutaneous Adipose-*CLIC1*, p<3.27 x 10^-5^). Upregulation of *KREMEN1* and *RBM6*, and downregulation of *PFKFB4* were also associated with insulin- and diabetes-related phenotypes (Insulin aspart-Liver-*KREMEN1*, p=2.81 x 10^-5^; insulin glargine-Liver-*KREMEN1*, p=4.16 x 10^-5^; insulin isophane-Liver-*KREMEN1*, p=4.31 x 10^-5^; linagliptin-Skin Not Sun Exposed-*PFKFB4*, p=3.91 x 10^-5^; insulin isophane-Brain Substantia Nigra-*RBM6*, p=3.96 x 10^-5^). In the overall cohort, downregulation of *RNF123* was associated with the benzodiazepine sedative temazepam (Artery Aorta-*RNF123*, p=2.76 x 10^-6^), stimulant laxative sennosides (Esophagus Mucosa-*RNF123*, p=1.33 x 10^-5^; Skeletal Muscle-*RNF123*, p=6.88 x 10^-6^), and diuretic hydrochlorothiazide (Esophagus Muscularis-*RNF123*, p=3.73 x 10^-5^). Upregulation of *KREMEN1* was also associated with the hydrochlorothiazide (Adipose Visceral Omentum-*KREMEN1*, p=4.55 x 10^-5^). Downregulation of *PFKFB4* was associated with type 2 diabetes medication empagliflozin (Skin Not Sun Exposed-*PFKFB4*, p=1.58 x 10^-5^) and anti-diarrheal loperamide (Skin Not Sun Exposed-*PFKFB4*, p=2.19 x 10^-5^). Additional overall AN-GReX associations with medications include antibiotic bacitracin (DLPFC-*C3orf62*, p=4.28 x 10^-5^), SSRI antidepressant sertraline (Brain Cerebellum-*MGMT*, p=2.53 x 10^-5^), analgesic salicylic acid (Brain Caudate basal ganglia-*NICN1*, p=1.90 x 10^-5^), HIV antiviral lopinavir (Brain Hippocampus-*SLC26A6*, p=1.72 x 10^-6^) and immunosuppressant chloroquine (Cells Transformed lymphocytes-*SLC2A10*, p=3.71 x 10^-5^).

Within the BMI-stratified groups, we find that *CLIC1* upregulation is similarly associated with glucagon in individuals in the Mid BMI group (Mid-Spleen-*CLIC1*, p=2.09 x 10^-6^), and the association of *CLIC1* GReX with glucagon remains fairly similar across BMI groups (**Figure S9**). Chemotherapy drug Paclitaxel is associated with upregulation of *SUOX* (Mid-DGN Whole Blood-*SUOX*, p=1.15 x 10^-6^) and upregulation of *PFKFB4* with anti-muscarinic drug mirabegron in Mid BMI individuals (Mid-Esophagus Mucosa-*PFKFB4*, p=1.77 x 10^-6^). In individuals of High BMI, downregulation of MST1 was associated with anti-diarrheal hormone octreotide (High-Adrenal gland-MST1, p=4.71 x 10^-6^) and downregulation of *PFKFB4* was associated with antidiabetic repaglinide and anti-tremor and gut antispasmodic Hyoscyamine (Repaglinide-High-Skin Sun Exposed-*PFKFB4*, p=7.92 x 10^-6^; Hyoscyamine-High-Skin Not Sun Exposed-*PFKFB4*, p=1.39 x 10^-5^). Among Low BMI individuals, top associations include upregulation of *GPX1* with prescription for lanolin oil (Low-Adipose Visceral Omentum-*GPX1*, p=8.25 x 10^-6^; Low-Ovary-*GPX1*, p=3.33 x 10^-6^). Additionally, downregulation of *USP19* was associated with osteoporosis drug Teriparatide in individuals with Low BMI (Low-Pituitary-*USP19*, p=7.84 x 10^-6^). Additional associations of AN-GReX with medications are found in **Table S11**.

***OBGYN History***

OBGYN phenotypes were extracted from the EHR and contained the following categories along with date of outcome: abortion, current pregnancy, ectopic pregnancy, para (parity), gravida, preterm birth, spontaneous abortion, to-term birth and therapeutic abortion. PheWAS were run on two (iterations) of the data: (1) Overall having any phenotype as a categorical TRUE/FALSE and (2) continuous counts of each phenotype (Sample numbers are available in **Table S19**). Upregulation of *KREMEN1* in individuals with high BMI was associated with overall parity, or the number of births per gestation (Subcutaneous adipose-*KREMEN1*, p=1.16 x 10^-4^). Downregulation of *SLC26A10* was also associated with parity, but in individuals of low BMI (Subcutaneous adipose-*SLC26A10*, p=2.03 x 10^-5^). All associations are shown in **Table S17**. For continuous measures of OBGYN outcomes, *CLIC1* upregulation was associated with greater counts of preterm pregnancies in the overall cohort (Adipose Subcutaneous-*CLIC1*, p=4.05 x 10^-6^) and in High BMI individuals (High-Adipose Subcutaneous-*CLIC1*, p=1.55 x 10^-8^). Downregulation of *LAMB2* was also associated with number of preterm pregnancies in High BMI individuals (High-Adipose Subcutaneous-*LAMB2*, p=2.68 x 10^-6^). Additional associations of AN-GReX with continuous OBGYN outcomes are described in **Table S17**.

***Personal History***

We tested a total of 35 phenotypes listed as “Personal History of Disease” in Bio*Me*™ (**Table S16**). Upregulation of *ARIH2*, *C3orf62*, *CCDC36*, *CELSR3*, *DALRD3*, *NCKIPSD*, *NDUFAF3*, *NICN1* and *P4HTM*, and downregulation of *CCDC71*, *GPX1* and *WDR6*, were associated with personal history of Lupus in the overall cohort, as well as in individuals with Mid BMI, in multiple tissues (Overall, p<1.42 x 10^-3^; Mid, p<1.41 x 10^-3^)(**Figure S16**). All of these genes fall within the chromosome 3 locus from our S-PrediXcan results (**Figure 1, Table S4**). Additionally, MHC-gene *CLIC1* was also associated with a personal history of Lupus in the overall cohort (Skeletal Muscle-*CLIC1*, p=7.57 x 10^-4^; Spleen-*CLIC1*, p=6.72 x 10^-4^; Subcutaneous Adipose-*CLIC1*, p=2.01 x 10^-4^). In individuals with Mid BMI, upregulation of *ARIH2*, *C3orf62*, *CCDC36*, *CELSR3*, *DALRD3*, *NCKIPSD*, *NICN1*, *P4HTM* and *USP19*, and downregulation of *CCDC71*, *GPX1*, *LAMB2*, and *WDR6*, was associated with a personal history of high blood pressure (p<1.42 x 10^-3^). Similarly, upregulation of *ARIH2*, *CCDC36*, *CELSR3*, *DALRD3*, *NCKIPSD*, *NDUFAF3*, *P4HTM* and downregulation of *WDR6* was associated with a personal history of kidney disease in individuals with Mid BMI (p<1.39 x 10^-3^). Additional associations are shown in **Table S16**.

***Social History* (additional results)**

Sample numbers for social history phenotypes can be found in **Tables S20** through **S23**. Phenotypes of sexual history, such as sexual activity, contraceptive use and type, as well as information about sexual partner, were derived from the social history questionnaire. A total of eight phenotypes were tested for association with AN-GReX*: (1) Sexually active (Y/N) (2) Self-reported female or male partner (3) Contraceptive method: abstinence, condom, injection, IUD, and pill (*phenotypes are categorized as Y/N or T/F). In the overall cohort, upregulation of *RBM6* and *SEMA3F* and downregulation of *MST1R* and *RNF123* were associated with being sexually active across multiple tissues (*RBM6*, p<5.46 x 10^-3^; *SEMA3F*, p<4.51 x 10^-3^; *MST1R*, p<4.60 x 10^-3^; *RNF123*, p<4.50 x 10^-3^). Within our stratified BMI analyses, downregulation of *APEH* was associated with being sexually active in individuals of High BMI (High-Esophagus Muscularis-*APEH*, p=4.63 x 10^-5^). Additionally in individuals of High BMI, upregulation of *GPX1* was associated with IUD use (High-Esophagus Mucosa-*GPX1*, p=8.11 x 10^-5^). All associations are described in **Table S9**.

***Anthropometric traits and Vital signs***

Vital signs were measured for individuals at most if not all encounters with the healthcare system. Measurements of blood pressure, height, pain score and location, pulse, pulse oximetry, respirations, temperature and weight were recorded for each individual for each encounter. Vital sign measurements were standardized and QC of these measurements is described in the **Supplementary Methods**. Full results for vital signs are shown in **Table S6**.

*Blood Pressure*

Blood pressure measures were split into three factors: diastolic pressure, systolic pressure, and pulse pressure (see **Supplementary Methods**), with highest ever measure, lowest ever measure, mean measure and variance calculated for each phenotype. In the overall cohort, upregulation of *RBM6* and downregulation of *MST1R* and *RNF123* were associated with diastolic and systolic highest measurements in multiple tissues (Diastolic highest-*RBM6*, p<9.70 x 10^-4^; Diastolic highest-*MST1R*, p<9.27 x 10^-4^; Diastolic highest-*RNF123*, p<8.56 x 10^-4^; Systolic highest-*RBM6*, p<9.74 x 10^-4^; Systolic highest-*MST1R*, p<7.08 x 10^-4^; Systolic highest-*RNF123*, p<8.89 x 10^-4^). Upregulation of NCKIPSD, *NDUFAF3*, and *P4HTM*, and downregulation of *WDR6* were associated with lowest systolic pressure in the overall cohort (multiple tissues, p<9.68 x 10^-4^). BMI-stratified associations are described in the full summary statistics in **Table S6**.

*Height*

Height for individuals in Bio*Me*™ was standardized to centimeters units of measure (see **Supplementary Methods**). In the overall cohort, there were four genes associated with measures of mean height. Upregulation of *CLIC1*, *GPR75* and *LINC00324* and downregulation of *CTNNB1* were associated with mean height (Adipose Subcutaneous-*CLIC1*, p=1.39 x 10^-5^; Adipose Subcutaneous-*GPR75*, p=3.51 x 10^-4^; Artery Tibial-*GPR75*, p=5.07 x 10^-4^; Skin Sun Exposed-*LINC00324*, p=6.51 x 10^-4^; Brain Amygdala-*CTNNB1*, p=2.28 x 10^-4^; Brain Caudate basal ganglia-*CTNNB1*, p=1.67 x 10^-4^; Brain Putamen basal ganglia-*CTNNB1*, p=4.65 x 10^-4^). *GPR75* and *CLIC1* upregulation were also associated with mean height among the Mid BMI individuals (Mid-Adipose Subcutaneous-*CLIC1*, p=4.31 x 10^-4^; Mid-Artery Tibial-*GPR75*, p=1.44 x 10^-4^). Full summary statistics are available in **Table S6**.

*Pain Score and Location (Additional results)*

Pain score and location measurements were described within the vital sign files from the Bio*Me*™ BRSPD files. QC of pain score and pain location phenotypes are available in the **Supplementary Methods**. Final phenotypes available for pain score include highest ever, sum pain score and mean pain score. Pain location resulted in a total of 25 location phenotypes: abdomen, ankle, back, breast, chest, costal left, costal right, elbows, feet, generalized, groin, hands, head, knees, left leg, lower extremities, neck, pelvis, perineum, right leg, sacrum, shoulder, throat, upper extremities and wrist. Pain score by location phenotypes included measures of highest ever, sum and mean pain scores for each location type.

Upregulation of *SLC2A10* was associated with mean pain score in the overall cohort (Adrenal Gland-*SLC2A10*, p=9.94 x 10^-4^). Upregulation of *SLC2A10* was also associated with mean pain score in Low BMI individuals (Low-Adrenal Gland-*SLC2A10*, p=6.54 x 10^-5^). In High BMI individuals, downregulation of *MGMT* was associated with mean pain score in multiple tissues (High-*MGMT*, p<9.48 x 10^-4^). Downregulation of *PROS1* was also associated with mean pain score in the High BMI group (High-Artery Aorta-*PROS1*, p=8.06 x 10^-4^). Downregulation of *CLIC1* and downregulation of *MGMT* were associated with pain score sum in Low and High BMI groups respectively (Low- Testis*-CLIC1*, p=2.83 x 10^-4^; High-Spleen-*MGMT*, p=1.85 x 10^-5^).

For measures of pain location only, many genes were associated with pain location in the overall cohort, as well as among the stratified BMI groups. In the overall cohort, upregulation of *ARIH2*, *C3orf62, DALRD3*, *NCKIPSD* and downregulation of *GPX1* and *WDR6* were associated with throat pain. *CTNNB1* was associated with back (upregulation), chest (upregulation) and left leg pain (downregulation). Upregulation of *MST1R* and *RNF123* was associated with breast pain in the overall cohort, and upregulation of *CHAC2* and *P4HTM*, and downregulation of *LINC00324* were associated with costal left pain. All summary statistics for AN-GReX associations with pain location are located in **Table S10**. Within our stratified BMI groups, we see large numbers of associations of generalized pain location and foot pain with Low BMI individuals. There are a total of 55 gene-tissue associations of AN-GReX with generalized pain, and 129 associations with foot pain (Generalized p<1.88 x 10^-3^, Foot p <2.03 x 10^-3^). Some of the genes included in those associations include *RBM6* upregulation with generalized pain (32 associations, p<1.16 x 10^-3^), *MST1R* downregulation with generalized pain (13 associations, p<1.07 x 10^-3^), *CCDC36* downregulation with foot pain (19 associations, p<1.76 x 10^-3^), *NCKIPSD* downregulation with foot pain (33 associations, p<1.90 x 10^-3^), and upregulation of *WDR6* with foot pain (32 associations, p<2.03 x 10^-3^). Full summary statistics for each body location are available in **Table S10**.

When merging pain score with pain location, multiple AN genes were associated with measure of pain in various bodily locations (**Table S10**). In the overall cohort, upregulation of *CTNNB1*, *TMEM89* and *TNFSF12* was associated with throat pain score sum (Nerve Tibial-*CTNNB1*, p=4.92 x 10^-10^; Artery Aorta-*TMEM89*, p=1.25 x 10^-5^; Brain Cerebellar Hemisphere-*TMEM89*, p=2.39 x 10^-6^; Nerve Tibial-*TNFSF12*, p=8.20 x 10^-7^). Upregulation of *KREMEN1* was associated with highest ever elbow pain as well as mean elbow pain measures (Highest-Artery Aorta-*KREMEN1*, p=3.62 x 10^-6^; Mean-Artery Aorta-*KREMEN1*, p=1.43 x 10^-6^). Downregulation of *ASB3* and *SEMA3F*, and upregulation of *LAMB2* were associated with knee pain score sum (Cells EBV-transformed-*ASB3*, p=3.81 x 10^-5^; Brain Substantia nigra-*SEMA3F*, p=2.03 x 10^-6^; DGN Whole Blood-*LAMB2*, p=3.86 x 10^-5^). Upregulation of *EBF3* was associated with hand pain score sum, while downregulation of *PFKFB4* and *SPINK8* were associated with back pain score sum and costal left pain score sum respectively (**Table S10**). Upregulation of CTNNB1, SLC2A10, SPINK8, TNFSF12 and downregulation of KREMEN1 were associated with knee pain score measures.

Within the stratified BMI groups, many genes were associated with various pain scores and locations. *MST1* downregulation was associated with highest neck pain score in Mid BMI individuals (Multiple tissues, p<4.00 x 10^-5^). Among the Low BMI individuals, six genes were associated with abdomen pain score sum: *CLIC1*, *EBF3*, *GPR75*, *NICN1*, *SLC2A10* and *TNFSF12* (**Table S10**). Upregulation of *ARIH2*, *MST1* and *NICN1* in Low BMI individuals was associated with right leg pain score sum. In High BMI individuals, *CTNNB1*, *KREMEN1*, *SLC2A10*, *SPINK8* and *TNFSF12* were associated with measures of knee pain, and *PFKFB4*, *SUOX* and *NICN1* were associated with back pain, head pain, and ankle pain respectively. All summary statistics for pain score by location can be found in **Table S10**.

*Pulse*

Pulse measurements of beats per minute (bpm) were measured as the other vital sign continuous measurements as highest ever, lowest ever, mean and variance (**Supplemental Methods**). Downregulation of two genes, *NCKIPSD* and *TNFSF12*, were associated with mean pulse and pulse variance respectively in the overall cohort (Colon Transverse-*NCKIPSD*, p=1.60 x 10^-4^; Brain Substantia nigra-*TNFSF12*, p=9.20 x 10^-4^). Among individuals of Low BMI, a different set of genes is associated with measures of pulse. Downregulation of *ARIH2* was associated with highest pulse, while downregulation of *MST1R* and *RNF123* was associated with mean pulse (Low-Liver-*ARIH2*, p=2.57 x 10^-4^; Low-Colon Sigmoid-*MST1R*, p=5.81 x 10^-4^; Low-Pituitary-*RNF123*, p=5.65 x 10^-4^). Upregulation of *ARIH2OS* was associated with mean pulse in Low BMI individuals as well (Low-Nerve Tibial-*ARIH2OS*, p=7.52 x 10^-4^). Full summary statistics are located in **Table S6**.

*Pulse Oximetry*

AN-GReX was not significantly associated with measures of pulse oximetry in the overall Bio*Me*™ cohort, however, when we stratified by BMI, we did see significant associations of AN-GReX with pulse oximetry within the High and Low BMI individuals. Downregulation of *MGMT* was associated with mean pulse oximetry measurements in High BMI individuals (High-Brain Cerebellar Hemisphere-*MGMT*, p=6.84 x 10^-4^), while downregulation of *GPX1* and upregulation of *ARIH2* were associated with pulse oximetry variance in Low BMI individuals (Low-Ovary-*GPX1*, p=1.95 x 10^-4^; Low-Brain Amygdala-*ARIH2*, p=9.03 x 10^-4^). Full summary statistics are located in **Table S6**.

*Respirations*

Associations of AN-GReX with respirations, or breaths per minute, were measured using highest ever, lowest ever, mean and variance for the respiration phenotypes (**Supplemental Methods**). Downregulation of *ARIH2*, *CCDC36*, and *MST1R*, and upregulation of *CCDC71*, *GPX1*, *RBM6* and *WDR6* were associated with measures of highest ever respiration rate (Multiple tissues, p<9.84 x 10^-4^, **Table S6**). Additionally, downregulation of *CCDC36* and *P4HTM*, and upregulation of *GPX1*, were associated with mean respiration rate among the overall cohort (Multiple tissues, p<8.53 x 10^-4^). Among the BMI stratified groups, we find that downregulation of *GPX1* and upregulation of *NICN1* is associated with lowest ever respiration rates in High BMI individuals (Multiple tissues, p<8.99 x 10^-4^). Among Low BMI individuals, we see similar patterns of association of AN-GReX with respirations as we saw in the overall cohort. Downregulation of *ARIH2*, *NCKIPSD*, and *CCDC36*, and upregulation of *CCDC71* and *WDR6* are associated with highest respiration rate in the Low BMI individuals (Multiple tissues, p<9.93 x 10^-4^). Full summary statistics are located in **Table S6**.

*Temperature*

Temperature measurements were standardized to degrees Celsius (**Supplemental Methods**), and highest ever, lowest ever, mean and variance calculated. In the overall cohort, upregulation of *MGMT* was associated with mean temperature (Testis-*MGMT*, p=5.63 x 10^-4^). Among individuals in the Mid BMI group, downregulation of *EBF3* was associated with highest ever temperature (DLFPC-*EBF3*, p=7.24 x 10^-4^; Ovary-*EBF3*, p=7.15 x 10^-4^). Full summary statistics are located in **Table S6**.

*Weight (Additional results)*

Regression lines (with confidence intervals) for individual *CLIC1*-GReX plotted against highest and lowest recorded weight measures are shown in **Figure S17 and S18**.

Multiple genes were associated with weight change over time, which was measured by kg change per year, in all four BMI groups. Downregulation of *ARIH2*, *NCKIPSD*, *NDUFAF3* and *P4HTM* and upregulation of *WDR6* were associated with weight change over time in Overall, High and Low BMI groups in multiple tissues (**Figure S5**, **Table 3**). Downregulation of *RNF123* was associated with weight change over time in the overall cohort and in individuals with High BMI (Overall-Uterus-*RNF123*, p=3.18 x 10^-4^; High-Artery Aorta-*RNF123*, p=3.59 x 10^-4^), and upregulation of *RNF123* was associated with weight change in individuals with Mid and Low BMI (Mid-Brain Amygdala-*RNF123*, p=8.91 x 10^-5^; Mid-Cells Transformed Fibroblasts-*RNF123*, p=2.90 x 10^-4^; Low-Vagina-*RNF123*, p=5.16 x 10^-4^). Upregulation of *CCDC71* and *DHFR2*, and downregulation of *DALRD3* were associated with weight change over time in High and Low BMI individuals (High-Brain Cerebellar Hemisphere-*CCDC71*, p=4.71 x 10^-4^; High-Brain Cerebellum-*CCDC71*, p=1.67 x 10^-4^; High-Spleen-*CCDC71*, p=2.43 x 10^-4^; High-Thyroid-*CCDC71*, p=7.53 x 10^-4^; Low-Tibial Nerve-*CCDC71*, p=6.82 x 10^-4^; High-DGN Whole Blood-*DHFR2*, p=3.47 x 10^-4^; Low-DGN Whole Blood-*DHFR2*, p=7.48 x 10^-4^; High-Multiple tissues-*DALRD3*, p<8.21 x 10^-4^; Low-Brain Cerebellum-*DALRD3*, p=5.85 x 10^-4^; Low-Esophagus Gastroesophageal Junction-*DALRD3*, p=5.01 x 10^-4^). In individuals with High BMI, downregulation of *C3orf62*, *CCDC36*, *CELSR3* and *PROS1* were associated with weight change over time (High-DLPFC-*C3orf62*, p=8.59 x 10^-4^; High-Adipose Visceral Omentum-*C3orf62*, p=6.45 x 10^-4^; High-DGN Whole Blood-*C3orf62*; p=7.19 x 10^-4^;High-DGN Whole Blood-*CCDC36*, p=3.05 x 10^-6^; High-Subcutaneous Adipose-*CCDC36*, p= p=1.40 x 10^-5^; High-Tibial Nerve-*CELSR3*, p=1.09 x 10^-4^;High-Whole Blood-*PROS1*, p=3.46 x 10^-4^). Full summary associations of AN-GReX with weight change over time are shown in **Table 3**.

***Distribution of CLIC1-GReX by BMI***

To assess whether our BMI-group specific *CLIC1*-GReX associations are driven by differences in the distribution of GReX measures in these BMI subgroups, we tested for pairwise differences in the distribution of *CLIC1*-GReX for four tissues (Breast Mammary Tissue, Spleen, Subcutaneous Adipose and Whole Blood) using the Kolmogorov-Smirnov test (**Figure S19**). We found no significant difference in the distribution of *CLIC1*-GReX across BMI groups for any tissue (p_Significance_<0.0167; Breast Mammary Tissue, p_KS_>0.0495; Spleen, p_KS_>0.0368; Subcutaneous Adipose, p_KS_>0.9262; Whole Blood, p_KS_>0.2726).
