## Supplementary figures and images for "The Phenome-wide Consequences of Anorexia Nervosa Genes"

### Supplemental Figure 1

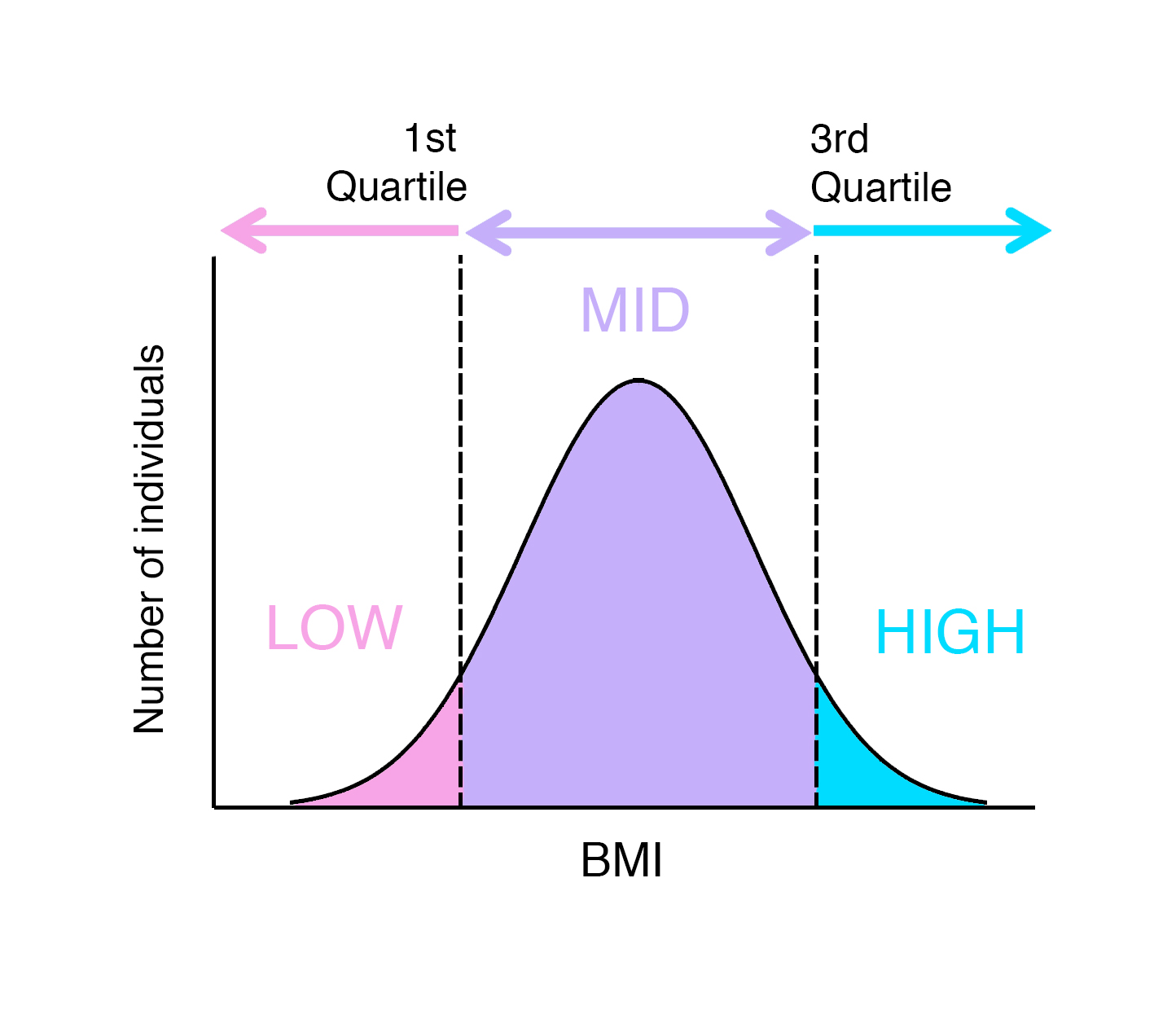

### Supplemental Figure 2

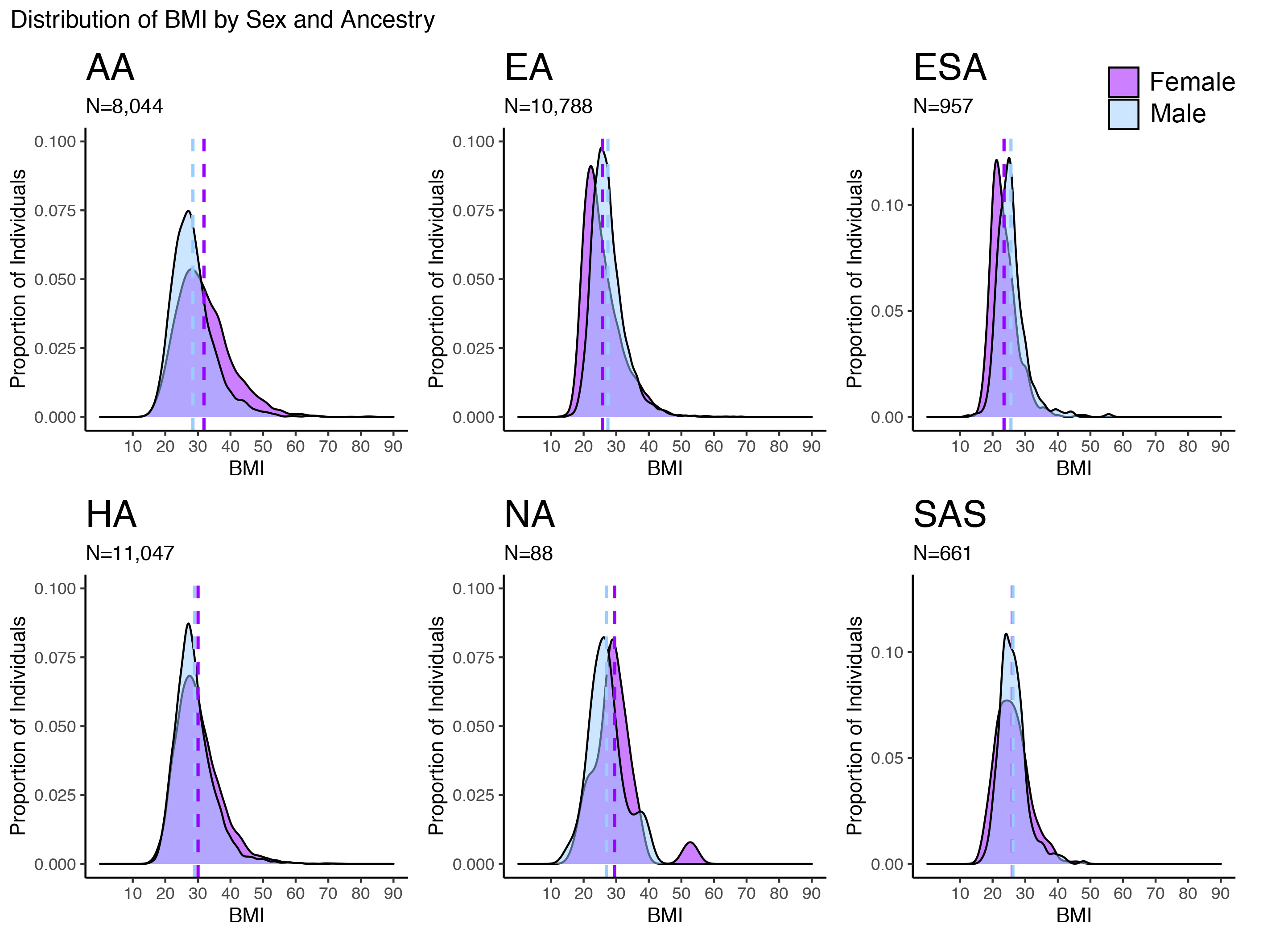

### Supplemental Figure 3A

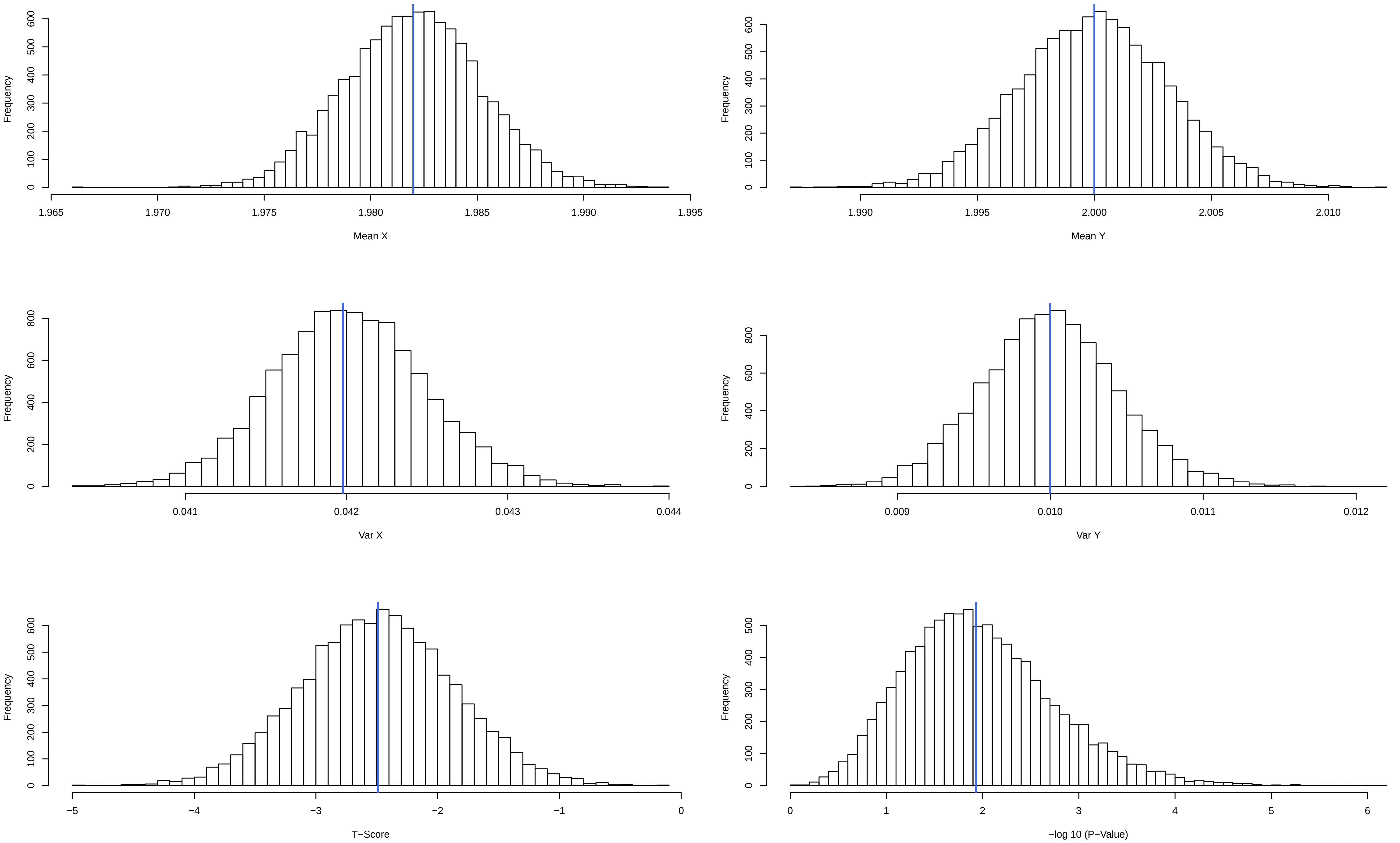

### Supplemental Figure 3B

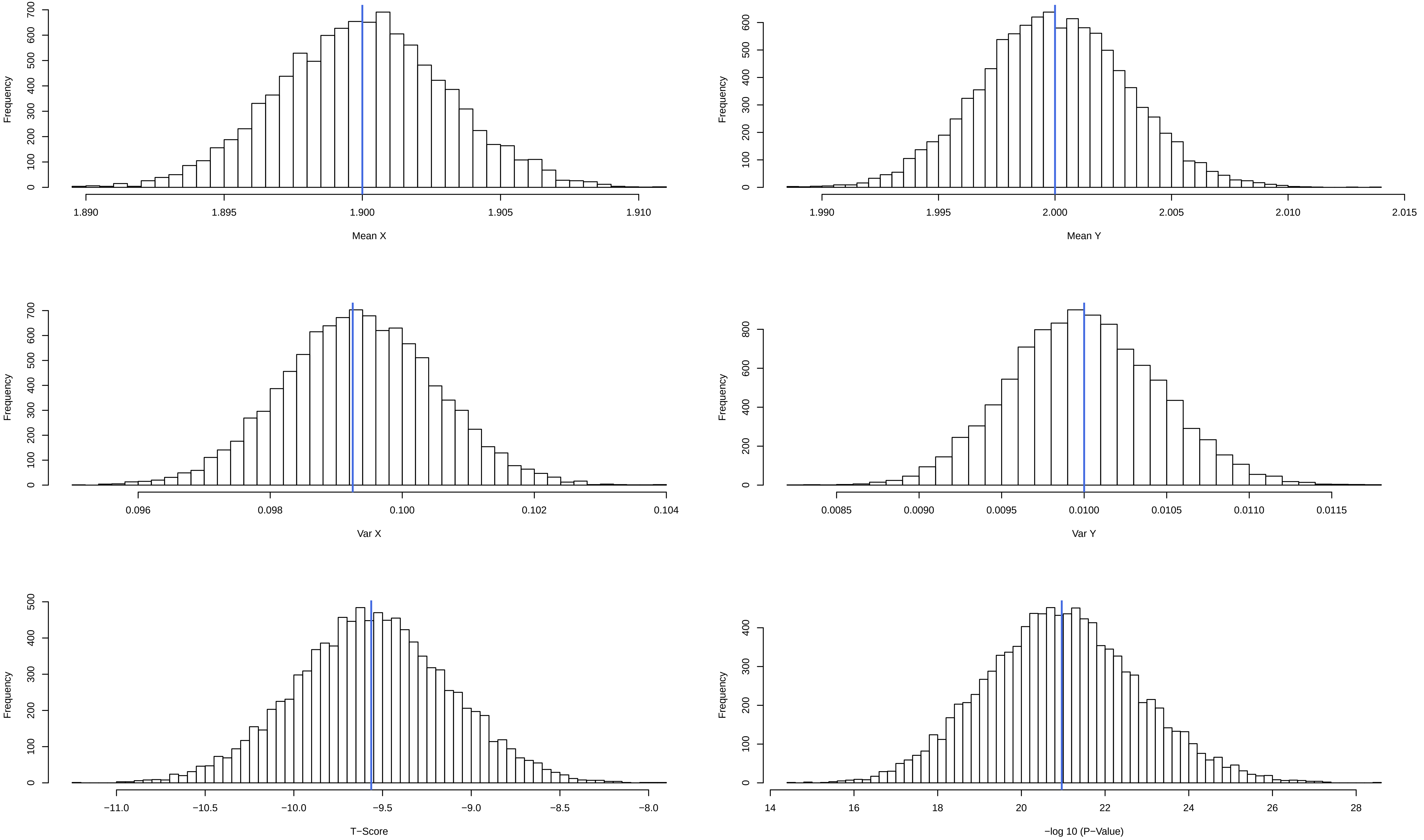

### Supplemental Figure 3C

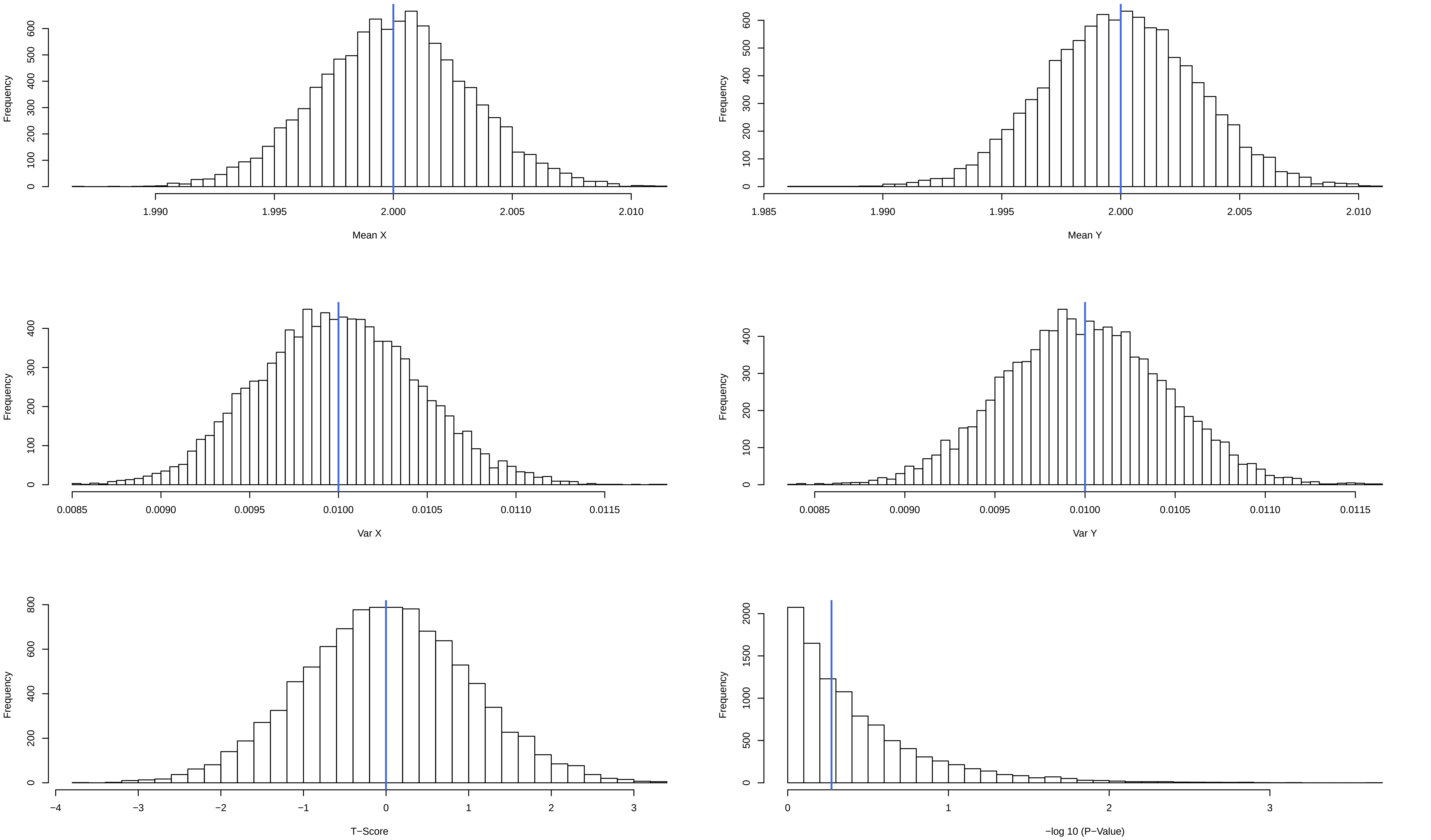

### Supplemental Figure 3D

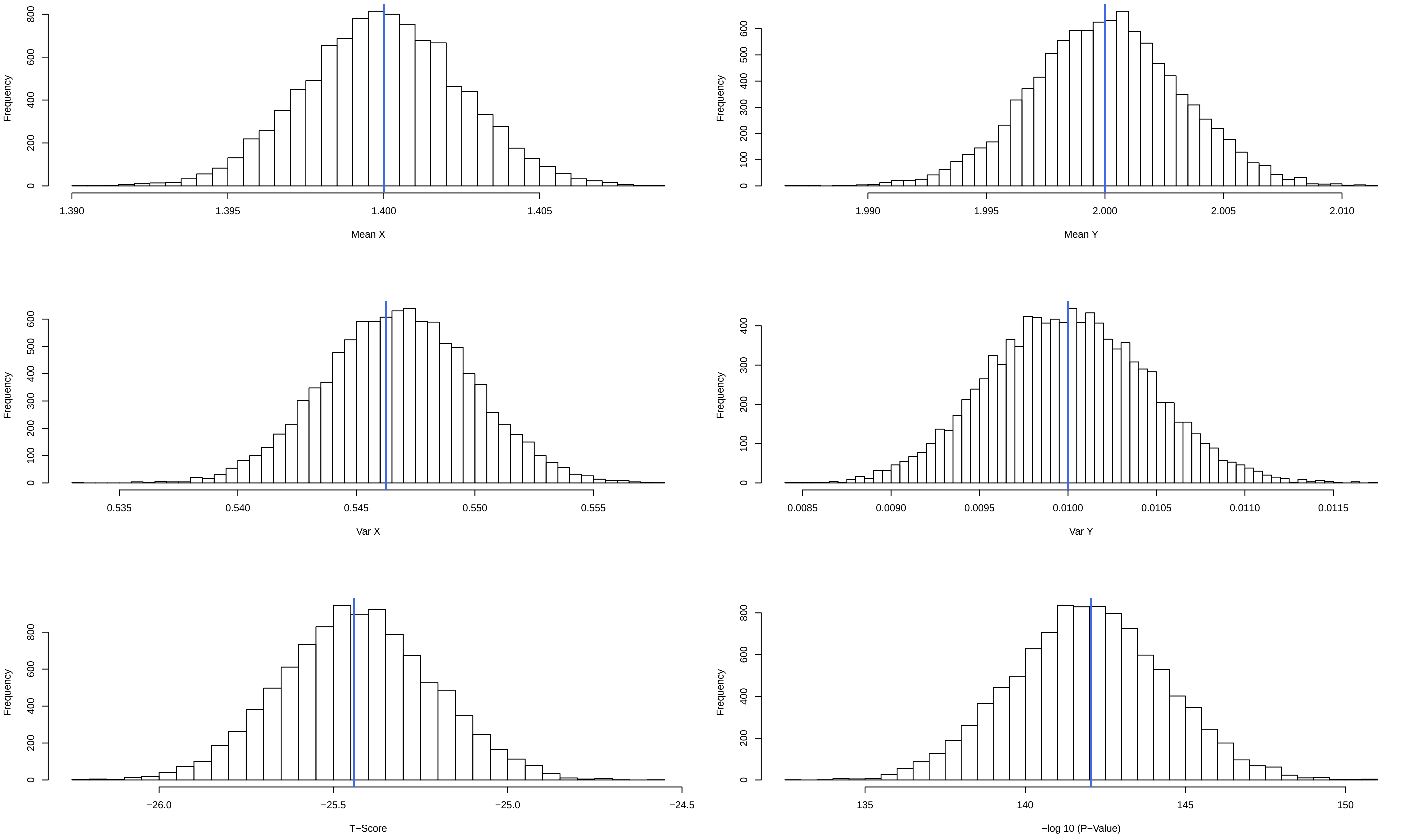

### Supplemental Figure 4

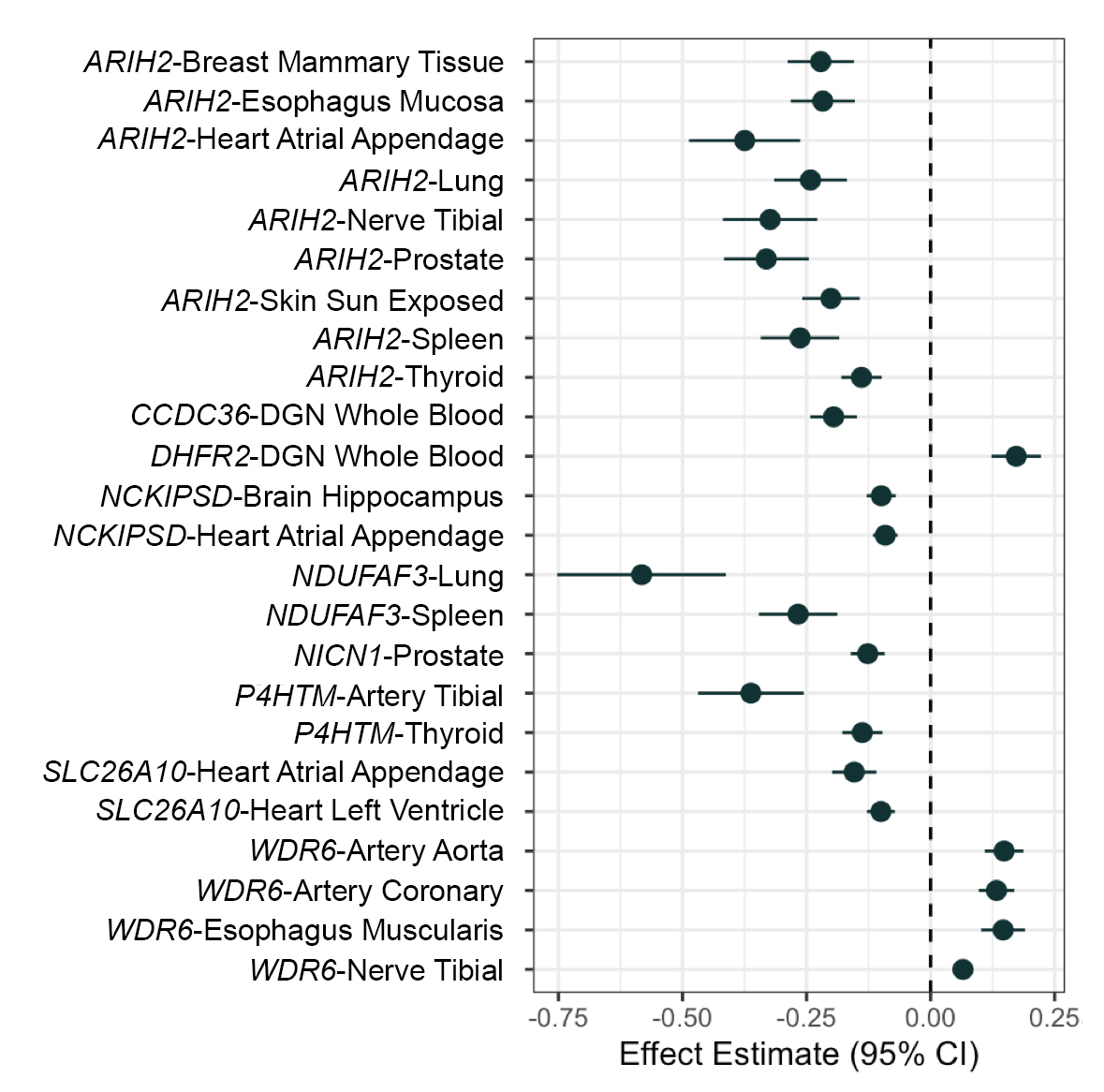

### Supplemental Figure 5

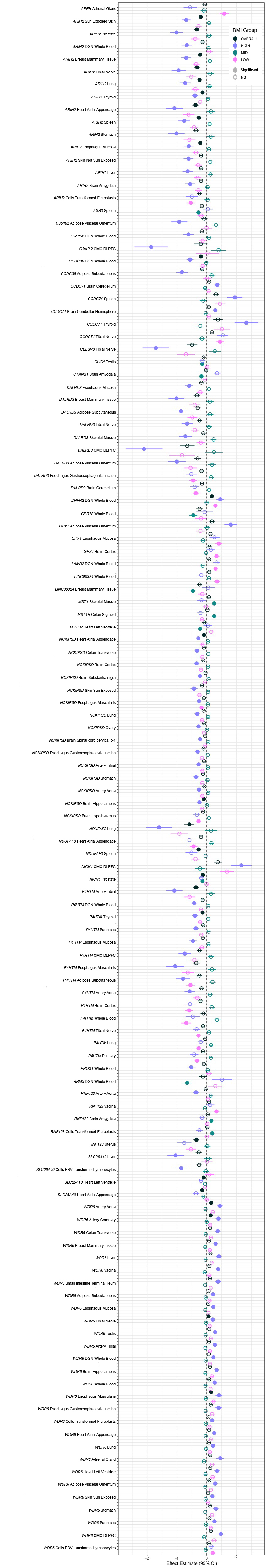

### Supplemental Figure 6

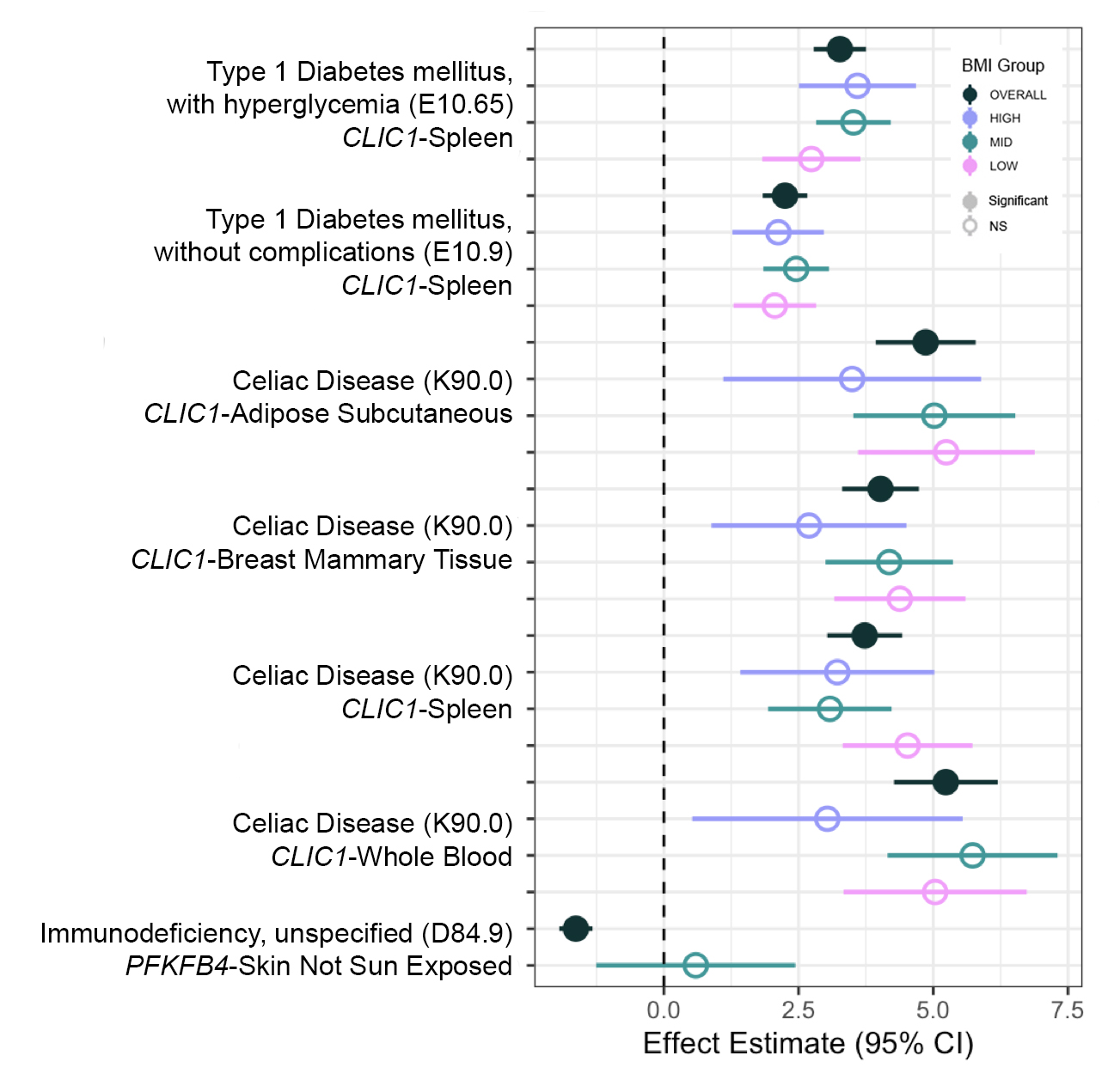

### Supplemental Figure 7

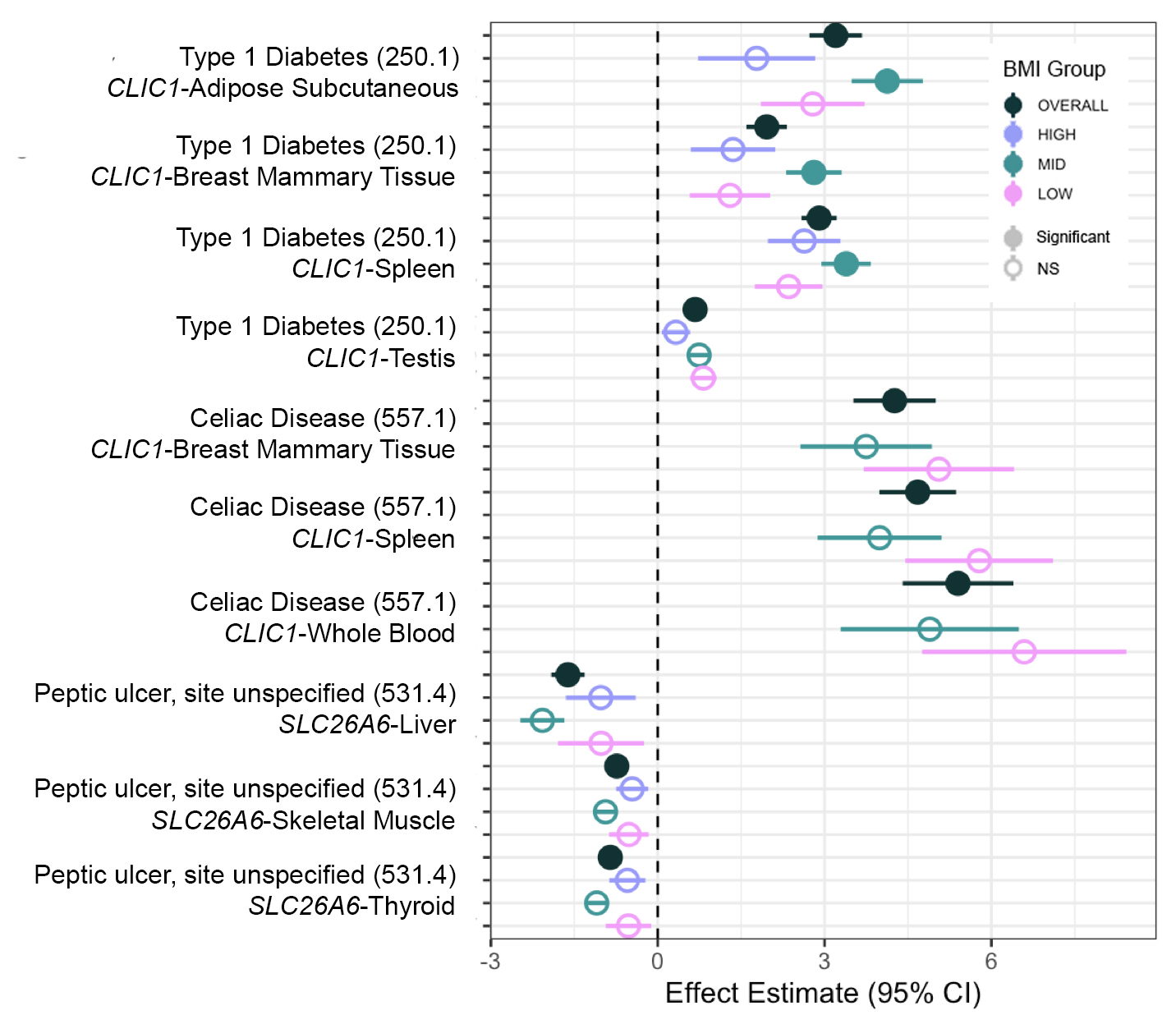

### Supplemental Figure 8

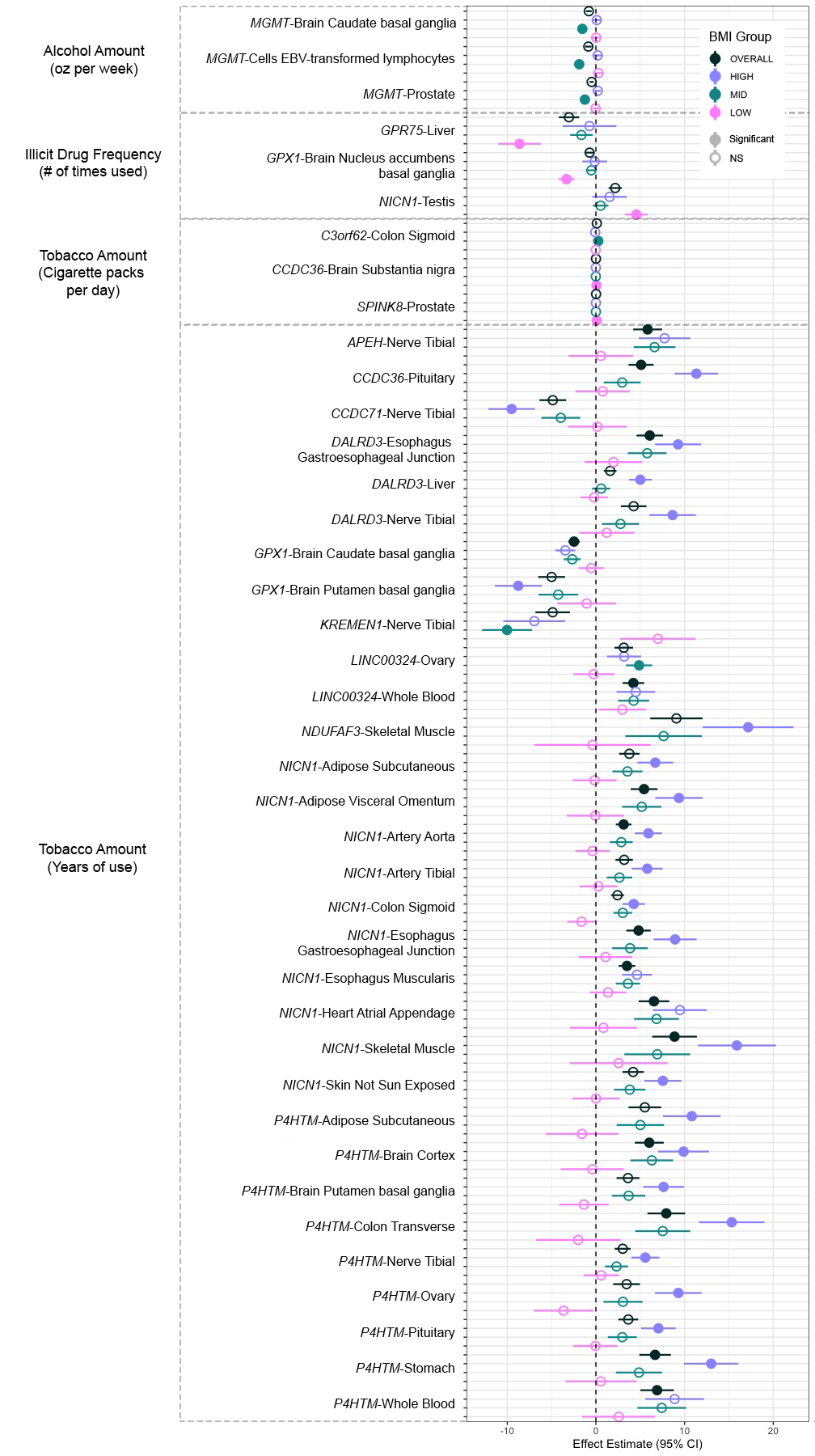

### Supplemental Figure 9

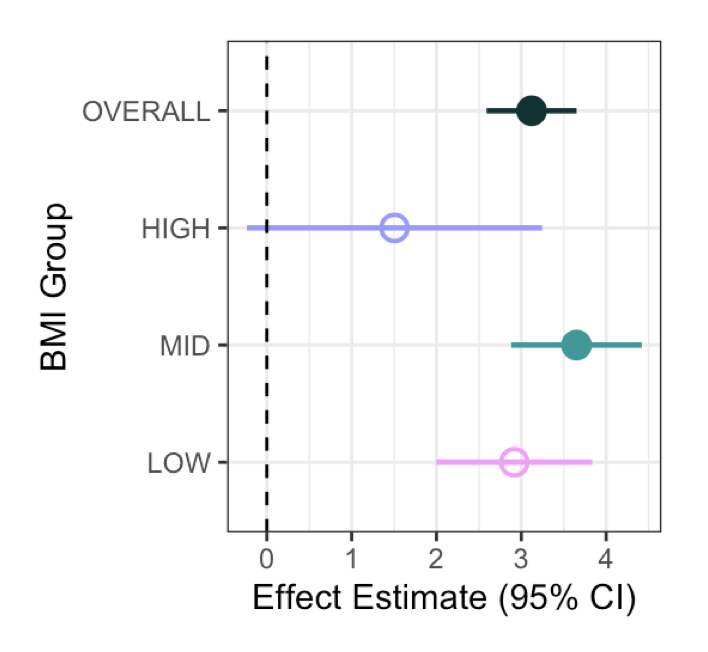

### Supplemental Figure 10

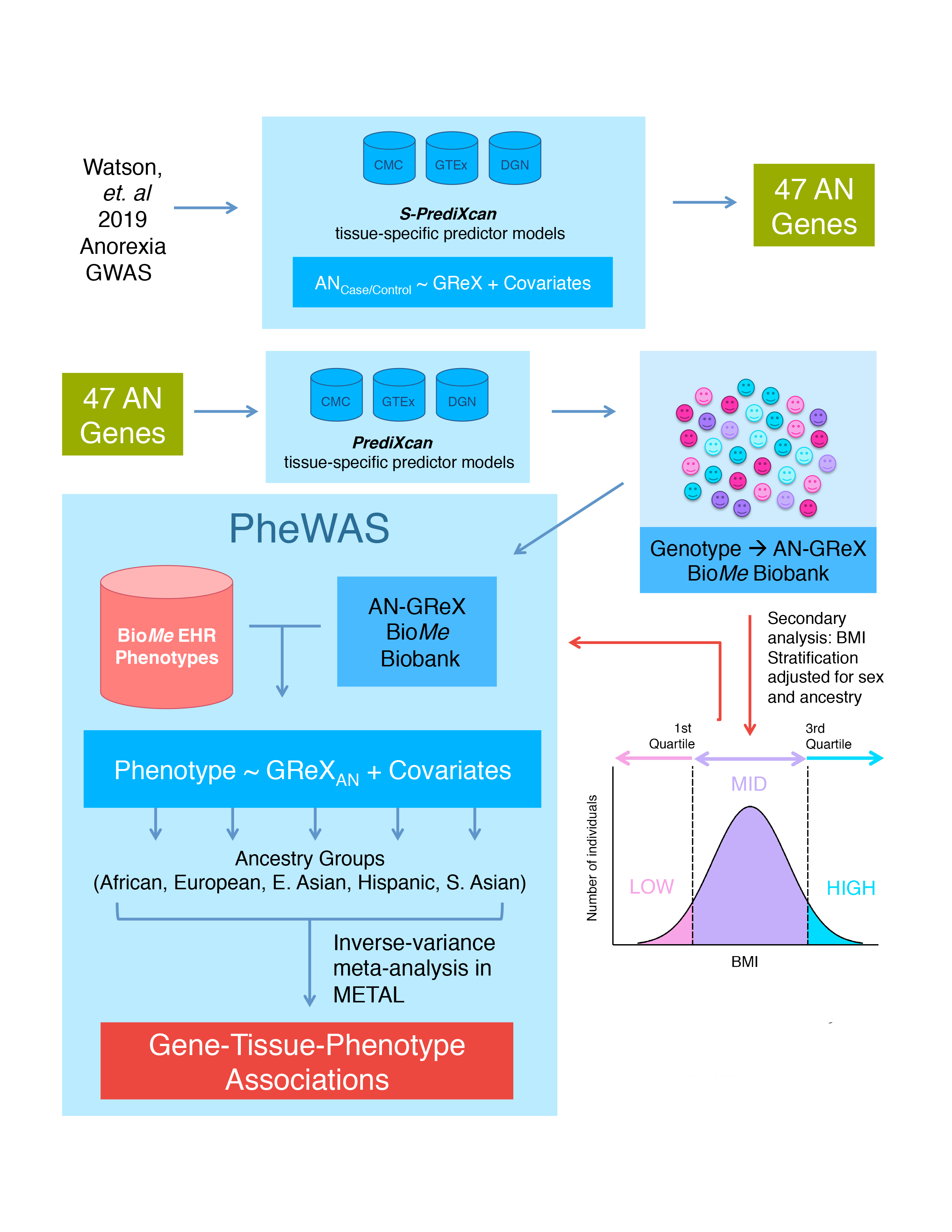

### Supplemental Figure 11

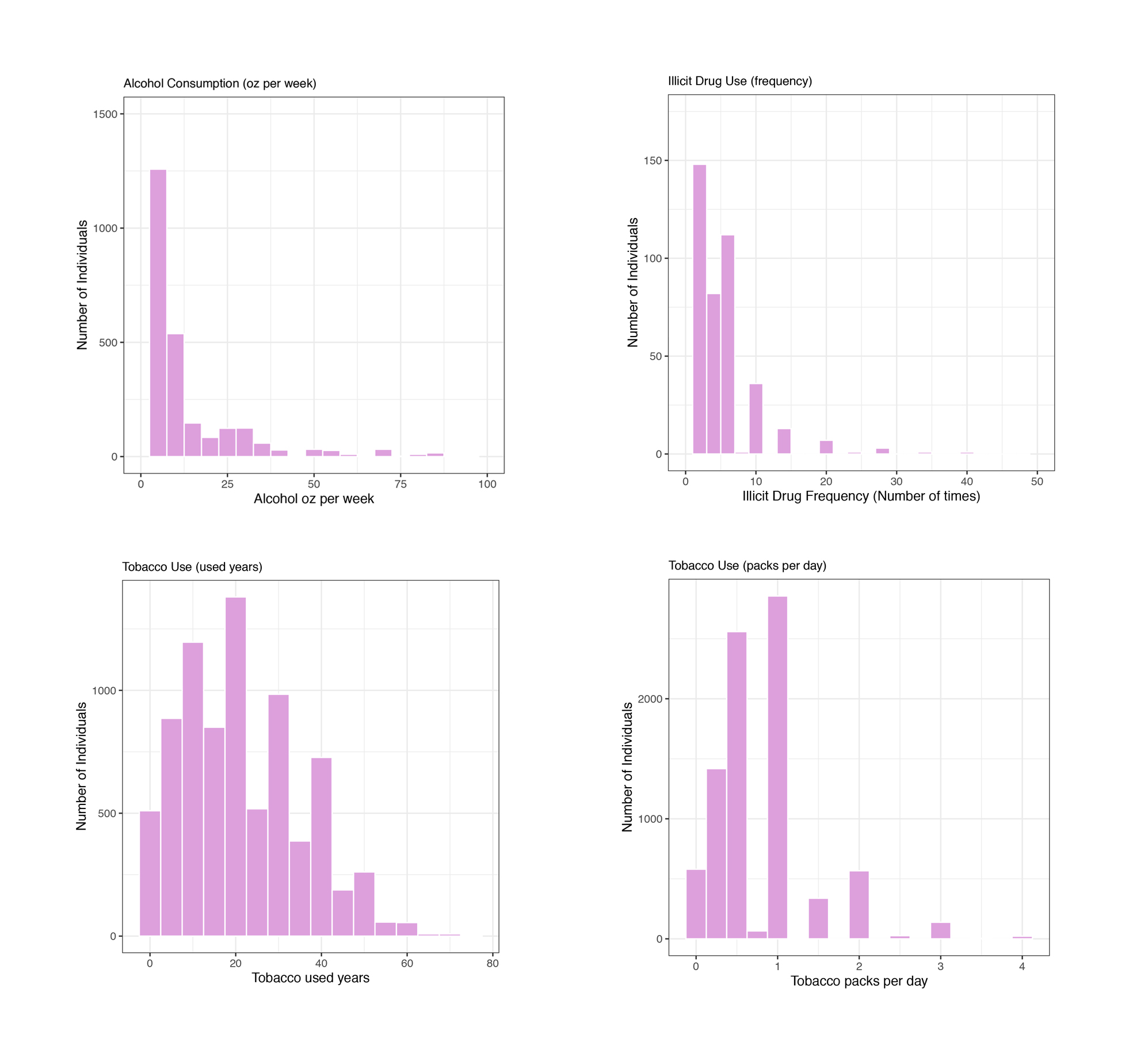

### Supplemental Figure 12

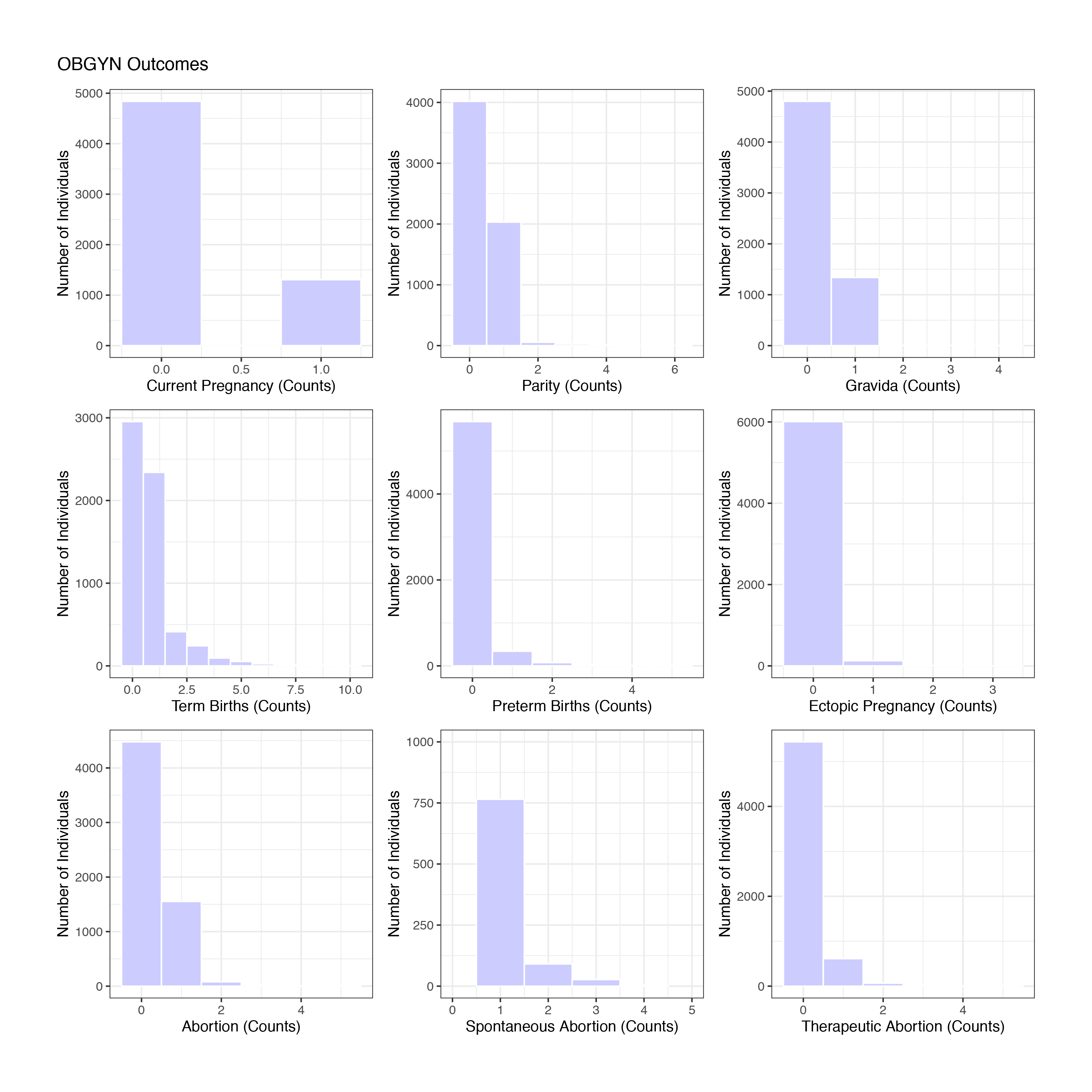

### Supplemental Figure 13

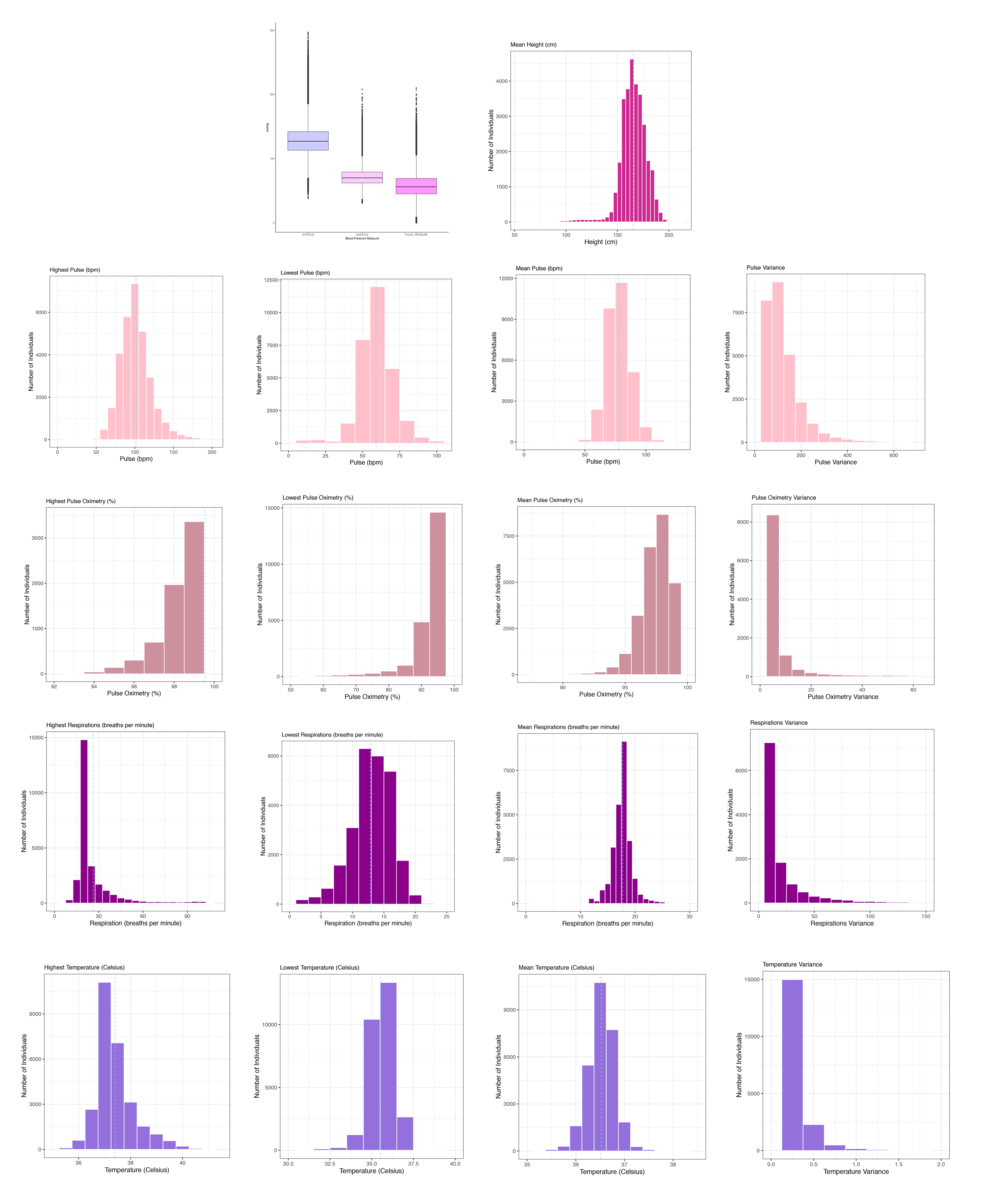

### Supplemental Figure 14

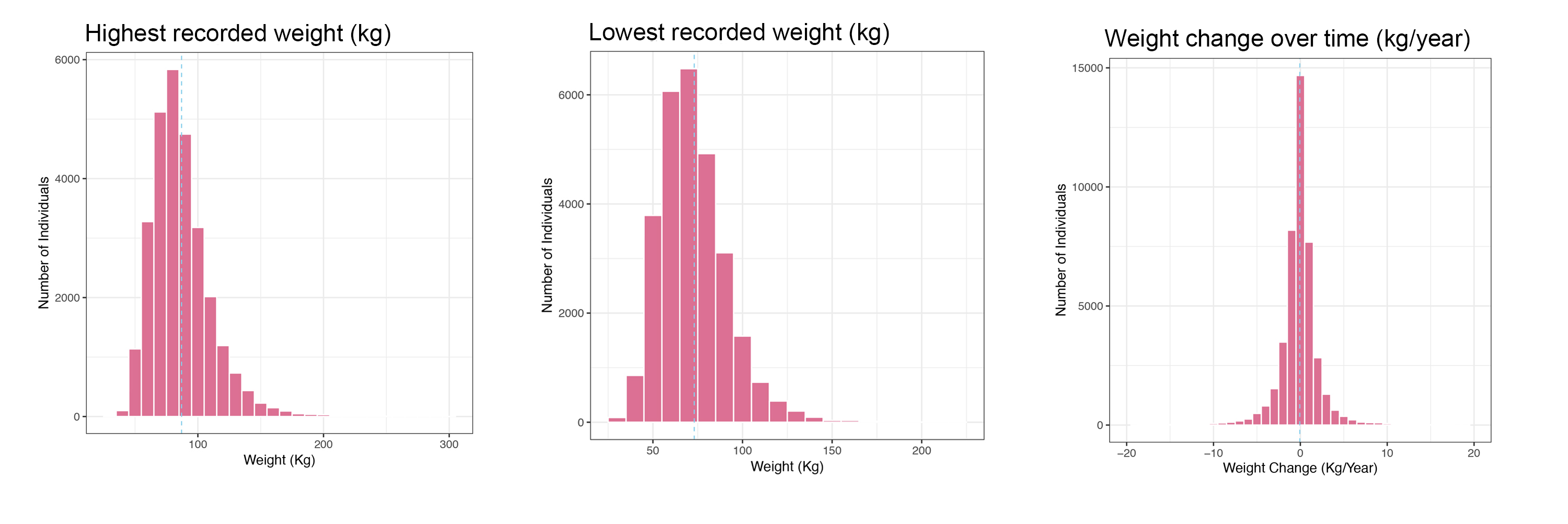

### Supplemental Figure 15

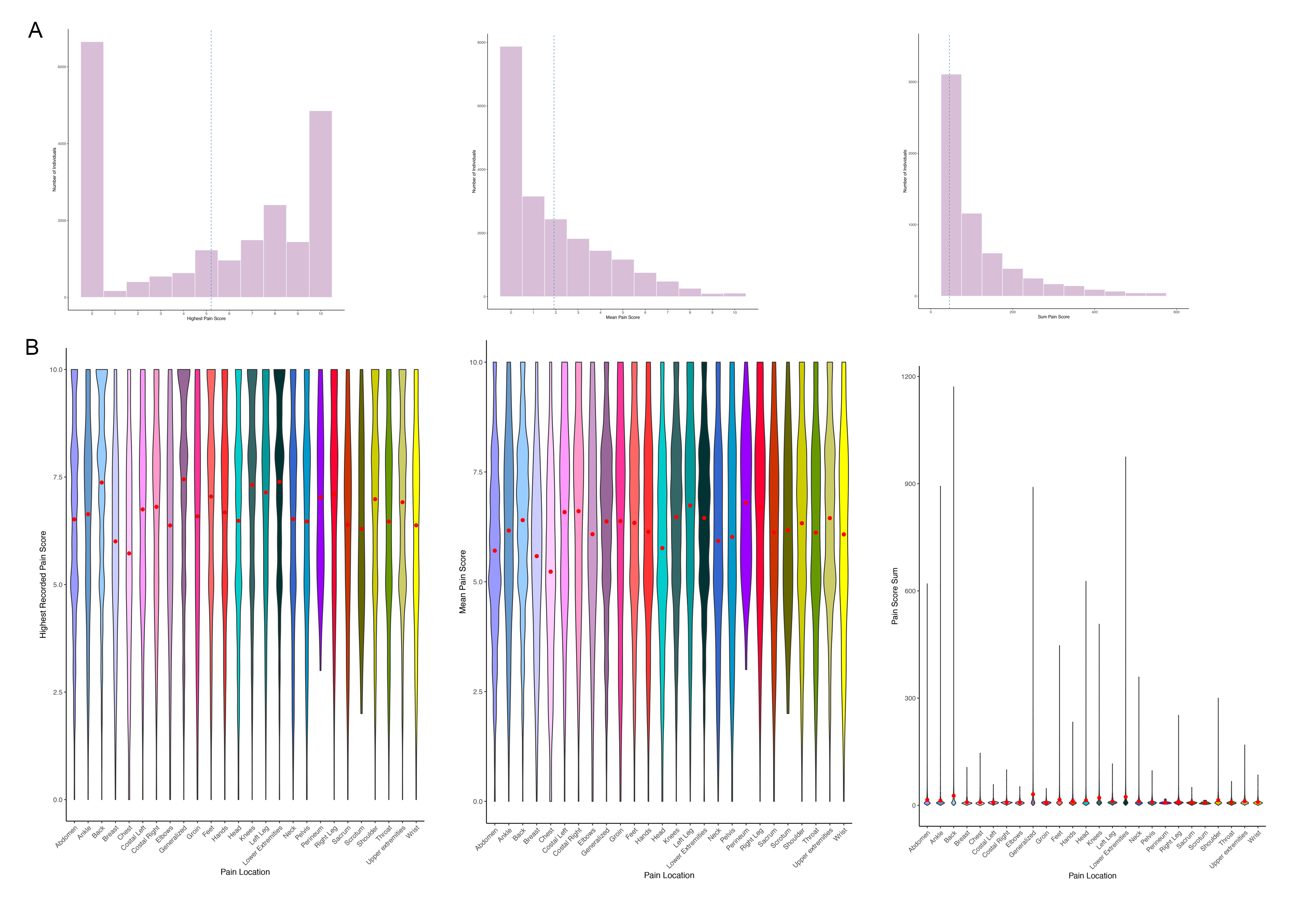

### Supplemental Figure 16

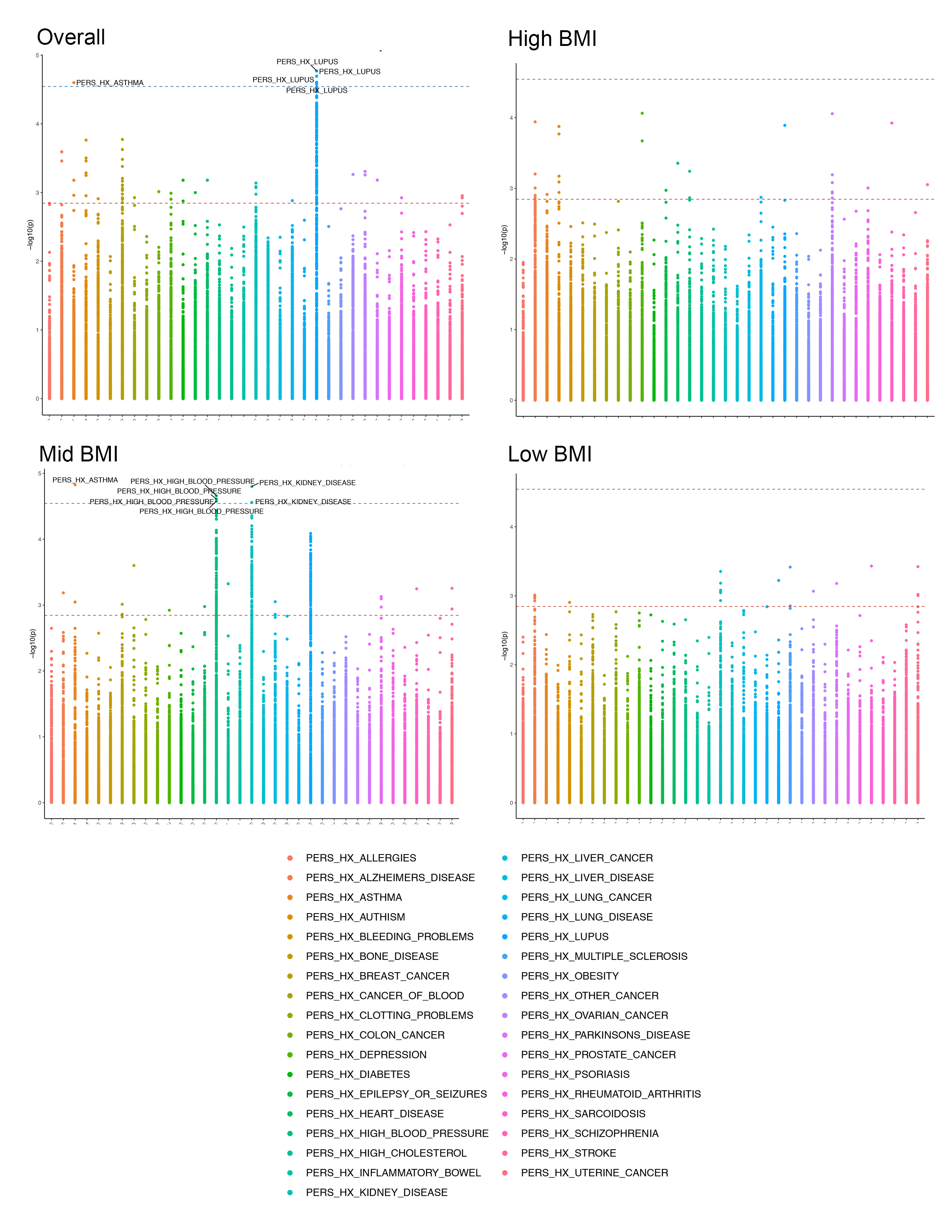

### Supplemental Figure 17

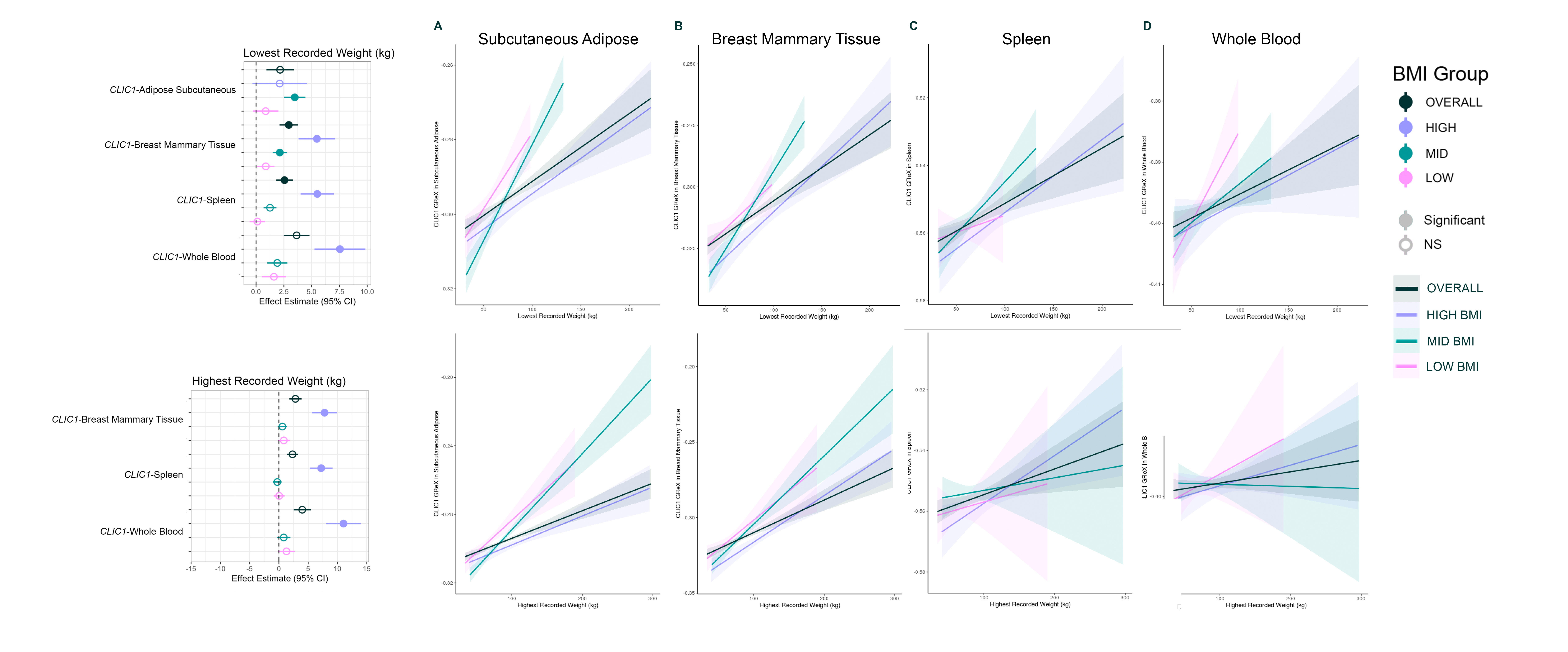

### Supplemental Figure 18

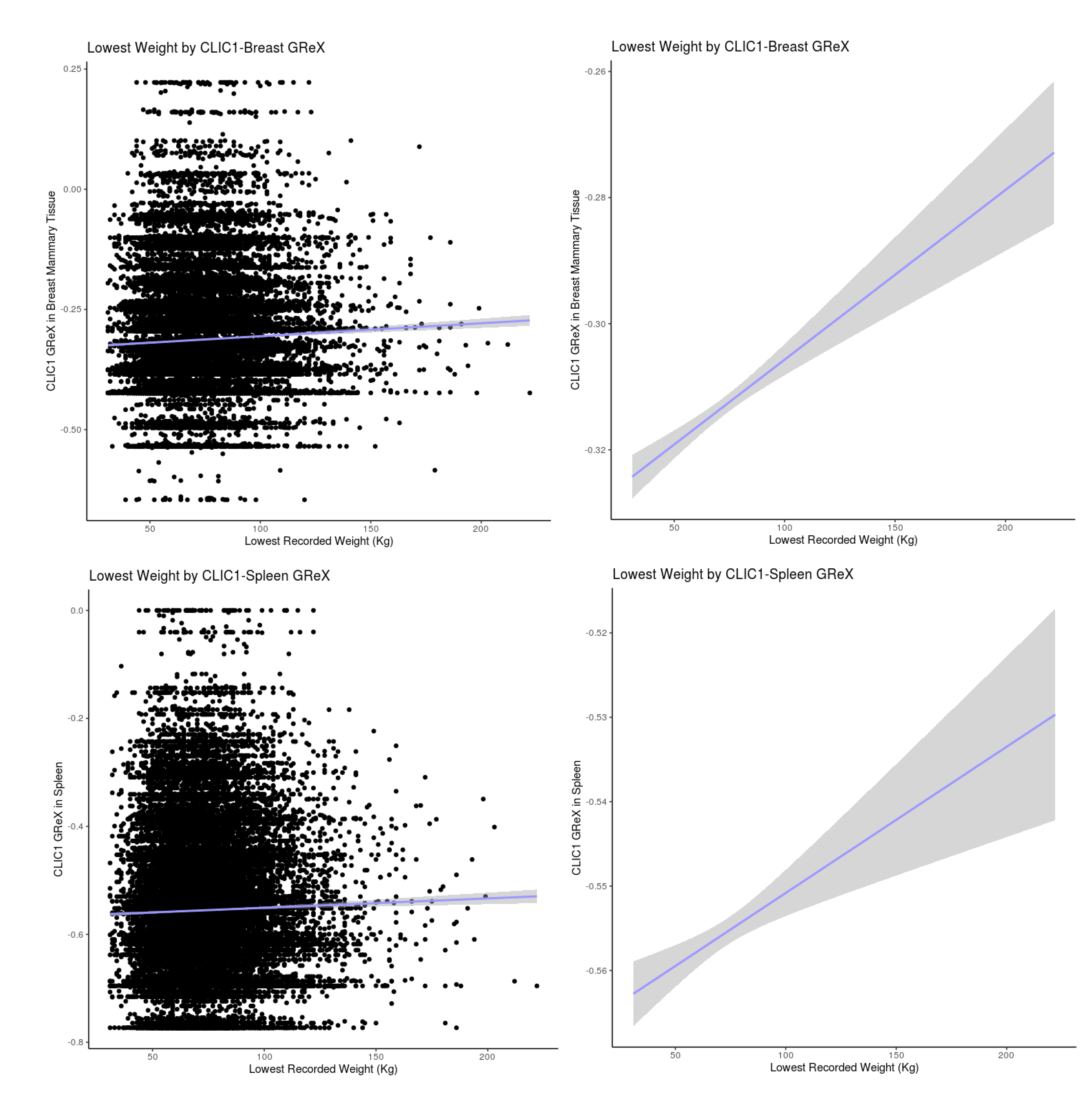

### Supplemental Figure 19

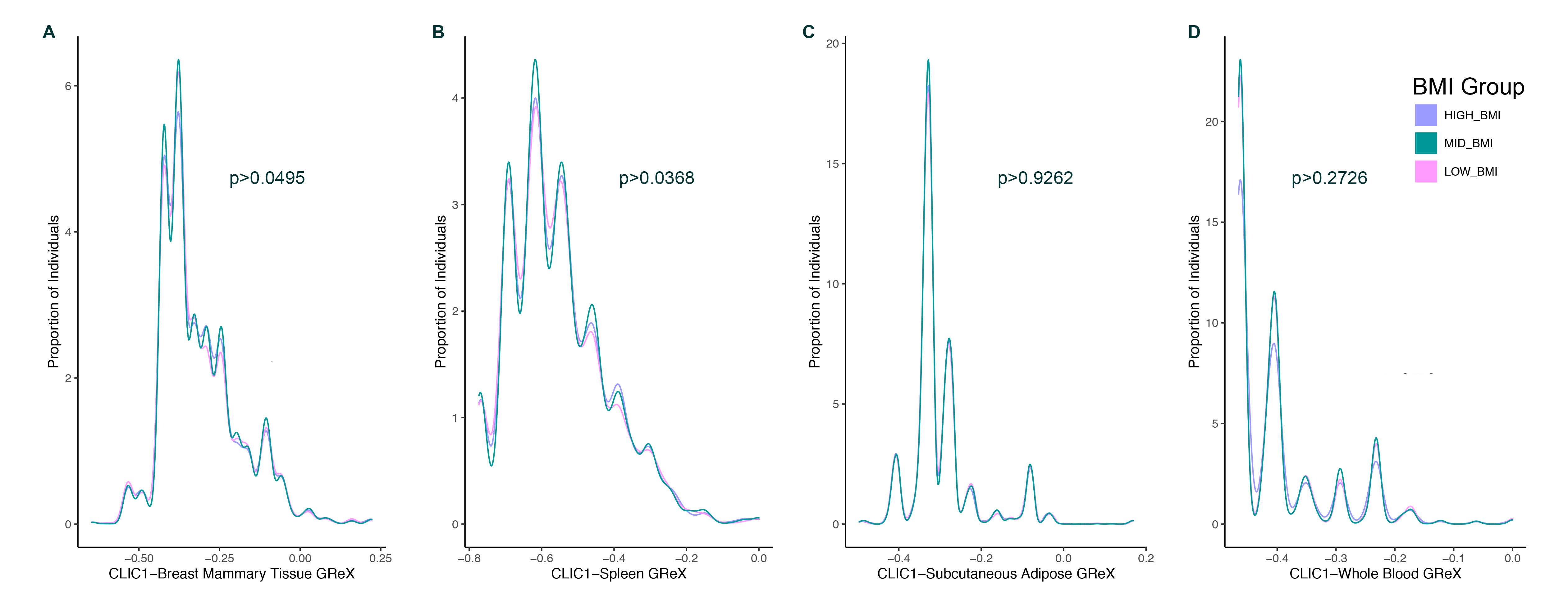

### Supplemental Table 2

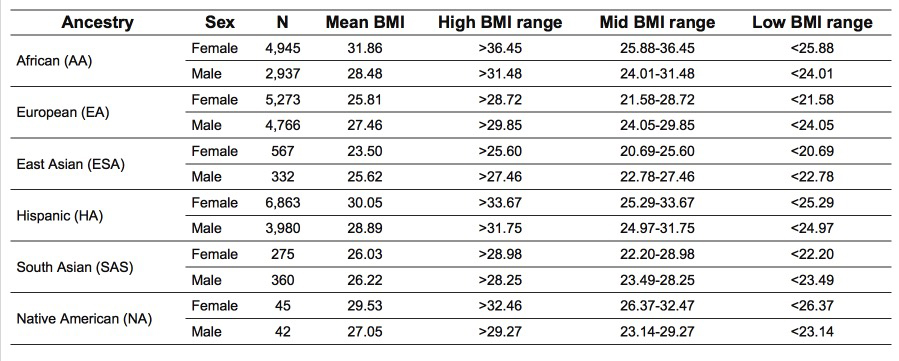

### Supplemental Table 3

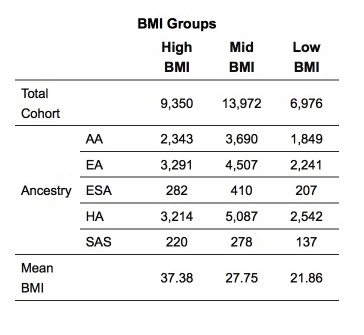

### Supplemental Table 19

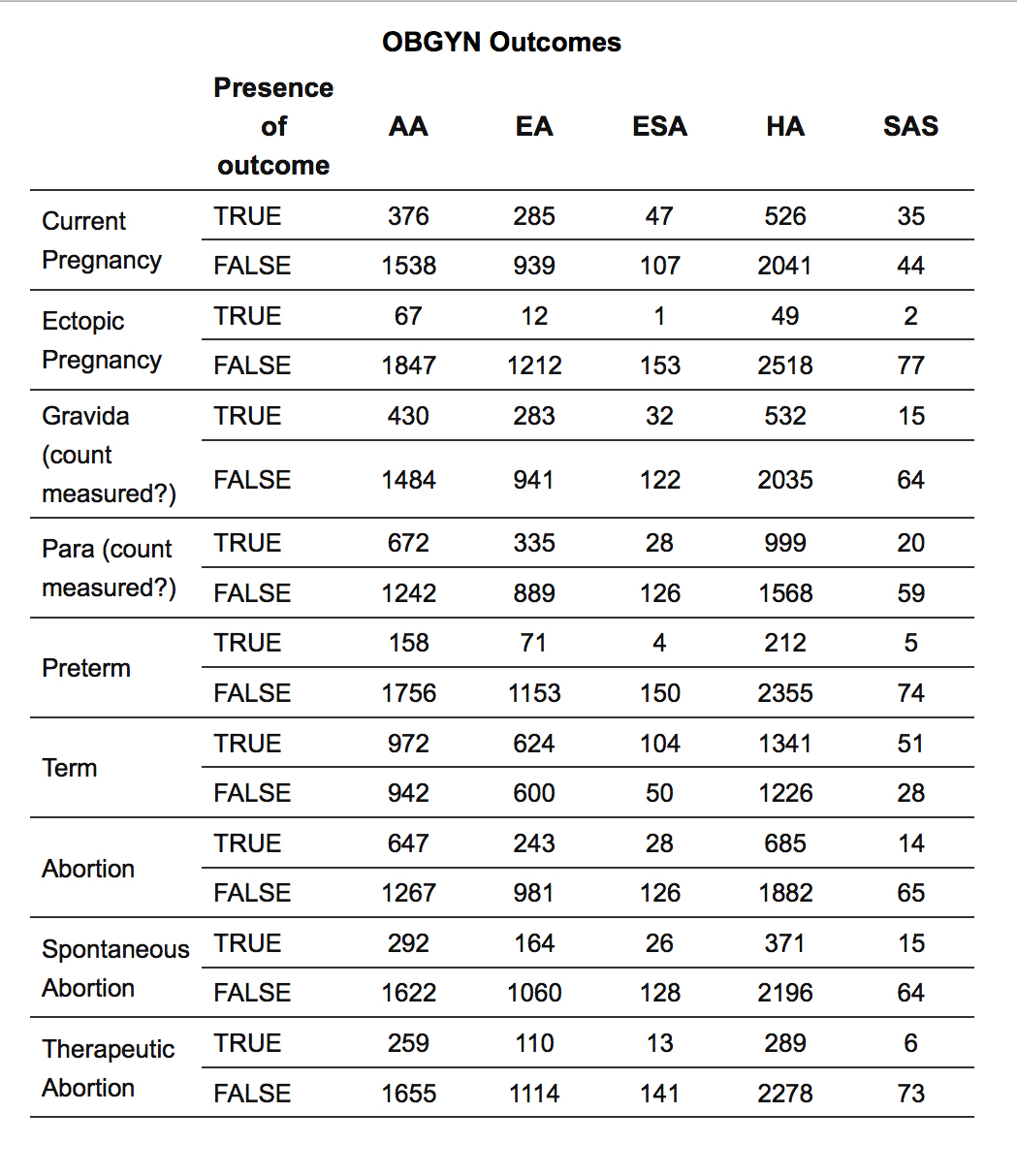

### Supplemental Table 20

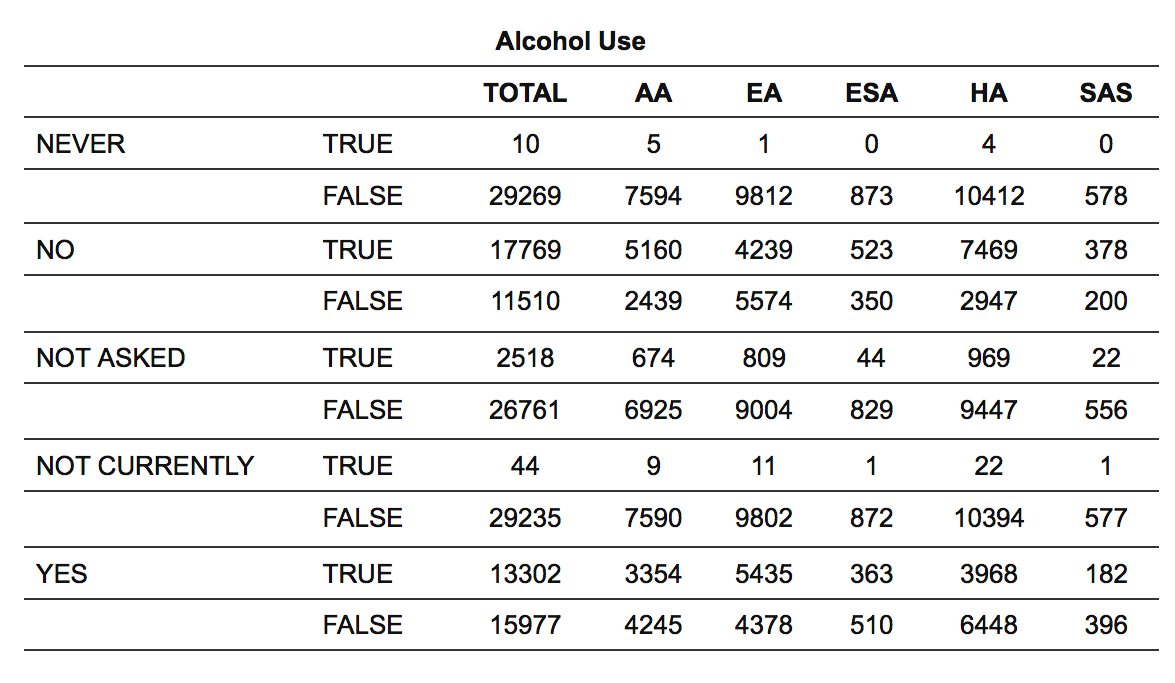

### Supplemental Table 21

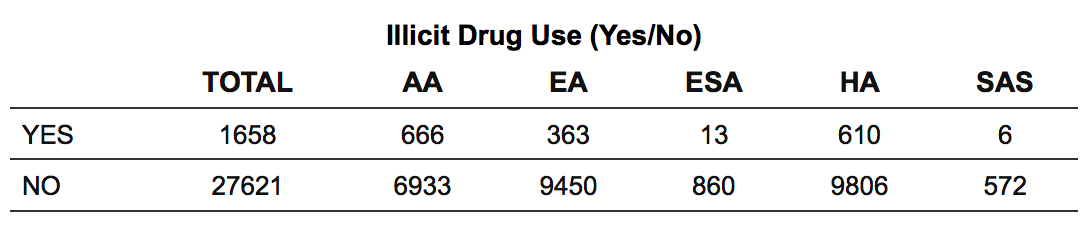

### Supplemental Table 22

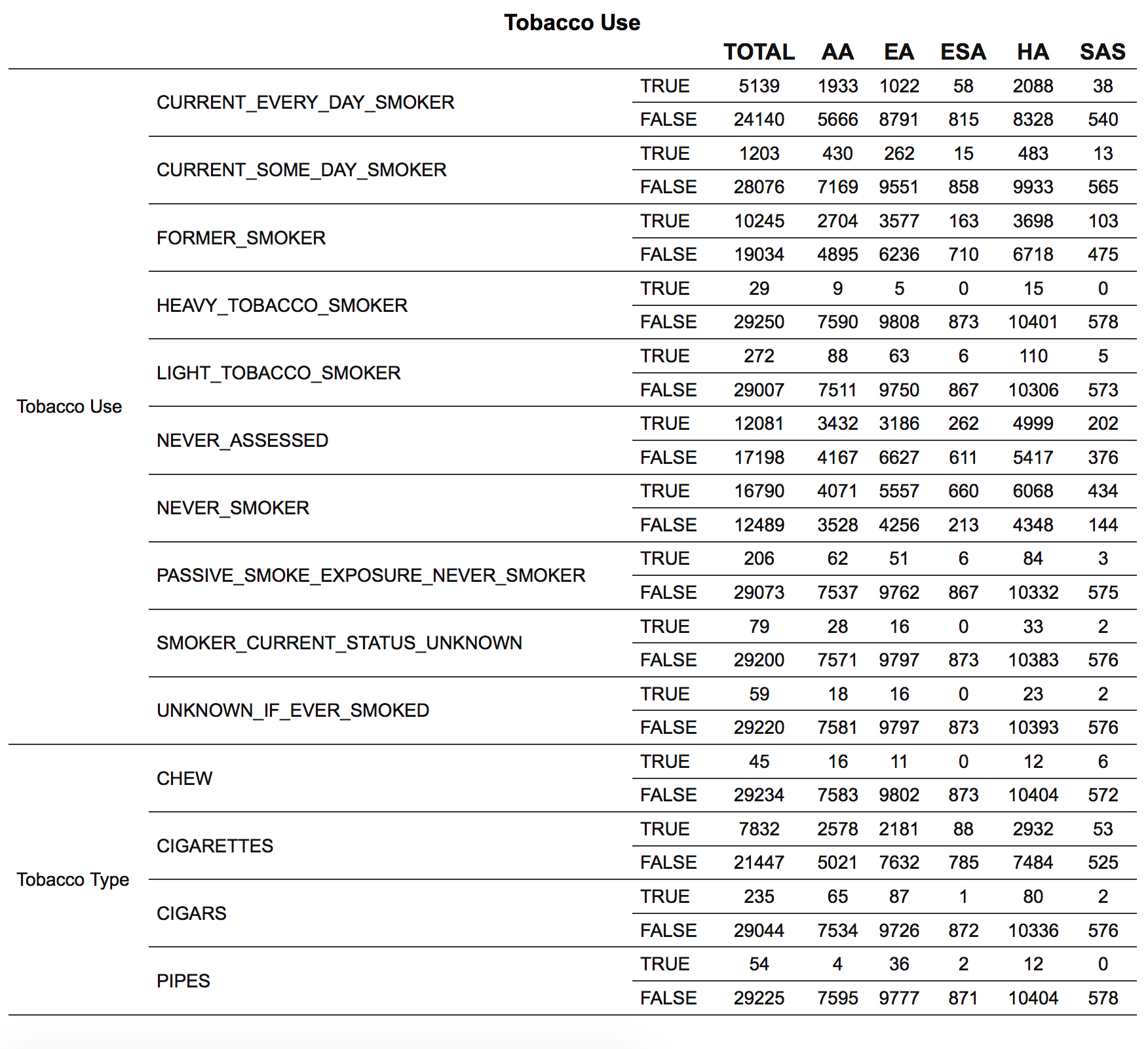

### Supplemental Table 23

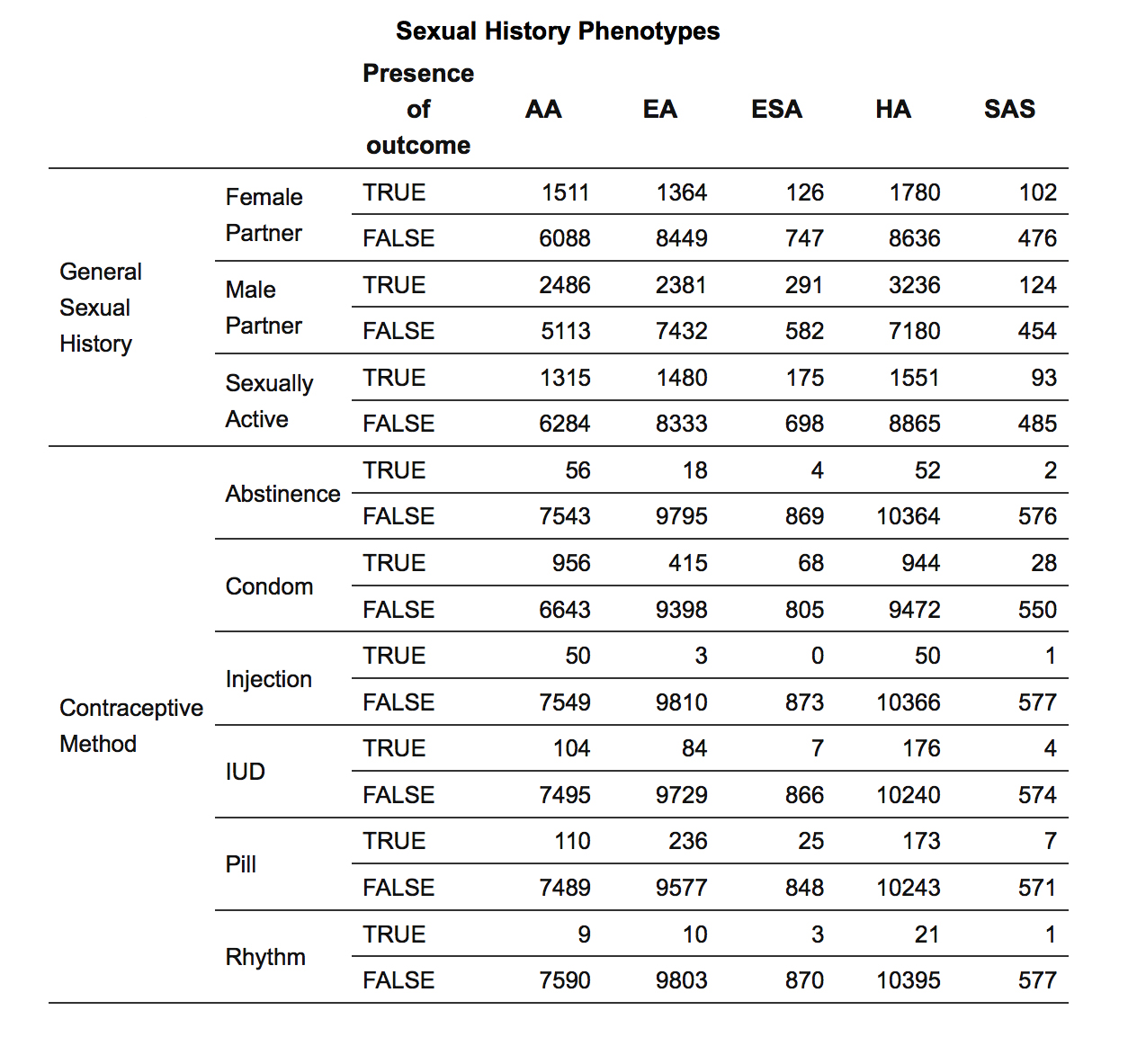
